# A risk-of-contagion index using a Bayesian based model for the COVID-19 epidemic in Mexico

**DOI:** 10.64898/2026.06.09.26355274

**Authors:** Ruth Corona-Moreno, M. Adrian Acuña-Zegarra, Mario Santana-Cibrian, Jorge X. Velasco-Hernandez

**Affiliations:** Instituto de Matemáticas, Universidad Nacional Autónoma de México, Juriquilla, Querétaro, Mexico; Departamento de Matemáticas, Universidad de Sonora, Blvd. Luis Encinas y Rosales S/N, 83000, Hermosillo, Sonora, Mexico; Escuela Nacional de Estudios Superiores, Universidad Nacional Autónoma de México, Juriquilla, Querétaro, Mexico

## Abstract

During the COVID-19 pandemic, limited testing capacity and reporting delays complicated epidemic surveillance and decision-making in Mexico. We calibrated *covidestim*, a Bayesian nowcasting model, to estimate the total SARS-CoV-2 infections from reported cases and deaths using Mexican surveillance data. Disease-progression distribution priors were calibrated using Mexico City records and validated through comparisons with national seroprevalence surveys, hospitalization data, and annual reported severe-case rates across all states.

Using the reconstructed estimates of active infections, we implemented an event-based risk framework that quantifies the probability of encountering at least one infectious individual in gatherings of different sizes. This probability was subsequently translated into a four-level epidemiological traffic-light indicator and computed at both state and municipality levels. The resulting estimates revealed substantial spatial heterogeneity that is obscured by state-level aggregation, particularly in states with marked differences between urban and rural municipalities.

To evaluate consistency with public-health indicators, we compared the proposed risk classification with the official Mexican epidemiological traffic-light system, considering interpretable gathering sizes relevant to public-health decision making.

Weekly reports derived from this framework were delivered to policymakers in the State of Queretaro in Mexico, as an anticipation tool for school reopening and public-space management. This demonstrates that this Bayesian reconstruction of infections combined with event-based risk metrics can provide an interpretable and generalizable municipality-level complement to routine surveillance systems, particularly in regions with limited testing capacity and heterogeneous local transmission dynamics.

## 1 Introduction

During the early phase of the COVID-19 pandemic, before vaccines were widely available and in the absence of broadly accessible effective treatments, non-pharmaceutical interventions were the primary tools to reduce transmission. Many countries implemented strict lockdowns to limit contact rates and population mobility. In Mexico, however, sustained and stringent confinement was not feasible for a large fraction of the population due to the large informal labor sector [1]. In addition, limited digital infrastructure and the social costs of prolonged school closures increased pressure to reopen educational activities, particularly among middle- and low-income households [2].

Under these constraints, public health authorities sought practical strategies to communicate epidemic risk and support individual and institutional decision-making. The Mexican government implemented a weekly epidemiological “traffic-light” system intended to summarize epidemic severity at the state level and guide recommendations on mobility, gatherings, and the operation of schools and public spaces [3] (see Appendix A).

This official indicator was implemented from June 1, 2020 to April 18, 2022 and relied primarily on reported cases and healthcare system metrics. However, large-scale testing and systematic surveillance of asymptomatic infections were limited, an issue common in many middle-income countries, so reported data did not fully capture the true incidence of infection. In addition, delays in diagnosis and reporting further hindered accurate near-real-time assessment.

Moreover, the traffic-light system was defined and officially published at the state level, implicitly assuming homogeneous conditions within each state, despite substantial heterogeneity across municipalities in terms of population density, connectivity, and access to healthcare. This spatial aggregation can mask the transmission dynamics at the municipal level and dilute important signals, potentially leading to misperceptions of risk, particularly in rural areas.

Although the official methodology for the traffic-light system was publicly available, some of the information required for its implementation was not accessible, particularly data related to healthcare system occupancy. As a result, it was not possible to independently estimate the traffic-light status of a given state in advance or to apply the methodology at the municipal level. This posed a challenge for decision-makers who needed advance estimates to prepare and implement public health measures or to plan the reopening of public spaces in response to increasing public demand.

To address these limitations, we adapted a Bayesian reconstruction model (*covidestim*)[4] to infer the total number of daily SARS-CoV-2 infections in Mexico, including unobserved infections, using weekly reported cases and deaths while avoiding reliance on healthcare system occupancy data.

As the open official COVID-19 reported data included all individual records around the country, identifying their state and municipality of residence, we infer the total number of cases at both the state and municipal levels. This gave us an estimate of the total active cases in each location at certain day, which we used to compute an interpretable probability: the risk of encountering at least one infectious person in a gathering of size *k*.

In this way, by assigning a traffic-light color scale to this risk of exposure, we were able to propose alternative epidemiological traffic-light systems to the official one, for gatherings of various sizes at the municipal and state levels.

Our main objective was to provide a reproducible, and municipality-resolved risk metric regarding size of gatherings that complements routine surveillance and supports more informed decisions by policymakers and the general population.

This paper is organized as follows. In Section 2, we describe the models used to produce our estimates. In Section 3, we detail the Mexican data, as well as the calibration and validation of the models for this context. In Section 4, we present the resulting risk estimates at both state and municipality levels based on inferred true case counts, which were communicated to policymakers during the COVID-19 epidemic. Finally, Section 5 provides a discussion of our findings.

## 2 Methodology

### 2.1 Bayesian reconstruction of infections and infectious prevalence

*Covidestim* is a Bayesian modeling framework that reconstructs daily incident SARS-CoV-2 infections by accounting for reporting delays and time-varying case ascertainment [4]. It links latent infections to observed reported cases and deaths through a compartmental disease progression structure and stochastic observation models (typically negative-binomial likelihoods).

Conceptually, infections progress through clinically relevant stages (e.g., asymptomatic or latent, symptomatic, severe/hospitalized, and death). In parallel, diagnosis and reporting are modeled explicitly: infections may remain undiagnosed, be diagnosed after a delay, and be reported after an additional reporting delay (see Figure 1). Given probability distribution priors for the generation of secondary cases (e.g., a serial-interval distribution), *covidestim* jointly estimates latent infections, the effective reproduction number *R*_*t*_ and reconstructs time series for clinically and operationally relevant quantities [5].

**Figure 1.**
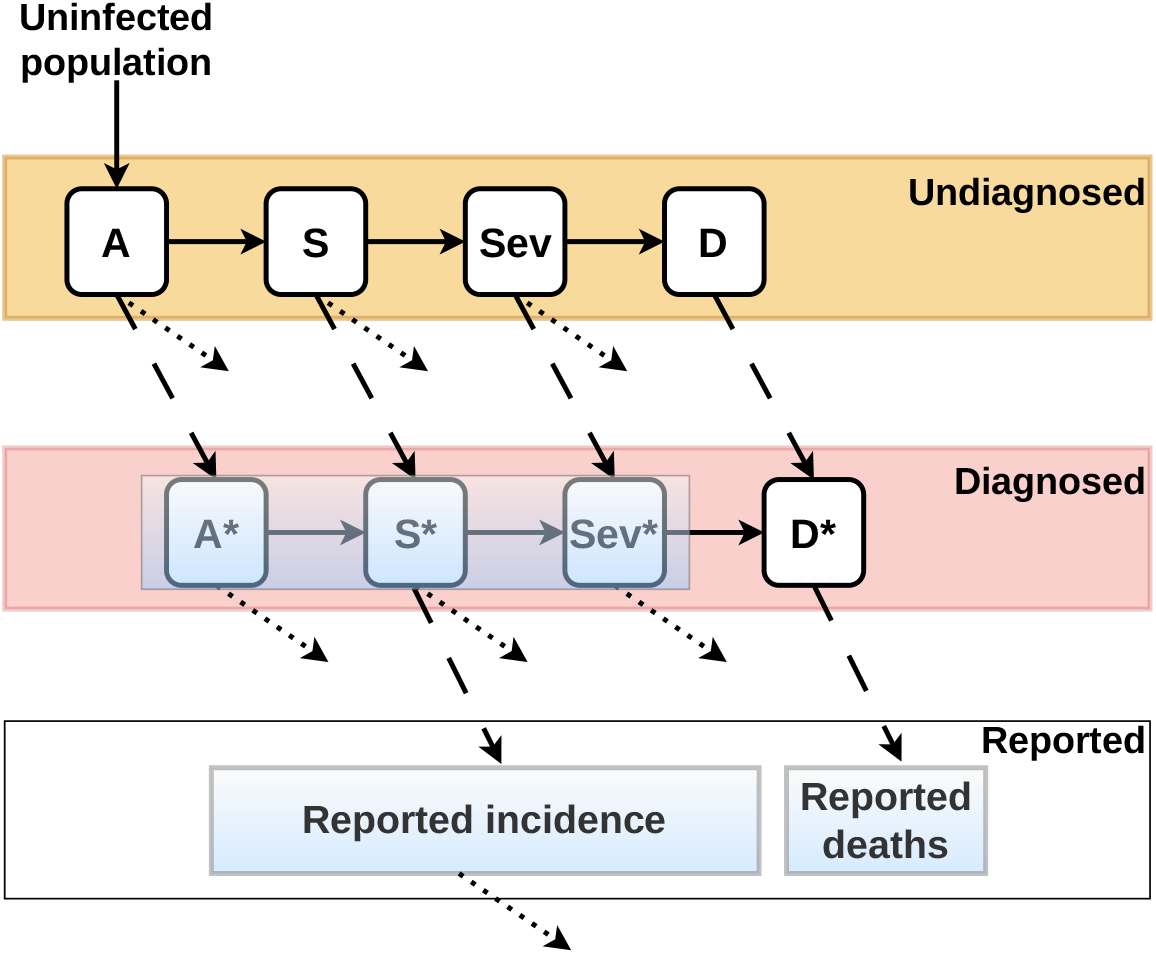
Compartmental structure of the covidestim model. Infected individuals progress through asymptomatic (A), symptomatic (S), severe (Sev), and death (D) states, with possible recovery (dotted arrows) at each stage. Cases may remain undiagnosed or become diagnosed (asterisk-marked compartments) and subsequently reported as confirmed COVID-19 cases. Dashed lines indicate transitions related to diagnosis and reporting. Adapted from Chitwood, M. H. *et al*.[4].

The *covidestim* implementation is distributed openly in R by its repository on GitHub [6]. Given (i) a population size, (ii) case and death time series (by symptom onset or report date) and (iii) prior distribution parameters of Table 1, the model returns posterior time series with associated 95% credible intervals for multiple outputs, including latent infections, adjusted and reported cases and deaths, *R*_*t*_, and measures of infectious prevalence or “population infectiousness” [7].

**Table 1.**
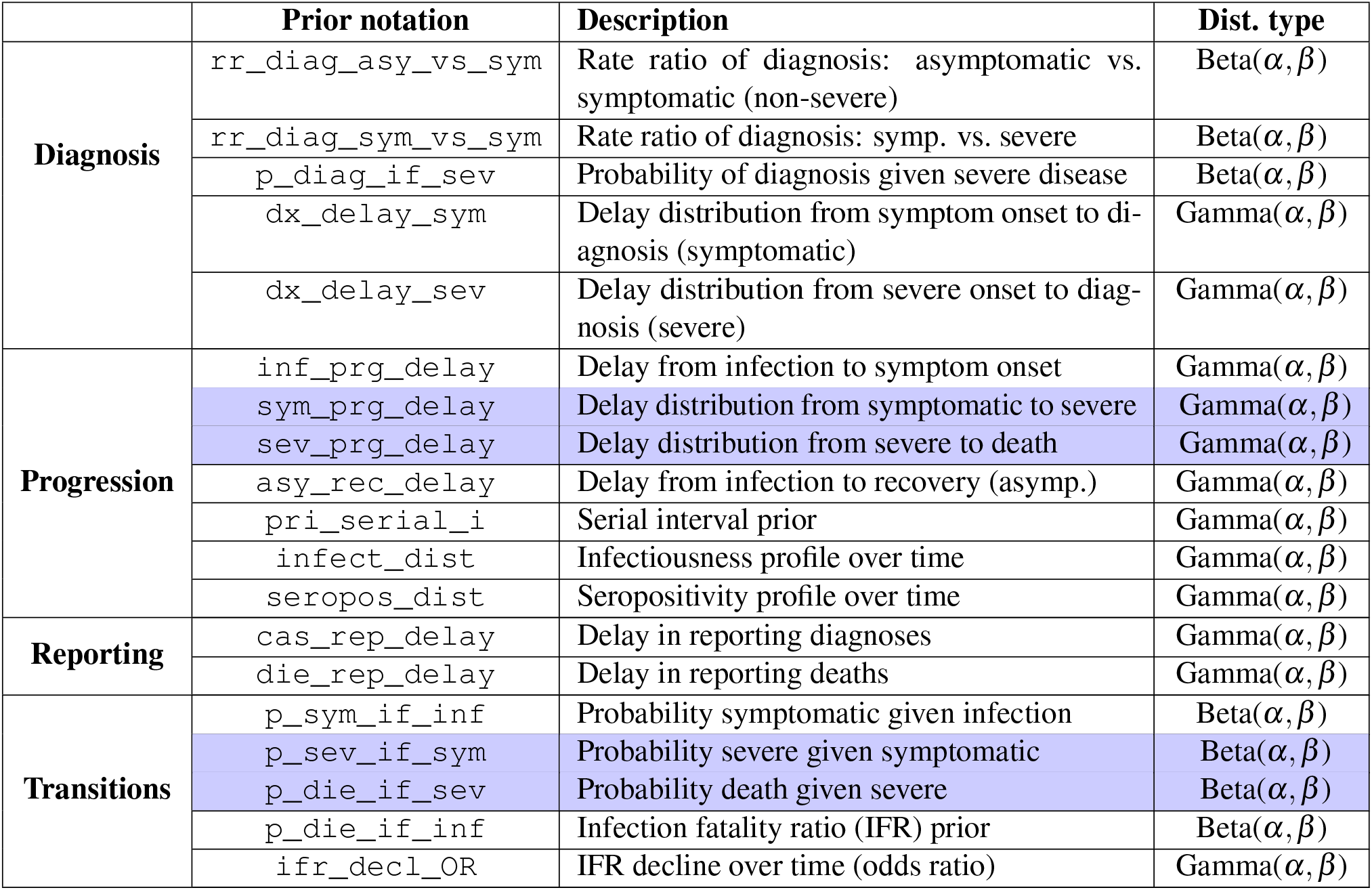
Configurable prior distributions in *covidestim*. Gamma parameters correspond to shape (*α*) and rate (*β*); Beta parameters correspond to shape parameters. Shaded rows indicate priors calibrated using Mexico City (CDMX) records and the others are taken from covidestim default configuration[7].

### 2.2 Event-based risk-of-contagion index

A practical way to communicate infection risk is to quantify the probability that a gathering includes at least one infectious person. Let *I*(*t*) denote the number of active infectious individuals in a locality on day *t*, and *N*(*t*) its population, which can be considered constant over the time. Define

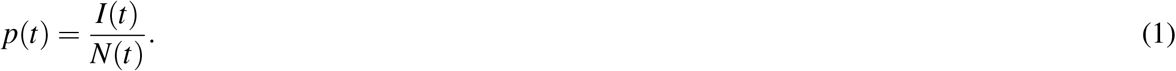

Under the simplifying assumption that attendees are sampled at random and independently from the population, the probability that a gathering of size *k* includes at least one infectious individual is

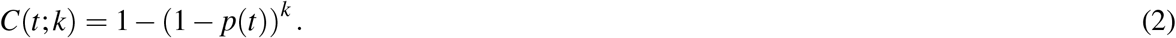

This quantity is closely related to “event risk” indicators proposed for public communication at county level in United States during COVID-19 epidemic[8].

## 3 Data and implementation in Mexico

During the pandemic, the Mexican Ministry of Health released a national COVID-19 open database with individual-level records [9]. Each record corresponds to a person identified by a unique numerical identifier, without direct personal identifiers, consistent with privacy guidelines. Variables include sociodemographic characteristics, comorbidities, indicators of severity, diagnostic information, and dates for symptom onset, registration, hospitalization, and death; definitions are provided in the official data dictionary [9].

We applied the methodology described above from November 2020 through October 2021, corresponding to the period before the completion of the first vaccination campaign for adults (18 years and older) [10]. In the database of these years we identified confirmed positive cases using categories 1–3 of the “final classification” variable (FINAL_CLASSIFICATION), following the official dictionary [9]. We constructed incidence time series by date of symptoms onset (FECHA_SINTOMAS) and mortality time series by date of death (FECHA_DEF). Because state and municipality of residence were recorded (ENTIDAD_RES and MUNICIPIO_RES), incidence and mortality were aggregated at state level and municipality level. Hospitalizations were derived from TIPO_PACIENTE=2 recorded by admission date (FECHA_INGRESO) and used for calibration and validation as described below.

### Software versioning and analysis window

We implemented *covidestim* using its second major version due to computational efficiency and the operational need for weekly reporting. Later updates incorporated vaccination effects, reinfections, and additional U.S.-specific parameterizations that were not directly supported or modifiable for Mexico in our workflow [5].

### Calibration of selected priors using Mexico City data

*Covidestim* relies on prior distributions governing delays and transition probabilities. Default values were largely informed by U.S. data. We therefore calibrated a subset of parameters using Mexico City (CDMX) records, leveraging its large case volume and relatively complete reporting during early pandemic waves. The calibrated priors (highlighted rows in Table 1) include delays from symptom onset to hospitalization, hospitalization to death, and key transition probabilities (symptomatic to severe; severe to death). Due to the lack of systematic testing, asymptomatic case tracking, and records of delays in reporting or test and recovery dates in Mexico, the distributions shown without shading in Table 1 cannot be estimated, so that we used the default values of covidestim configuration.

To estimate `sym_prg_delay`, we calculated the number of days between symptom onset and hospital admission among hospitalized individuals and fitted a gamma distribution using the `fitdistrplus` package in R. An analogous procedure was applied to estimate `sev_prg_delay`, using the interval between hospital admission and death.

For `p_sev_if_sym`, we computed the daily ratio of hospitalized to symptomatic individuals by date of symptom onset and fitted a beta distribution to these ratios. Similarly, `p_die_if_sev` was estimated using the daily ratio of deaths to hospitalized individuals, also indexed by date of symptom onset.

All distributions were fitted using data up to the 0.95 quantile of the empirical distribution to reduce the influence of extreme values. Goodness-of-fit was assessed using Q–Q and P–P plots, as well as comparisons between empirical and theoretical cumulative distribution functions (CDFs), presented in Appendix B.

The last calibration of these distributions for use in *covidestim* was performed using data from March 23, 2020 (the official date of school closures in Mexico) through January 31, 2021, corresponding to the early vaccination period in the country.

Table 2 presents the parameter estimates of the final fitted distributions, together with the default values used in the covidestim model. Figure 2 compares the empirical histograms with the fitted distributions and the covidestim defaults. The results show that the default covidestim distributions do not adequately represent the Mexican data.

**Table 2.** Calibrated prior parameters (CDMX fit) versus package defaults for the priors shaded in Table 1. The fitted distributions use Mexico City records from March 23, 2020 to January 31, 2021.

|  | Default | Default mean | CDMX fit | CDMX mean |
| --- | --- | --- | --- | --- |
| **sym_prg_delay** | G(1.624, 1.5225) | 0.65 | G(2.8574, 0.4762) | 2.09 |
| **sev_prg_delay** | G(2.061, 1.5939) | 0.62 | G(1.5064, 0.1672) | 5.97 |
| **p_sev_if_sym** | B(1.8854, 20.002) | 0.08 | B(7.1888, 55.0788) | 0.11 |
| **p_die_if_sev** | B(28.239, 162.3) | 0.03 | B(18.4077, 24.3287) | 0.43 |

**Figure 2.**
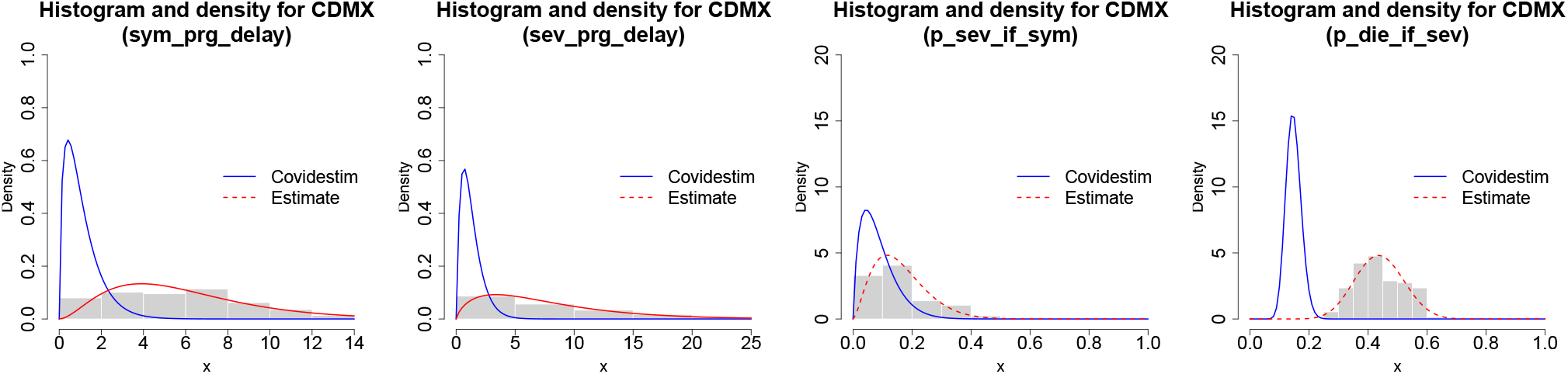
Empirical distributions and fitted priors for calibrated parameters in Table 2.

### Operational estimation and truncation

Given that, during the pandemic, the most recent data available at the time might have been incomplete due to delays in recording the onset of symptoms, diagnosis and reporting, the estimates for the most recent dates were not considered definitive. For weekly reports, we use estimates from up to seven days prior to the data download date as a near-real-time approximation, thereby improving timeliness compared to indicators that require longer retrospective periods.

### Computation

Weekly inference runs were executed on the supercomputer at the National Laboratory for Advanced Scientific Visualization at the National Autonomous University of Mexico (LAVIS, UNAM) [11].

### Validation

We compared estimated cumulative infections through November 2020 with national sero-prevalence from the National Health and Nutrition Survey (Ensanut), conducted August–November 2020 [12]. The survey estimated that approximately 24.7% of the national population had antibodies in November 2020, a value higher than the median cumulative infection estimate produced by *covidestim* for the same date. Nevertheless, the survey estimate falls within the posterior credible interval for cumulative infections, as indicated by the blue point in Figure 3a. Although our estimate was performed and validated until October 2021, if we consider the same calibration for covidestim until December 2021, we also observe the Ensanut survey of 2021 falls withing the credible interval, as the red point in Figure 3a shows.

**Figure 3.**
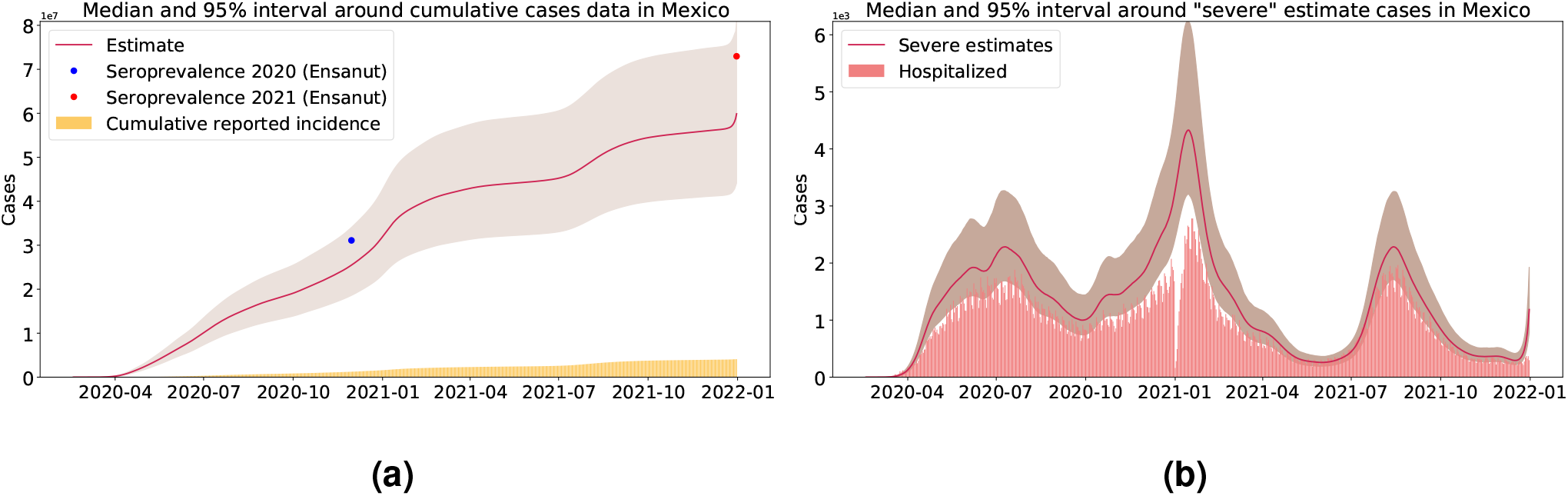
(a) Estimated and reported cumulative incidence in Mexico, with 95% credible intervals. The blue and red dots represent official seroprevalence estimates from the 2020 and 2021 “Ensanut” surveys, respectively. (b) Comparison between hospitalized records in Mexico and the covidestim estimated severe cases.

Moreover, we compared the severe infections estimated by covidestim with hospitalization records that were not directly used as model inputs, providing an additional face-validity assessment. The results showed good agreement, with hospitalization records closely matching the lower bound of the credible interval and following trends consistent with the estimated credible intervals (Figure 3b). The adjustment between covidestim estimate severe cases and hospitalization records for all states can be found as supplementary material.

Furthermore, Figure 4 shows that the annual hospitalization rates per 100,000 inhabitants for each state fall within, or are close to, the credibility intervals of the severe case rates per 100,000 inhabitants calculated using the covidestim estimates.

**Figure 4.**
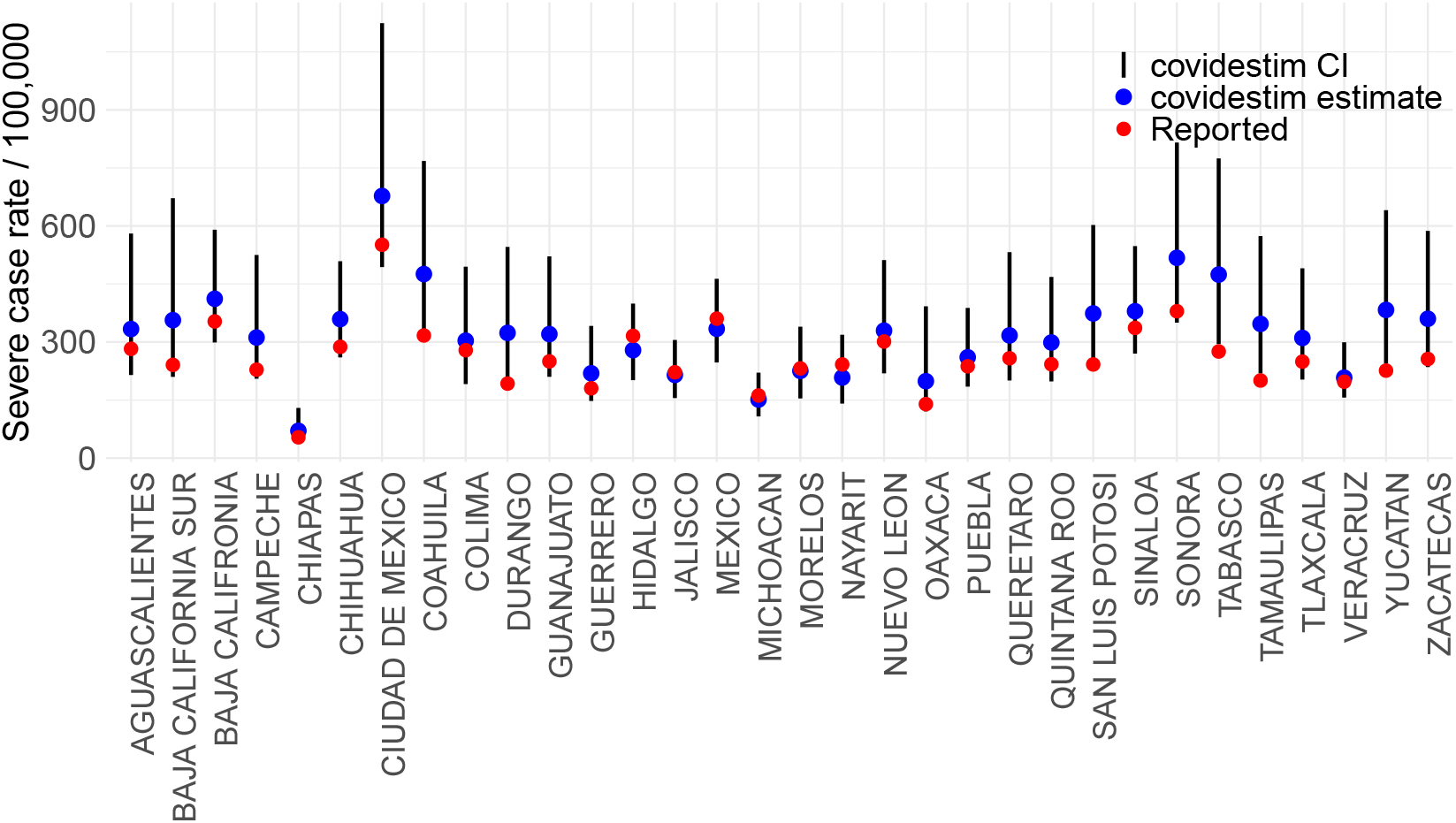
Comparison of the annual COVID-19 hospitalization rate recorded in 2020 with the credible interval of the covidestim estimates of severe cases.

Finally, the covidestim estimates of *R*_*t*_ were compared with those obtained using *EpiEstim* package [13]. While covidestim derives *R*_*t*_ from its reconstructed total infections (*A*_*t*_), the estimates from *EpiEstim* rely directly on reported case data. As shown in Figure 5, the covidestim trajectory of *R*_*t*_ shows similar temporal trends but appears smoother, reflecting the model’s regularization and joint inference framework. This pattern is consistently observed across all states (see Supplementary Material).

**Figure 5.**
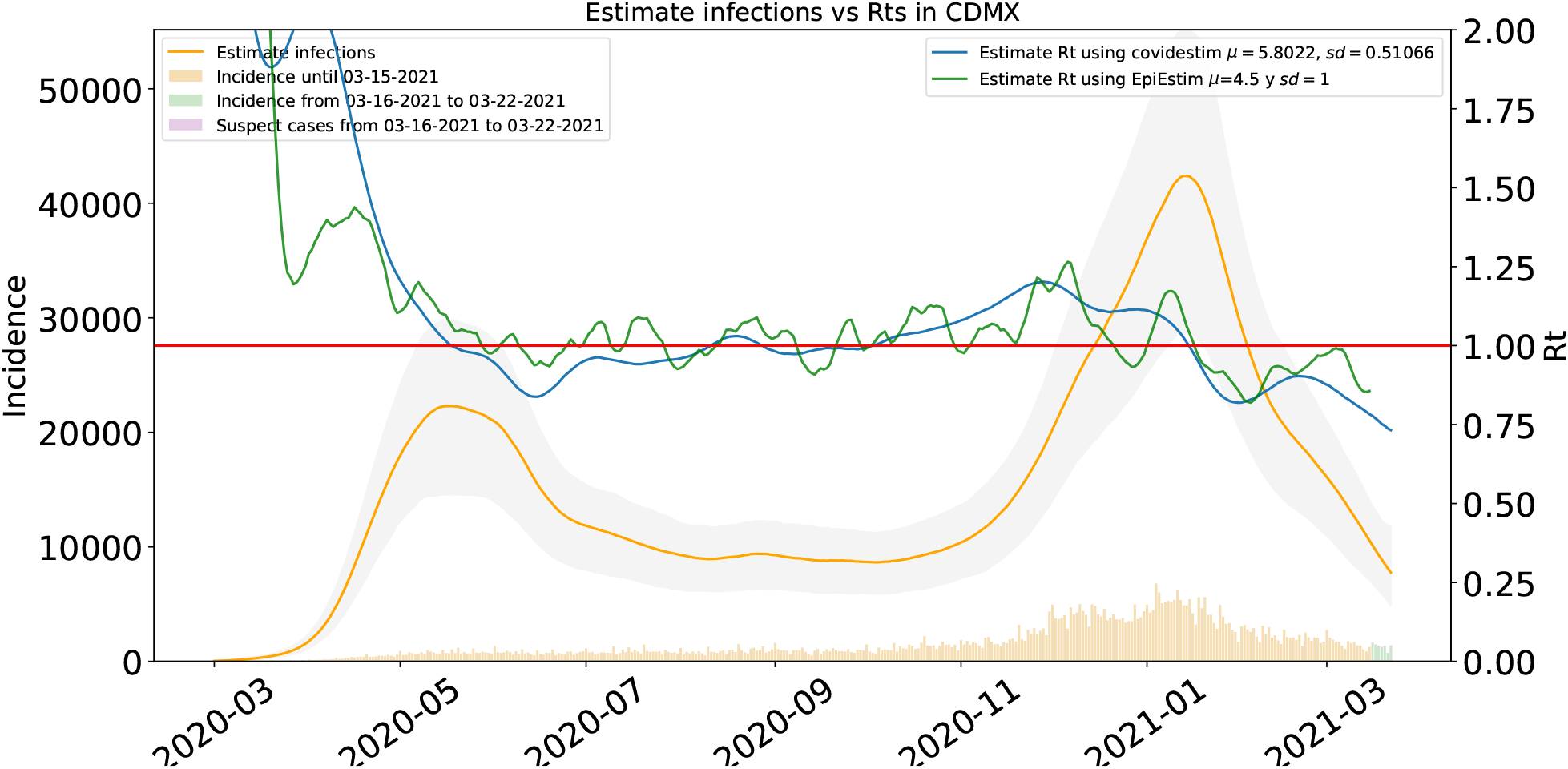
Illustrative weekly reconstruction for Mexico city (CDMX) through March 22, 2021, using data updated through March 29, 2021. Estimated infections and *R*_*t*_ in CDMX (posterior median and 95% CrI).

Overall, these comparisons support the plausibility of the reconstructed clinical progression.

### Risk scenarios and reporting choices

Once we obtained the covidestim `infections` estimates up to seven days prior to the data download date, we used the estimates from the preceding 14 days to approximate the total number of active cases, denoted by *I*(*t*) in equation (1). The quantity *N*(*t*) was defined as the constant total population of each location, based on the 2020 official population census conducted by INEGI (the Mexican National Institute of Statistics and Geography) [14].

Based on these quantities, and through their assignment to a traffic-light color scale, we generated alternative epidemiological traffic-lights for different gathering sizes using the estimated active cases up to seven days prior to the data download date.

To communicate event risk in settings relevant to policy and everyday decision-making, we computed and reported the risk *C*(*t*; *k*) for three gathering sizes (Table 3) representing typical capacities of indoor and public events. These categories are illustrative and not exhaustive.

**Table 3.** Gathering sizes used to compute the probability of encountering at least one infectious person in a gathering.

| Gathering size $k$ | Example setting |
| --- | --- |
| 40 | Classroom, public bus, or a small indoor social gathering |
| 100 | Small theater, lecture hall, or medium-size event venue |
| 400 | Large religious, political, or academic event |

Thereafter, to facilitate comparisons with Mexico’s official epidemiological traffic-light system, we transformed the continuous risk measure into a discrete four-level scale by evenly partitioning the interval [0, 100]. Risk values in [0, 25) were classified as green, those in [25, 50) as yellow, those in [50, 75) as orange, and those in [75, 100] as red. This categorization enhances interpretation while retaining the probabilistic nature of the underlying risk estimates.

## 4 Results

### 4.1 Reconstruction outputs

The validation of the previously described covidestim results enabled the generation of weekly estimates of the true epidemic dynamics for each state in Mexico, each municipality in Queretaro, and each borough in Mexico City, up to October 2021. The resulting reports are available in the section “Estimadores COVID-19” on the official website of the Multidisciplinary Node for Applied Mathematics (NoMMA) at the Institute of Mathematics of the National Autonomous University of Mexico (UNAM) [15]. These reports included visualizations such as those shown in Figures 3 and 5.

The methodology was subsequently extended to all municipalities in the country, and the corresponding figures can be found in the supplementary material.

### 4.2 Spatial heterogeneity in event risk

The disaggregated analysis reveals substantial heterogeneity in epidemic risk patterns at both the state and municipal levels, as we observe in Figures 6-8. These regional disparities highlight the importance of designing mitigation measures that address local epidemiological conditions, especially in regions where there is limited interaction between different population groups.

**Figure 6.**
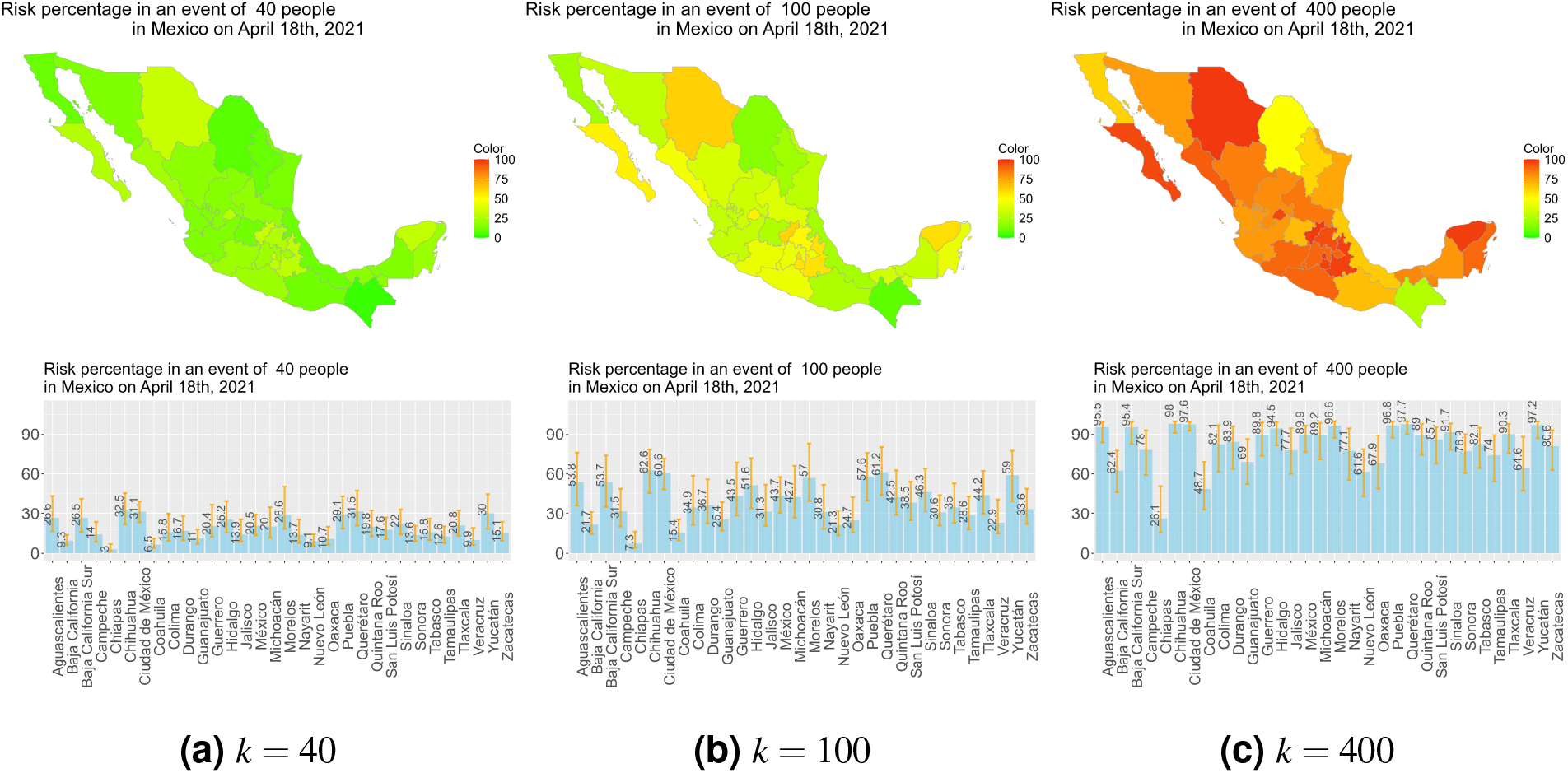
Risk estimate for gatherings size *k* = 40, 100, 400 across states of Mexico for April 26 to May 2, 2021.

In addition, incorporating gathering size into the risk traffic-light framework enhances the assessment of local epidemic severity. While some localities may maintain a low-risk level (green) for gatherings of both 40 and 400 individuals, other regions may experience a substantial increase in risk as gathering size increases. These differences reveal important heterogeneity in local transmission dynamics that may not be captured by risk classifications based solely on current incidence levels.

Such contrasts are evident at the state level when comparing Chiapas and Quintana Roo (Figure 6), and at the municipal level when comparing Huimilpan and Corregidora in the state of Queretaro (Figure 7).

**Figure 7.**
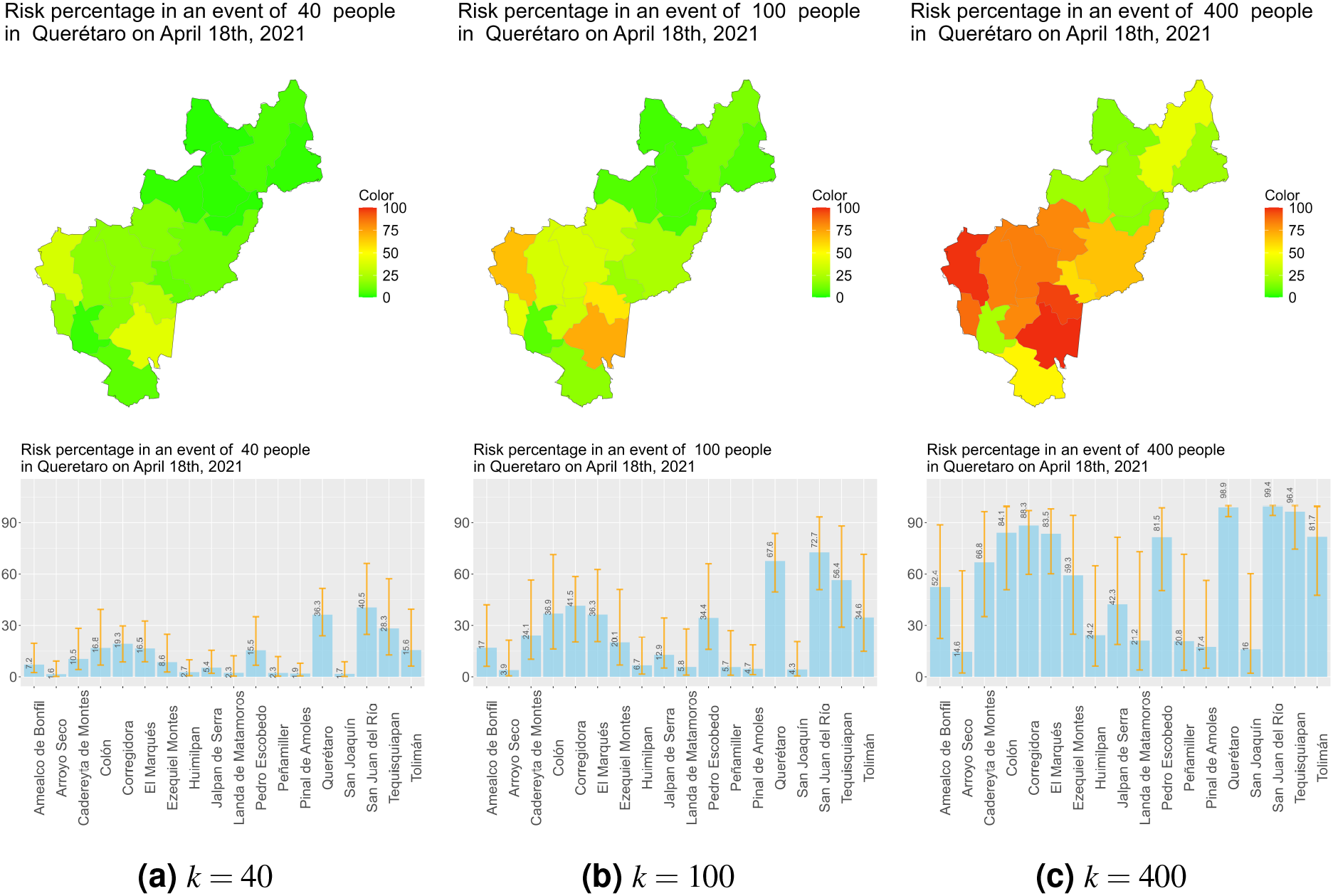
Risk estimate for gatherings size *k* = 40, 100, 400 across Queretaro for April 26 to May 2,2021.

However, such heterogeneity is not observed in all regions. For example, in Mexico City (CDMX), risk patterns are relatively homogeneous across all boroughs, which is likely associated with the high level of connectivity and population mixing throughout them. See Figure 8.

**Figure 8.**
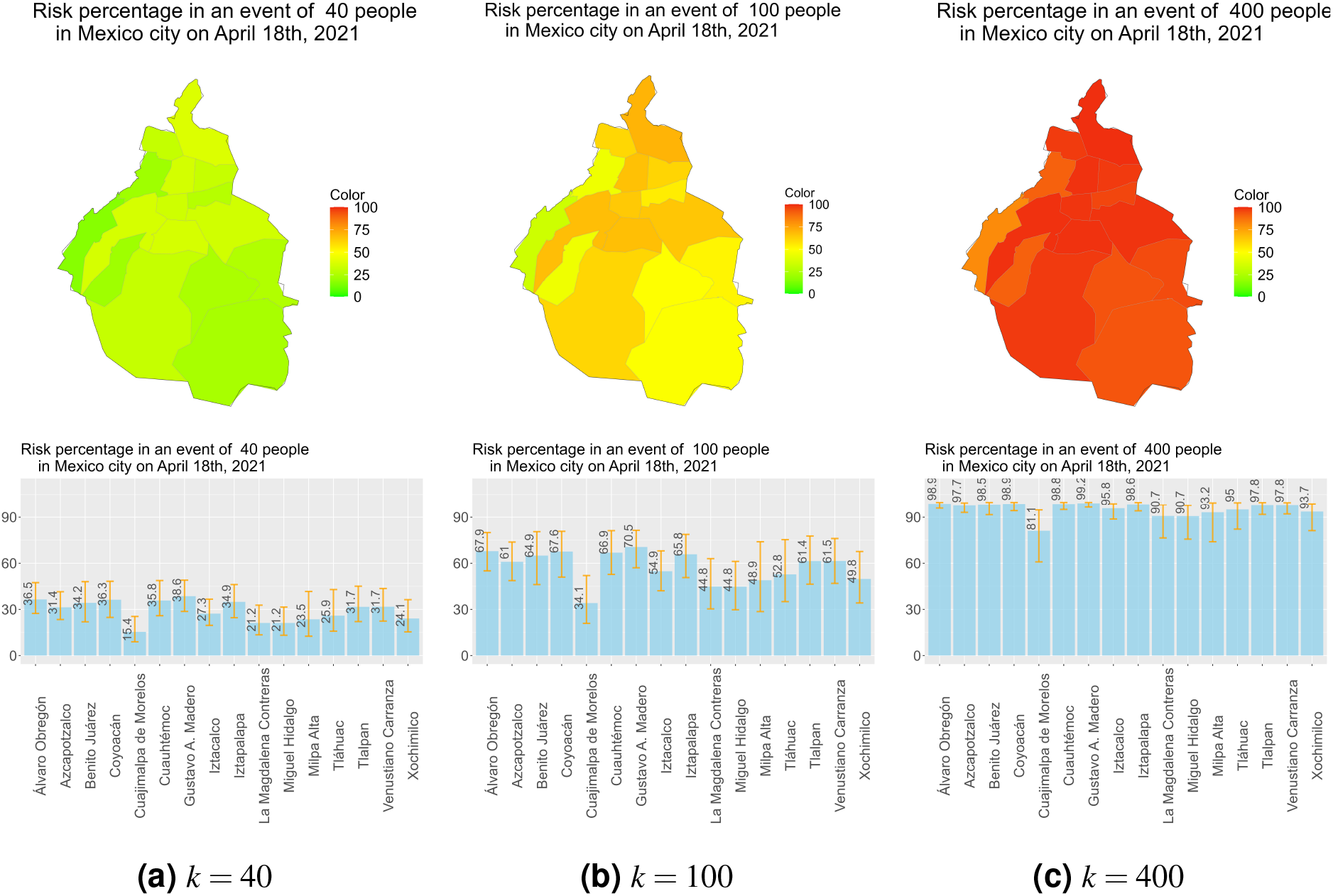
Risk estimate for gatherings size *k* = 40, 100, 400 across CDMX for April 26 to May 2, 2021.

This change in pattern reflects the nonlinear growth of *C*(*t*; *k*) as a function of *k*. Consequently, our weekly reports included risk estimates for gathering sizes *k* = 40, 100 and 400 at the level of states, municipalities in Queretaro, and boroughs of Mexico City. For example, Figures 6–8 were part of the report corresponding to the week of April 26 to May 2, 2021. These figures present estimated event risk for the specified gathering sizes, based on data updated through April 25, 2021 and truncated on April 18, 2021.

### 4.3 Temporal evolution and comparison with the official traffic-light system

In order to evaluate the agreement between the proposed risk traffic-light system and the official one, we discretized the estimated risk levels according to the previously defined categories and generated traffic-light classifications for gathering sizes ranging from 1 to 200 individuals for each state at every official reporting date between July 7, 2020, and October 4, 2021.

Agreement across all states and reporting dates was assessed using the weighted Cohen’s kappa statistic [16]. The highest agreement among the 512 traffic-light classifications was obtained for a gathering size of 86 individuals, yielding a kappa coefficient of 0.736, which suggests substantial concordance between the proposed and official risk-level classifications [16]. However, rather than selecting the value that maximized statistical agreement, we focused on gathering sizes with direct operational interpretation such as those listed in Table 3.

A gathering size of 100 individuals represents a common reference capacity for medium-size meetings. The corresponding agreement with the official traffic-light system remained substantial (*κ* = 0.712), only slightly below the maximum observed value. Therefore, we adopted an event size of 100 individuals as the primary reporting scenario because it provides a more intuitive and easily communicable benchmark while preserving a high level of concordance with the official classification.

Figure 9a presents the evolution of the estimated risk traffic-light classification for gatherings of size *k* = 100 across all states, and Figure 9b shows the corresponding official classifications.

**Figure 9.**
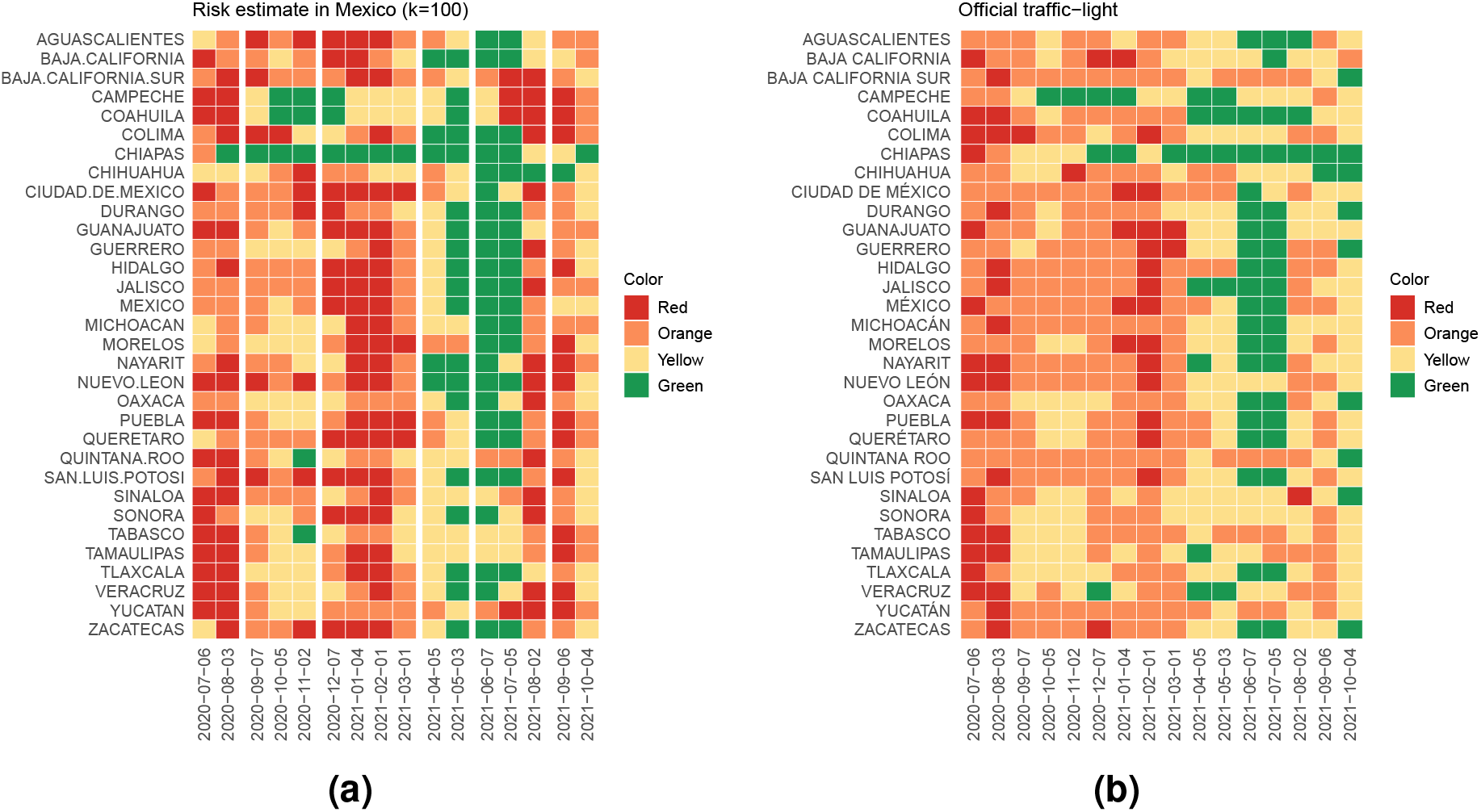
Evolution of traffic-light indicators across each state in Mexico. (a) Discretization of the risk estimate for gathering size *k* = 100 using the estimate of infections by covidestim. (b) Official traffic-light evolution.

These visualizations also provide a compact representation of epidemic waves and recovery periods, enabling rapid comparisons across localities.

Since our primary objective during the epidemic was to anticipate the official traffic-light classification for Querétaro, Figure 10 compares the evolution of reported cases with both the proposed and official traffic-light classifications for the state. The proposed classification coincided with the official one on 56% of the reporting dates, while it underestimated and overestimated the official risk level on 6% and 38% of the dates, respectively. It is important to note that the discrepancies with the official traffic-light system were by a single risk level.

**Figure 10.**
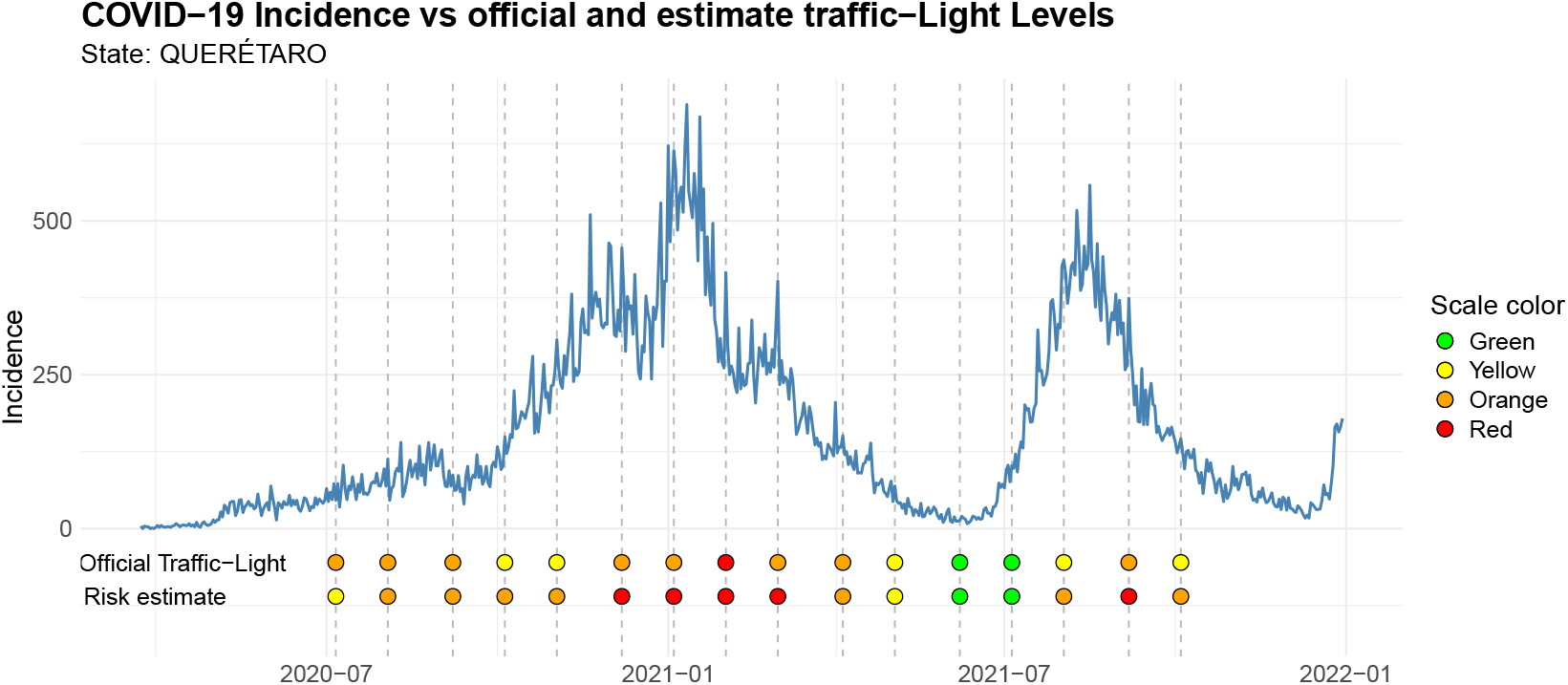
Comparison of the official traffic-light system and the discretized risk estimates for the state of Querétaro between July 6, 2020, and October 4, 2021, along with the reported COVID-19 incidence.

Furthermore, since covidestim provides estimates at the municipal level, we constructed a corresponding risk traffic-light at this spatial scale. The heatmaps in Figure 11 illustrate the evolution of risk across the boroughs of Mexico City and the municipalities of Querétaro, respectively, and compare these patterns with those obtained from state-level aggregated data, as well as with the official traffic-light color scale over time. The heatmaps from all remaining states can be found as the supplementary material.

**Figure 11.**
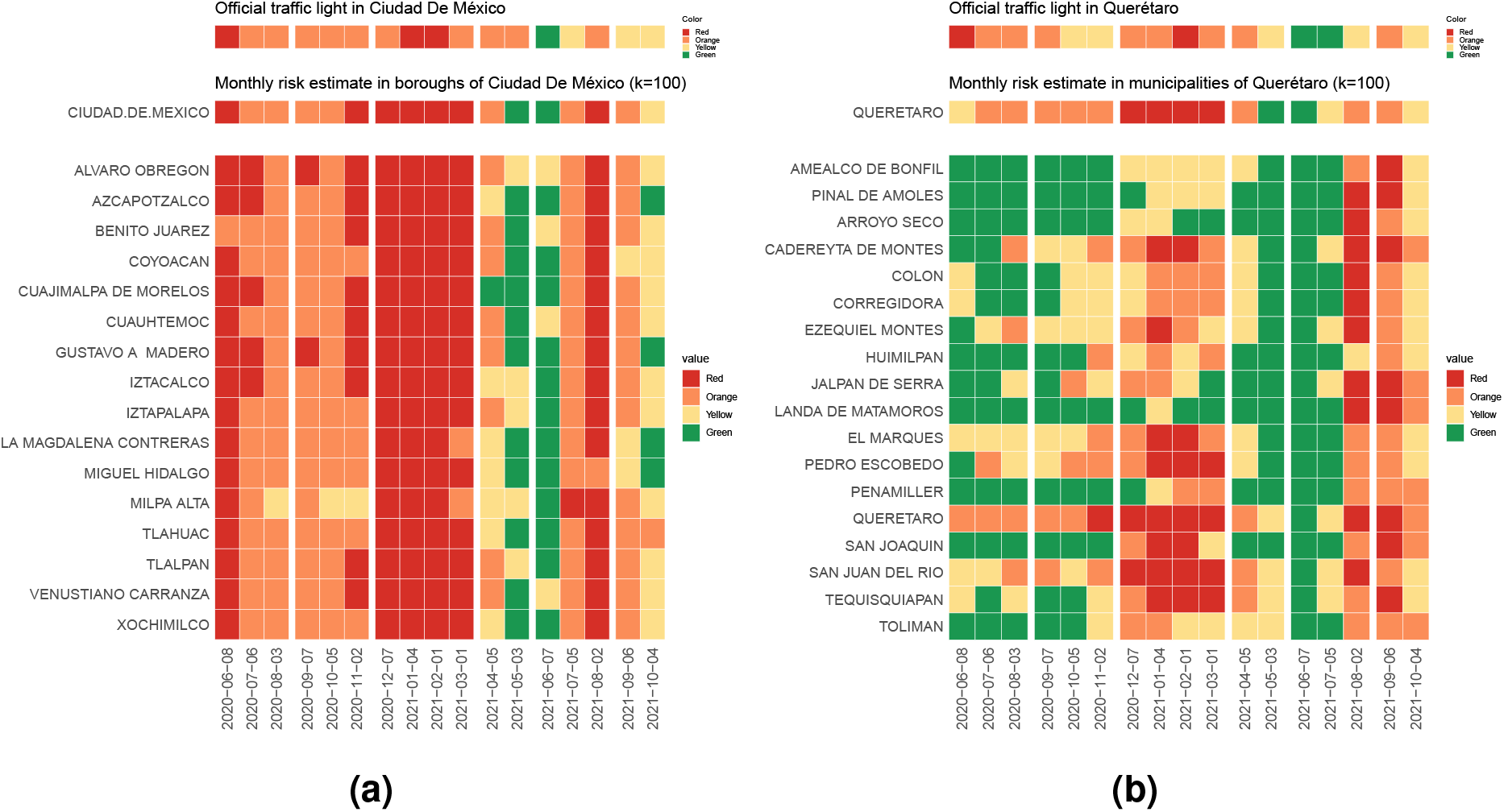
Comparison of the evolution of the official traffic-light of the state with the evolution of the discretized risk estimates for gathering size *k* = 100 in the state and its municipalities or boroughs. (a) Evolution of traffic-lights systems in Mexico City. (b) Evolution of traffic-lights in Queretaro.

In Figure 11a we observe an homogeneous behavior on the change of risk estimates across the boroughs of Mexico city, which may be due to the strong connection among them and the high population density there.

However, in Figure 11b a substantial heterogeneity across municipalities is shown, even on time.

### 4.4 Operational use in policy support

From November 2020 through October 2021, weekly reports summarizing the outputs of covidestim and the corresponding risk levels for Mexican states, boroughs of Mexico City, and municipalities of Querétaro were delivered to the Secretary of Public Education of the State of Queretaro (SEDEQ) as an early-warning tool prior to the publication of the official epidemiological traffic-light update. The aim was to support operational decision-making related to school reopening and the management of public spaces.

These weekly reports, along with a public dashboard displaying the estimates, can be consulted in the sections “Estimadores Covid-19” and “Dashboard Covid-19” on the official webpage of the the Multidisciplinary Node for Applied Mathematics (NoMMA) at the Institute of Mathematics of the National Autonomous University of Mexico[15].

### Data and resources

COVID-19 case data used in this study were obtained from the official open-data repository of the Mexican Ministry of Health [9].

The processed datasets, source code, and figures required to reproduce the analyses presented in this study are publicly available in our GitHub repository [17].

## 5 Discussion

During the pandemic, we were able to implement a municipal-level infection risk indicator based on estimates of the total number of cases from the probabilistic model covidestim. In contrast to the official epidemiological traffic-light indicator (Appendix A) used by the Mexican government during the pandemic, our methodology does not require hospital-capacity indicators (e.g., bed occupancy) but instead uses routinely reported cases and deaths only, to infer latent infections and prevalence. As described in the main text, after calibrating shaded priors distributions of Table 1 using Mexico City data, we held these priors fixed throughout the analysis period, obtaining a feasible weekly updating workflow.

Although those prior distributions for disease-progression delays and the probabilities of severe disease and death could, in principle, be estimated separately for each state, surveillance data across most regions of Mexico were affected by substantial reporting delays throughout the pandemic. Nevertheless, as described in Section 3, our country-level estimates are consistent with official ENSANUT serological results [12]. In addition, for each state, the estimated number of severe cases closely matches hospitalization data, and the estimated annual rate of severe disease is in agreement with reported values. This consistency suggests that the prior distributions calibrated using data from Mexico City capture key epidemiological features of the disease progression for each state, despite regional differences in surveillance quality and reporting practices.

It is important to emphasize that, although our initial objective was to predict the official traffic-light for Queretaro which is achieved with a gathering size of 86 individuals, our approach with 100 individuals also showed substantial agreement with the official traffic-light system. Moreover, the reduction in underestimations suggests that this gathering size may provide a more cautious assessment of epidemic risk while remaining broadly consistent with the official classification.

Our results highlight three key points. First, accounting for asymptomatic and unreported infections is critical for accurate risk estimation *C*(*t*; *k*). Given the uncertainty in the reconstruction of the true epidemic dynamics, reliance solely on reported cases leads to a substantial underestimation of event risk,as illustrated in Figure 12. Second, event risk increases nonlinearly with group size; even modest changes in underlying prevalence can shift risk from moderate to high as group size increases from small to large gatherings. Third, municipality-level heterogeneity can be substantial within a state. In contexts where policy decisions are implemented locally (e.g., school operations, municipal events, venue capacity limits), state-level aggregation may obscure low-risk municipalities while simultaneously understating risk in higher-transmission urban hubs.

**Figure 12.**
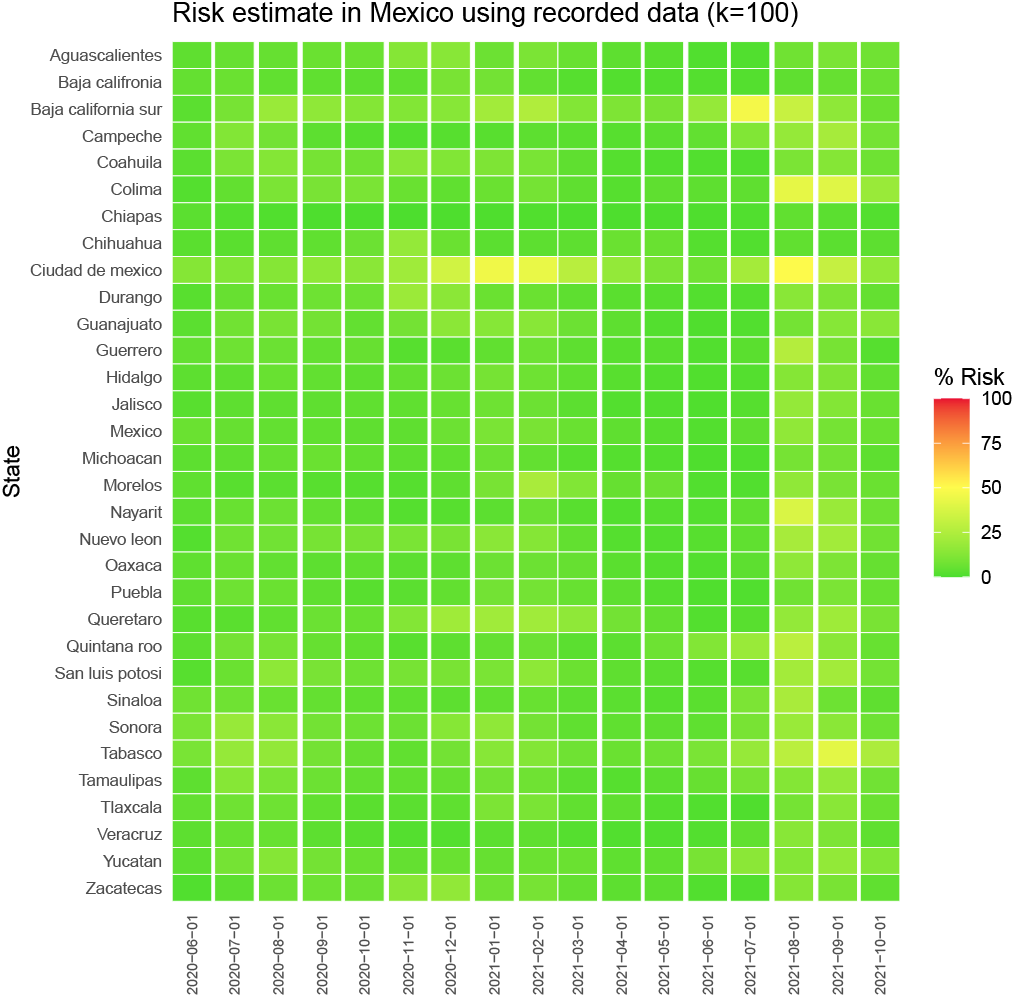
Evolution of risk estimate percentage for gathering size *k* = 100 using only reported data at state level.

Regarding our final highlight, Figure 11b suggests that both the official risk traffic-light system and the state-level risk traffic-light system with *k* = 100 for Querétaro predominantly capture the average risk dynamics of the most populous municipalities. Consequently, the overall state-level classification appears to be driven mainly by the epidemiological trends observed in the capital city and its surrounding municipalities, potentially masking heterogeneity in less populated regions. A similar pattern was observed across all Mexican states, with the exception of Mexico City, as shown in the Supplementary Material.

It is important to emphasize that, although our initial objective was to predict the risk traffic-light classification for Querétaro, the risk levels estimated using a gathering size of 100 individuals showed substantial agreement with the official traffic-light system. Moreover, the reduction in underestimations suggests that this gathering size may provide a more cautious assessment of epidemic risk while remaining broadly consistent with the official classification.

This work also highlights that access to higher-resolution data (e.g., at the postal code level) could enable more precise risk estimation; however, such granularity raises important concerns about patient data privacy and confidentiality.

Our methodology and approach have important limitations. The event-risk formulation assumes homogeneous random mixing and independent sampling of attendees, which may not hold when gatherings are structured by age, occupation, or geography. In addition, the version that we used of *covidestim* (version 2) assumes no reinfections and no vaccination; interpretation beyond mid-2021 should be cautious and ideally replaced by a framework that explicitly accounts for immunity and variant dynamics.

Despite these limitations, a municipality-resolved, probabilistically interpretable risk indicator can complement official surveillance and improve risk communication. In future work, integrating contact structure (e.g., age- or setting-specific mixing), mobility patterns, and vaccination/immune histories may yield more accurate and policy-relevant local risk estimates.

Unlike conventional epidemiological indicators, the proposed framework explicitly links infection prevalence to gathering size, allowing risk to be quantified in terms that are directly relevant to individual and policy decisions. By translating epidemic dynamics into the probability of encountering at least one infected individual in gatherings of different sizes, the indicator provides actionable information that can support more targeted and adaptive public health interventions.

## Data Availability

All data produced are available online at
https://github.com/RuthCoronaMoreno/COVID19-MX-Estimation

https://drive.google.com/drive/folders/1kDRYQZeHtBzf-QfbNa5k3_k9HFOZVmr0?usp=drive_link

## Acknowledgements

We acknowledge the technical support of Luis Aguilar, Alejandro De León, Alejandro Ávalos, and Jair García from the Laboratorio Nacional de Visualización Científica Avanzada (LAVIS, UNAM). We are also grateful to José Carlos Arredondo Velázquez, Secretary of Education of the state of Querétaro (SEDEQ) in 2021, for facilitating the operational use of the methodology described here in decision-making related to school reopening and the management of public spaces. We thank Antonio de Rosenzweig of the Instituto Mundial de Organización for his help in promoting the use of our results by the general public. We further thank the PhD. Nancy Leticia González and the students Samuel Romero, Natalia Ramírez, Santiago Espinoza and Eduardo Mendieta for their valuable assistance and contributions during the pandemic in the development of weekly reports.

## A Official epidemiological traffic-light system

The epidemiological traffic-light system is a technical tool established by the Ministry of Health of Mexico on June 1st, 2020, to define the risk of infection by state level for the reopening of social, educational, and economic activities on a weekly basis, although a month and a half later, it was changed to a biweekly update. This instrument served as the official basis for decision-making on reopening spaces or events until its last publication on April 18th, 2022, when the country had remained at the lowest color (green) for several consecutive periods [18].

The construction of the Mexican epidemiological traffic-light is based on weekly estimates of the effective reproduction number (*R*_*t*_) for COVID-19, the incidence rate of estimated active cases per 100,000 inhabitants, the mortality rate per 100,000 inhabitants, and the hospitalization rate per 100,000 inhabitants [3]. It also incorporates trends in COVID-19 syndrome cases, hospitalizations, and mortality (all standardized per 100,000 inhabitants). Additionally, the methodology considers the percentage of hospital occupancy of general beds and the positivity rate of SARS-CoV-2 PCR tests. To all indicators an score from 0 to 4 was assigned based on the range of their estimated or reported value, so that these scores were summed up to assign a color based on Table 4.

**Table 4.** Epidemic risk color according to the total score obtained from the indicators above mentioned [3].

| Color | Total score | Epidemic risk |
| --- | --- | --- |
| Red | 30 – 40 | Maximum |
| Orange | 20 – 29.5 | High |
| Yellow | 10 – 19.5 | Moderated |
| Green | 0 – 9.5 | Low |

Each color of the traffic-light correspond to some specific recommendations regarding mobility, use of face mask in public spaces and capacity of economic, social and school activities. Some examples of this activities are shown in Table 5 according to the official traffic-light methodology[3].

**Table 5.**
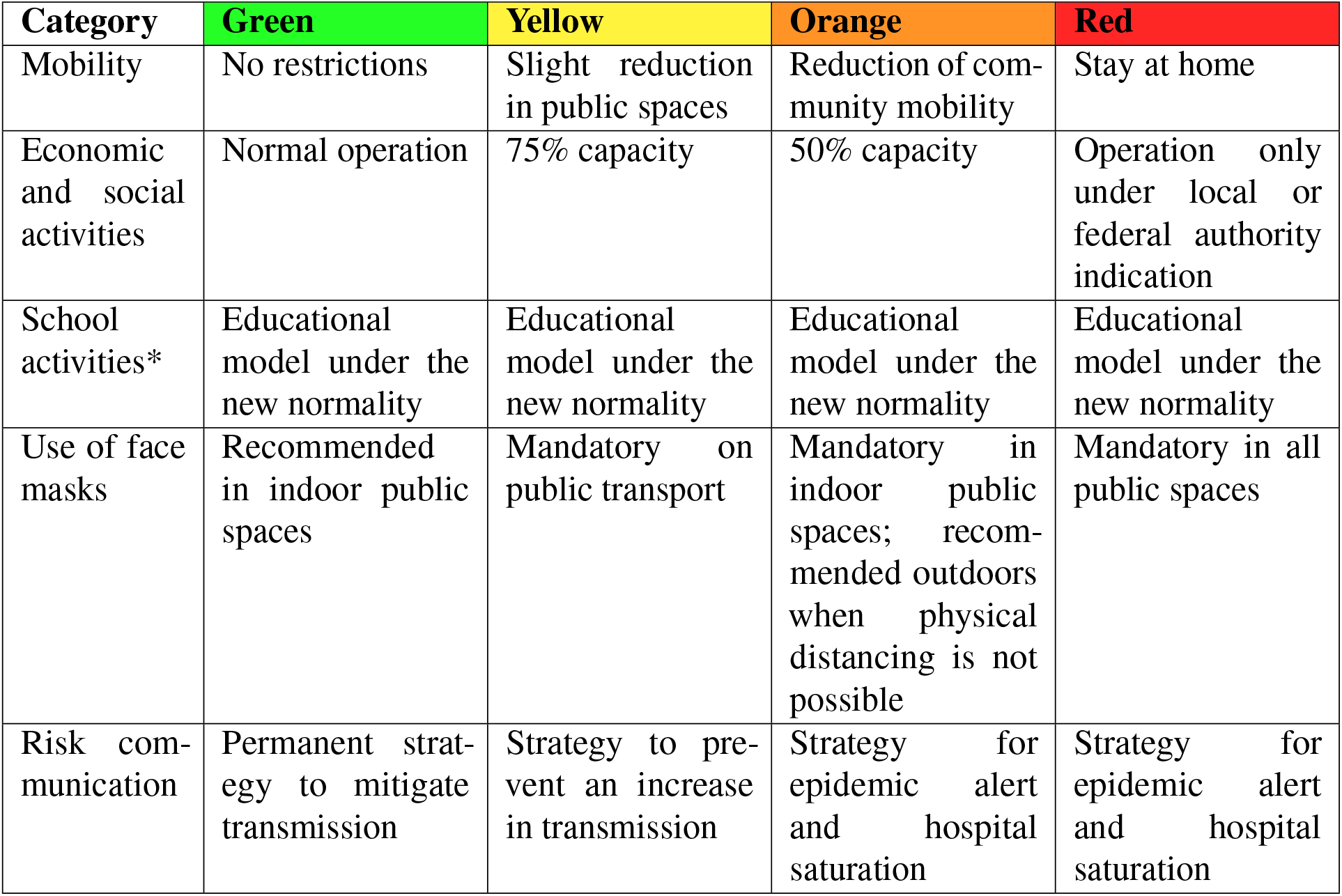
Examples of specific activities by epidemic risk level of the official epidemiological traffic-light [3]. *According to the provisions established by the Ministry of Public Education.

| Category | Green | Yellow | Orange | Red |
| --- | --- | --- | --- | --- |
| Mobility | No restrictions | Slight reduction in public spaces | Reduction of community mobility | Stay at home |
| Economic and social activities | Normal operation | 75% capacity | 50% capacity | Operation only under local or federal authority indication |
| School activities* | Educational model under the new normality | Educational model under the new normality | Educational model under the new normality | Educational model under the new normality |
| Use of face masks | Recommended in indoor public spaces | Mandatory on public transport | Mandatory in indoor public spaces; recommended outdoors when physical distancing is not possible | Mandatory in all public spaces |
| Risk communication | Permanent strategy to mitigate transmission | Strategy to prevent an increase in transmission | Strategy for epidemic alert and hospital saturation | Strategy for epidemic alert and hospital saturation |

Figure 13 presents the official weekly epidemiological traffic-light for each state in Mexico over the period from June 8, 2020, to October 25, 2021 [19].

**Figure 13.**
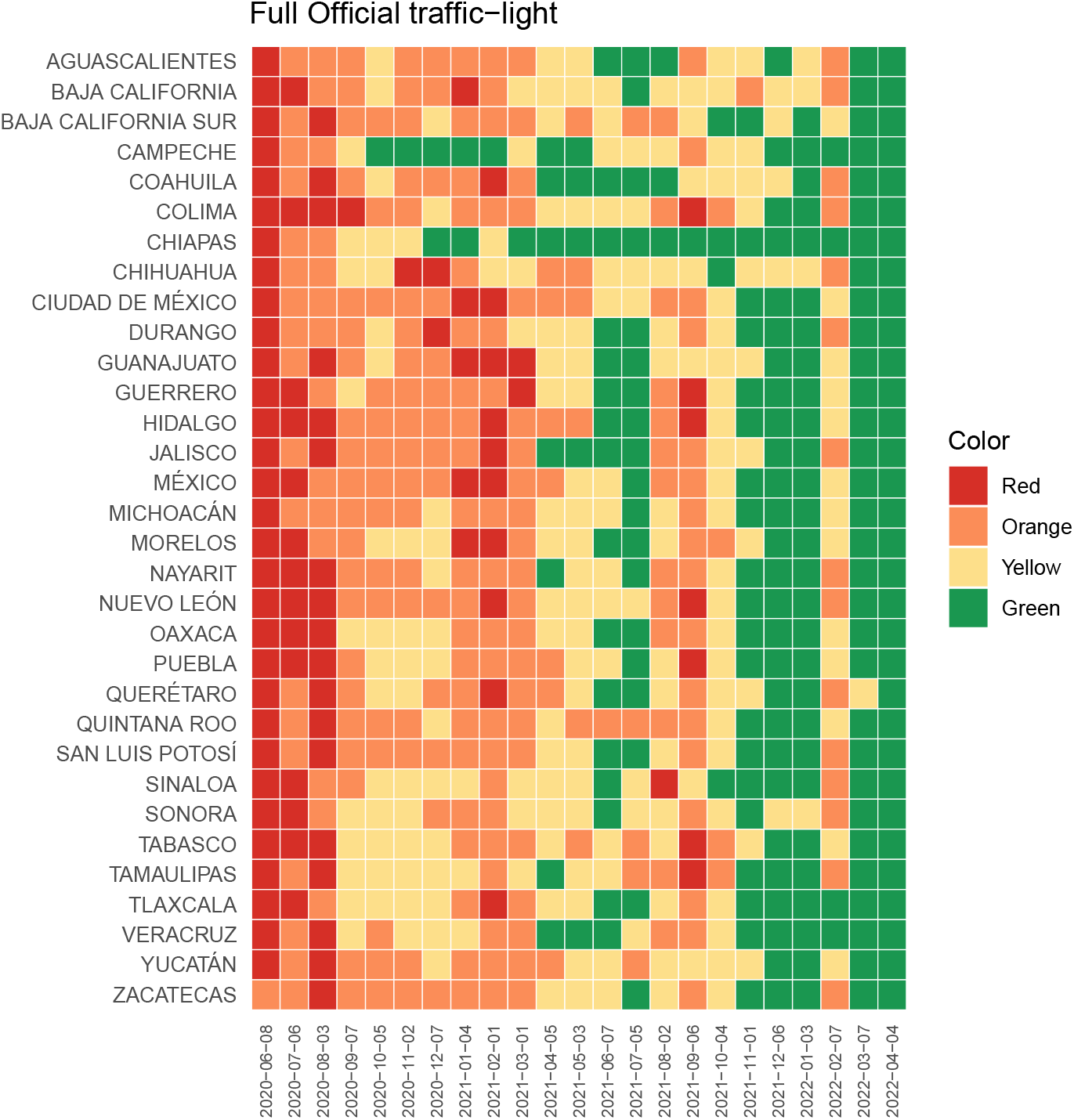
Official traffic-light

**Figure 14.**
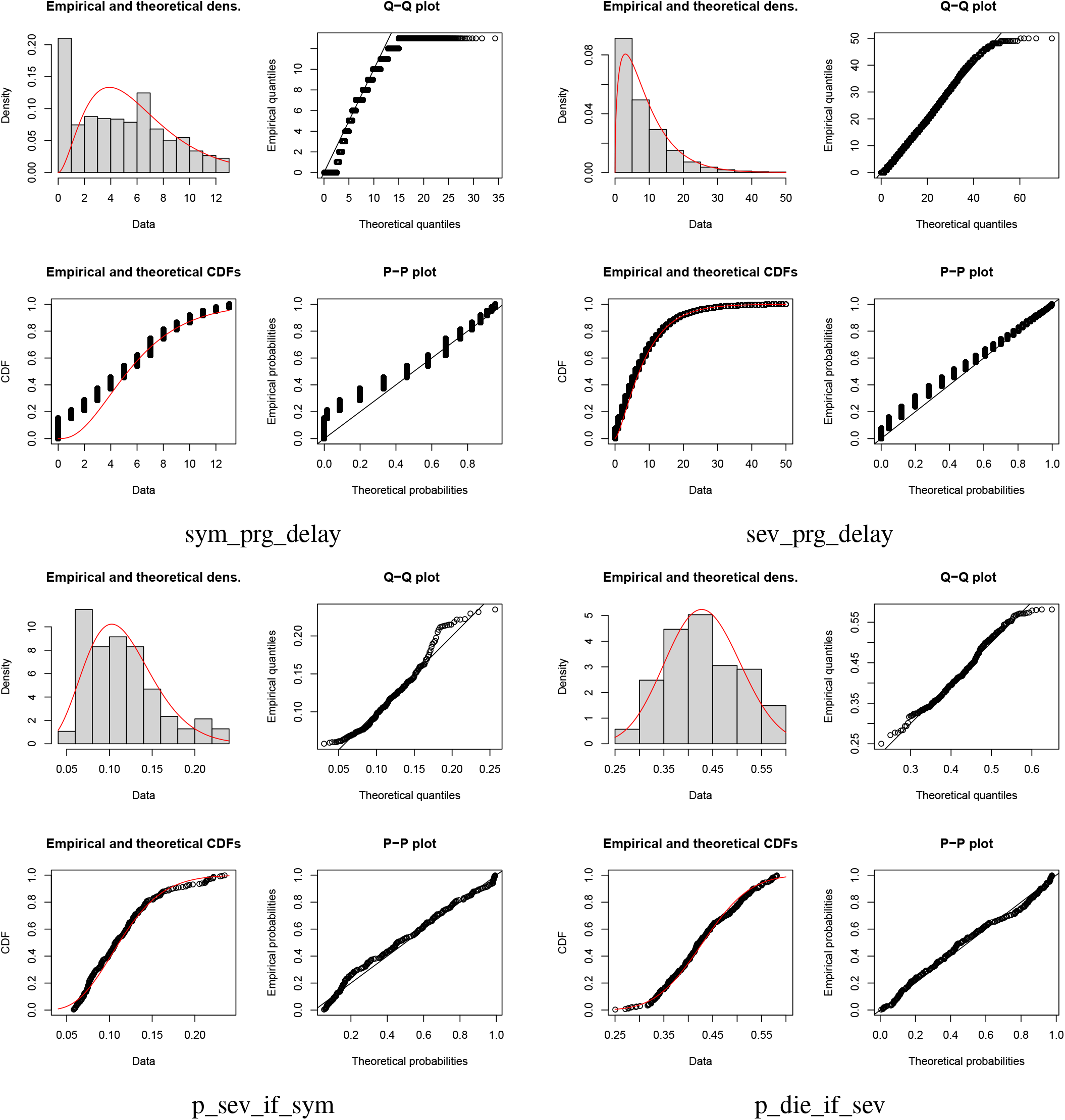
Distribution comparison plots of the estimate distributions fitted to the CDMX COVID-19 data from March 23, 2020, to January 30, 2021.

Given the guidelines for calculating the epidemiological traffic-light[3] and free access to the database of reported individual cases of COVID-19[9], we implemented the computation of the epidemiological traffic-light for the state of Queretaro. The official data[9] were used to compute the indicators except for both the hospital occupancy percentage of general and ventilator-equipped beds, which were consulted in the official webpage of *IRAG Network Information System*[20].

Following the official methodology[3], we used to download the official reported data of COVID-19 every Tuesday, in order to compute the epidemiological traffic-light for the following week. Confirmed cases were filtered from the database by the classifications 1, 2, and 3 of column “CLASIFICACION_FINAL”. In order to compute indicators related to trends and rates, cubic splines were applied to recorded data of confirmed cases, considering cuts of a few last days, because of the reporting delays, whether due to delays in patients arriving at hospitals or delays on the part of local authorities in capturing information in the Respiratory Disease Surveillance System (SISVER) and Acute Respiratory Infections (IRAG) networks.

Given that the data are recorded by date of symptom onset (FECHA_SINTOMAS), the last 14 days were excluded due to the reporting and symptoms onset delay. This data was used to compute the indicators “Trend of COVID-19 syndrome cases per 100,000 inhabitants” and “the effective reproduction number (*R*_*t*_) of COVID-19”. To compute this last indicator, the *Epiestim software* [13, 21], developed in the statistical program R, was used, with a 7-day moving average.

On the other hand, to compute the indicators “Hospitalization rate per 100,000 inhabitants” and “Trend in hospitalizations per 100,000 inhabitants”, hospitalized cases were recorded by the date of registration in the system (FECHA_INGRESO), up to 7 days before the date of database download, due to the reporting delay.

Regarding the indicators “Mortality rate per 100,000 inhabitants” and “Trend of mortality rate per 100,000 inhabitants, deaths were recorded by date of death (FECHA_DEF) up to 3 days before the date of database download, because of the reporting delays.

The proportion of suspected cases determined by the positivity rate has been added to the incidence in order to compute the “Incidence rate of estimated active cases per 100,000 inhabitants”.

Finally, the “SARS-CoV-2 PCR positivity percentage (USMER)” was calculated as the ratio of “the total of positive tests” to “the number of total tests” during the week before the last epidemiological week. Individuals who were tested are identified in the database by the value 1 in the TOMA_MUESTRA_LAB column. Of these, those who are confirmed by testing are identified by the value 1 in the column called RESULTADO_LAB.

## B Distribution fitting diagnostics

This appendix presents the distribution comparison plots for the estimated distributions fitted to the CDMX COVID19 data shown in Table 2.

For each estimated distribution, we display the empirical and theoretical densities, the empirical and theoretical cumulative distribution functions (CDFs), as well as the Q–Q and P–P plots. These graphical diagnostics were used to conduct a visual assessment of the goodness of fit of each distribution.

