## Supplemental material for "A risk-of-contagion index using a Bayesian based model for the COVID-19 epidemic in Mexico"

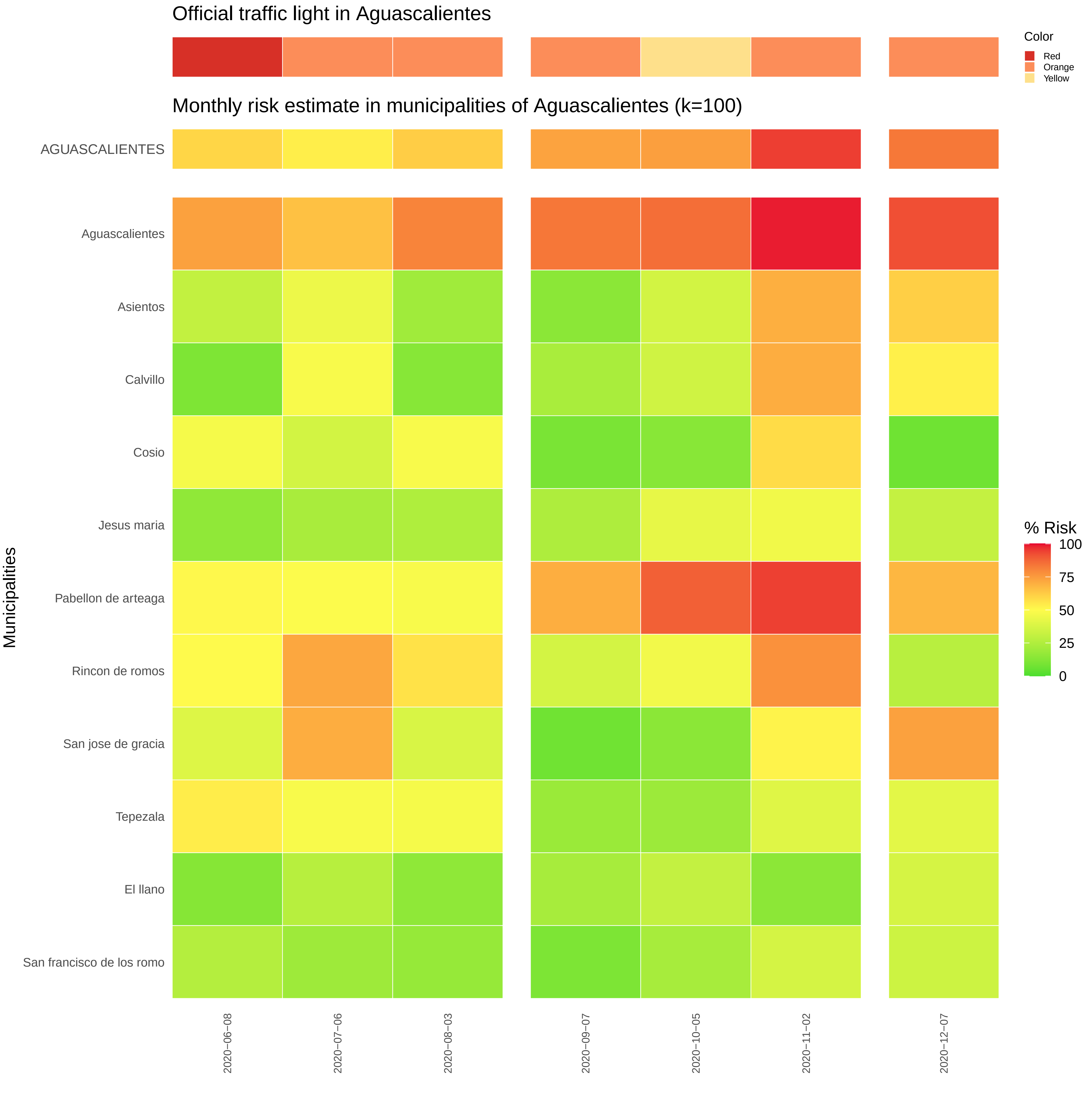

Official traffic light in Aguascalientes
Color
Red
Orange
Yellow
Monthly risk estimate in municipalities of Aguascalientes (k=100)
AGUASCALIENTES
Aguascalientes
Asientos
Calvillo
Cosio
Jesus maria
Pabellon de arteaga
Rincon de romos
San jose de gracia
Tepezala
El llano
San francisco de los romo
Municipalities
% Risk
100
75
50
25
0
2020−06−08
2020−07−06
2020−08−03
2020−09−07
2020−10−05
2020−11−02
2020−12−07

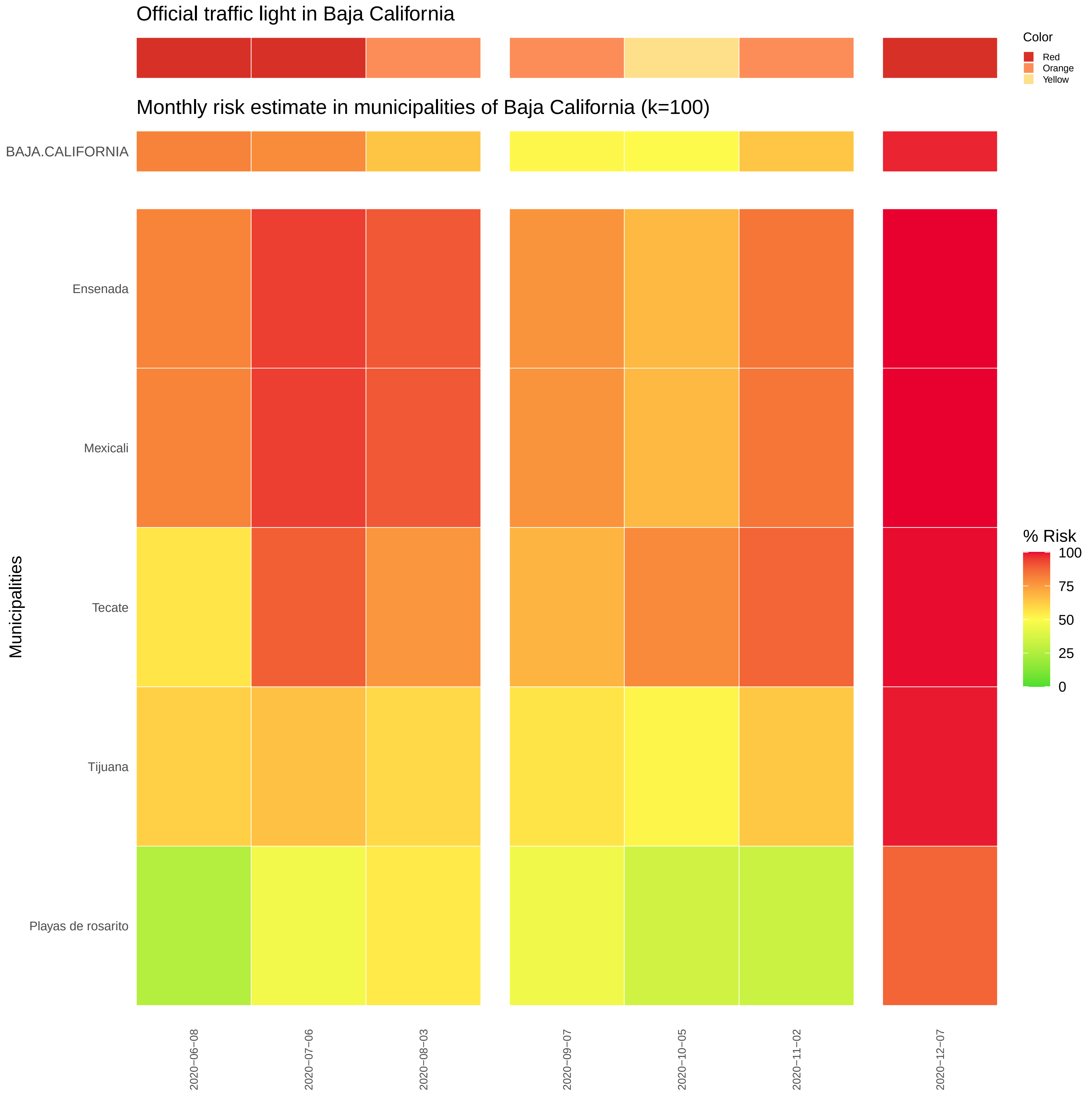

Official traffic light in Baja California
Monthly risk estimate in municipalities of Baja California (k=100)
BAJA.CALIFORNIA
Ensenada
Mexicali
Tecate
Tijuana
Playas de rosarito
Municipalities
2020−06−08
2020−07−06
2020−08−03
2020−09−07
2020−10−05
2020−11−02
2020−12−07
Color
Red
Orange
Yellow
% Risk
100
75
50
25
0

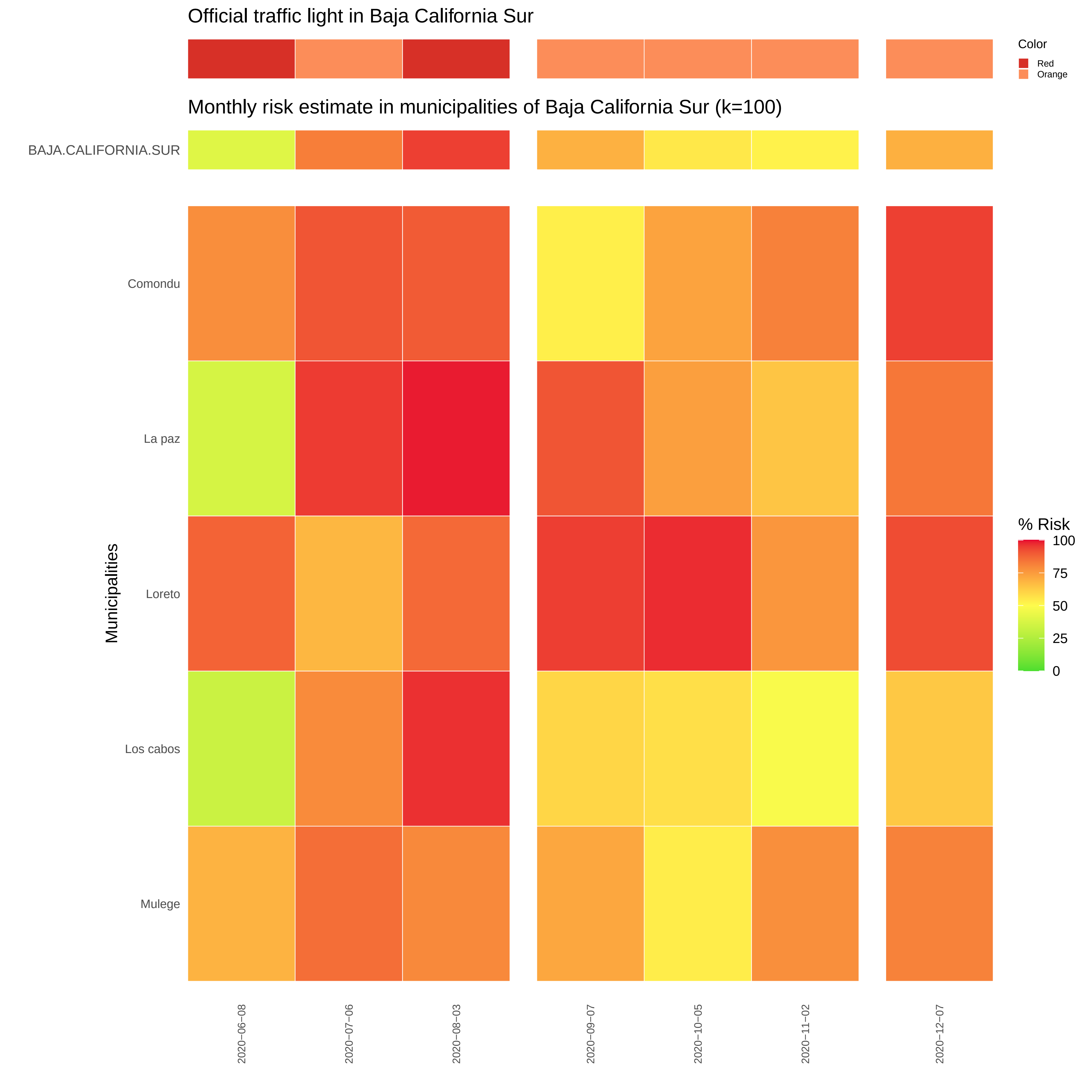

Official traffic light in Baja California Sur
Color
Red
Orange
Monthly risk estimate in municipalities of Baja California Sur (k=100)
BAJA.CALIFORNIA.SUR
Comondu
La paz
Loreto
Los cabos
Mulege
Municipalities
% Risk
100
75
50
25
0
2020−06−08
2020−07−06
2020−08−03
2020−09−07
2020−10−05
2020−11−02
2020−12−07

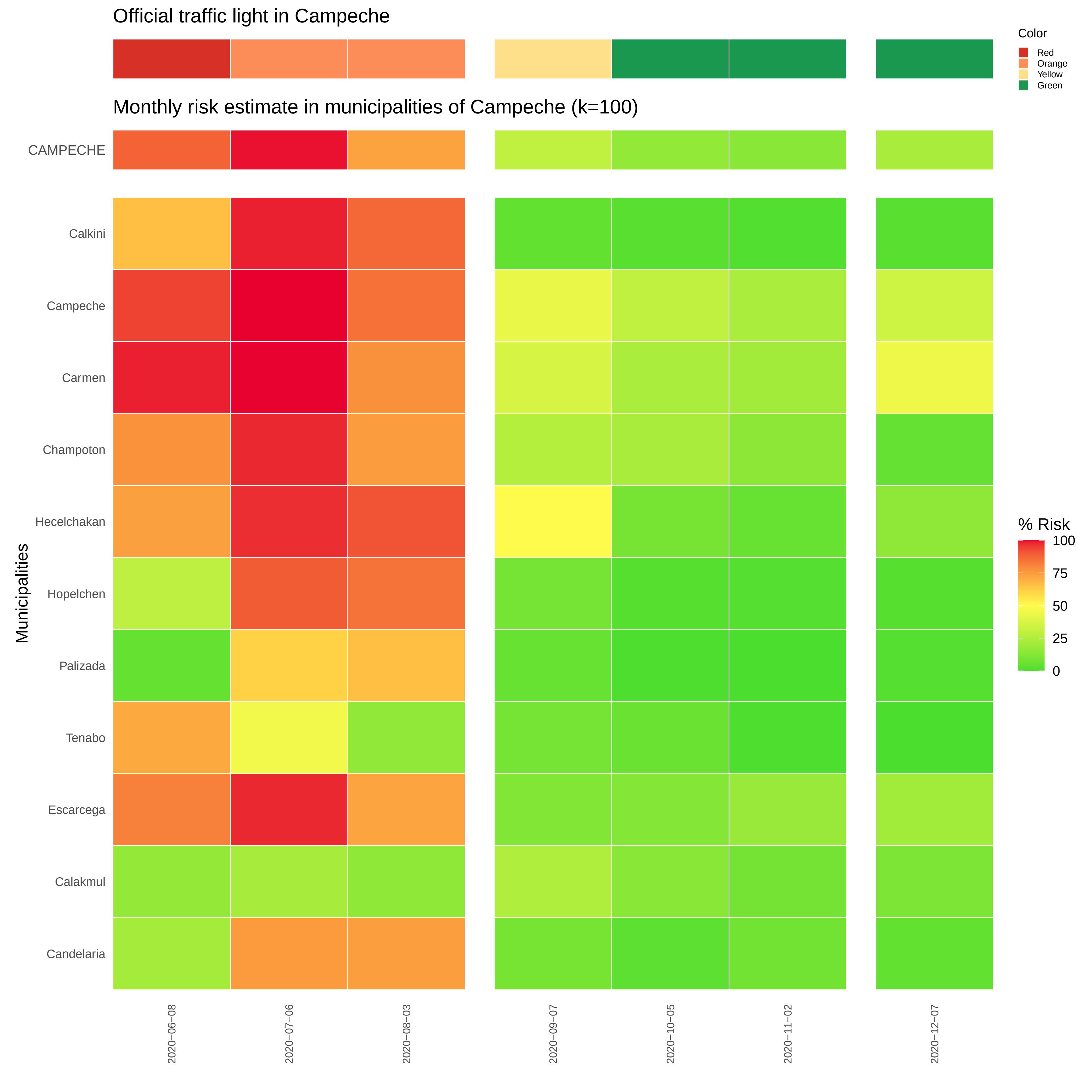

Official traffic light in Campeche
Color
Red
Orange
Yellow
Green
Monthly risk estimate in municipalities of Campeche (k=100)
CAMPECHE
Calkini
Campeche
Carmen
Champoton
Hecelchakan
Hopelchen
Palizada
Tenabo
Escarcega
Calakmul
Candelaria
Municipalities
% Risk
100
75
50
25
0
2020−06−08
2020−07−06
2020−08−03
2020−09−07
2020−10−05
2020−11−02
2020−12−07

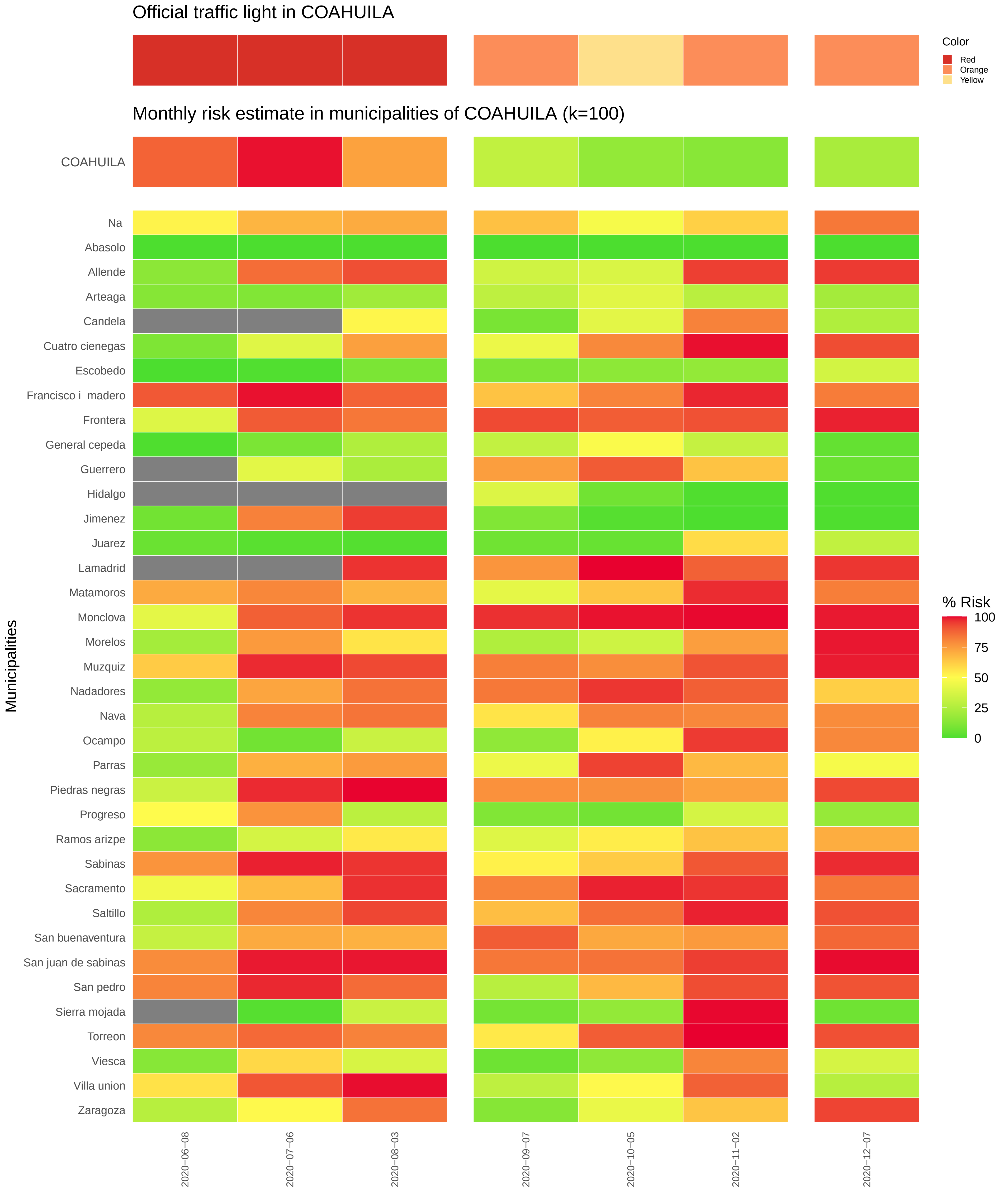
Official traffic light in COAHUILA
Color
Red
Orange
Yellow
Monthly risk estimate in municipalities of COAHUILA (k=100)
COAHUILA
Na
Abasolo
Allende
Arteaga
Candela
Cuatro cienegas
Escobedo
Francisco i madero
Frontera
General cepeda
Guerrero
Hidalgo
Jimenez
Juarez
Lamadrid
Matamoros
Monclova
Morelos
Muzquiz
Nadadores
Nava
Ocampo
Parras
Piedras negras
Progreso
Ramos arizpe
Sabinas
Sacramento
Saltillo
San buenaventura
San juan de sabinas
San pedro
Sierra mojada
Torreon
Viesca
Villa union
Zaragoza
Municipalities
% Risk
100
75
50
25
0
2020−06−08
2020−07−06
2020−08−03
2020−09−07
2020−10−05
2020−11−02
2020−12−07

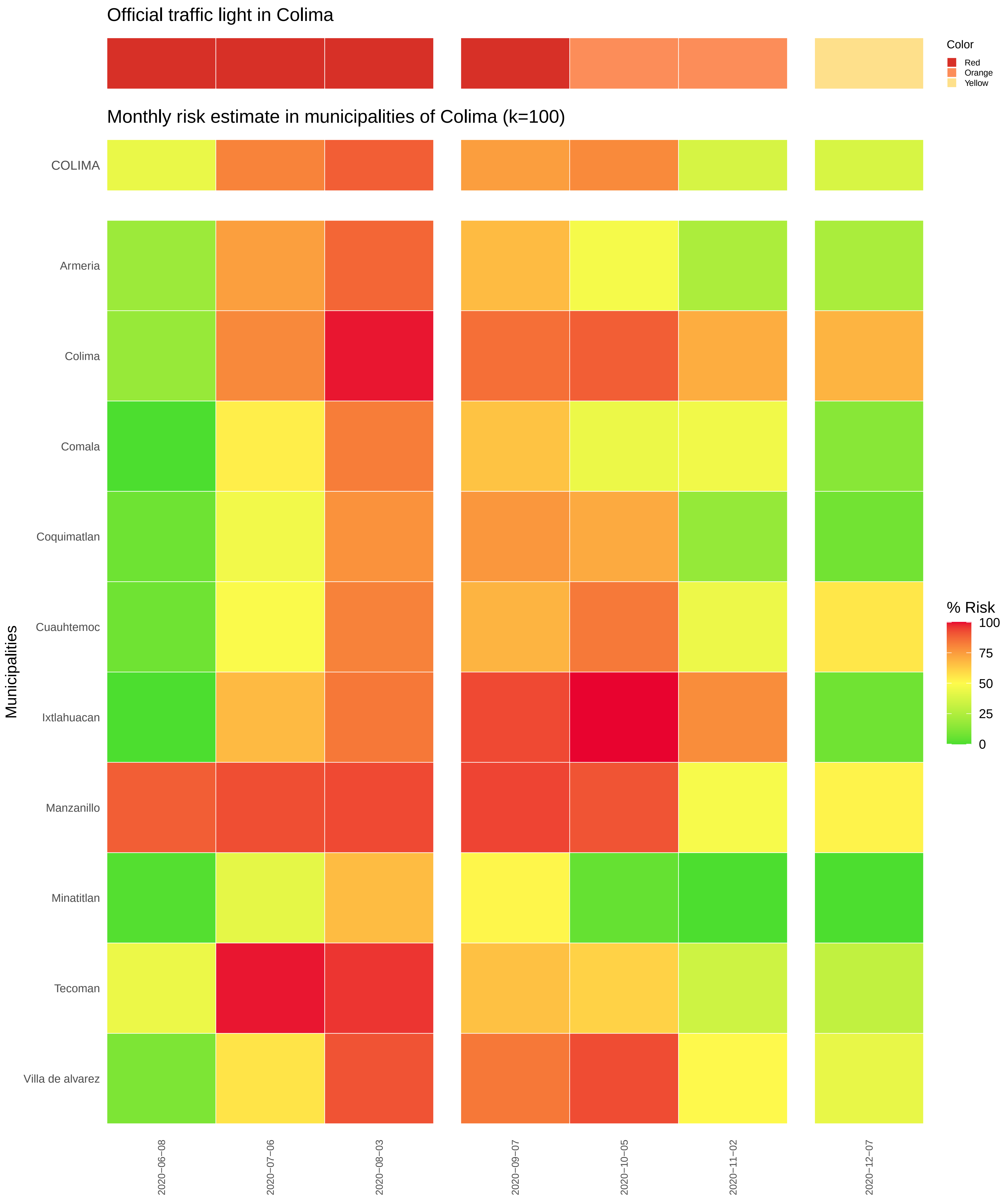
Official traffic light in Colima
Color
Red
Orange
Yellow
Monthly risk estimate in municipalities of Colima (k=100)
COLIMA
Armeria
Colima
Comala
Coquimatlan
Cuauhtemoc
Ixtlahuacan
Manzanillo
Minatitlan
Tecoman
Villa de alvarez
Municipalities
% Risk
100
75
50
25
0
2020−06−08
2020−07−06
2020−08−03
2020−09−07
2020−10−05
2020−11−02
2020−12−07

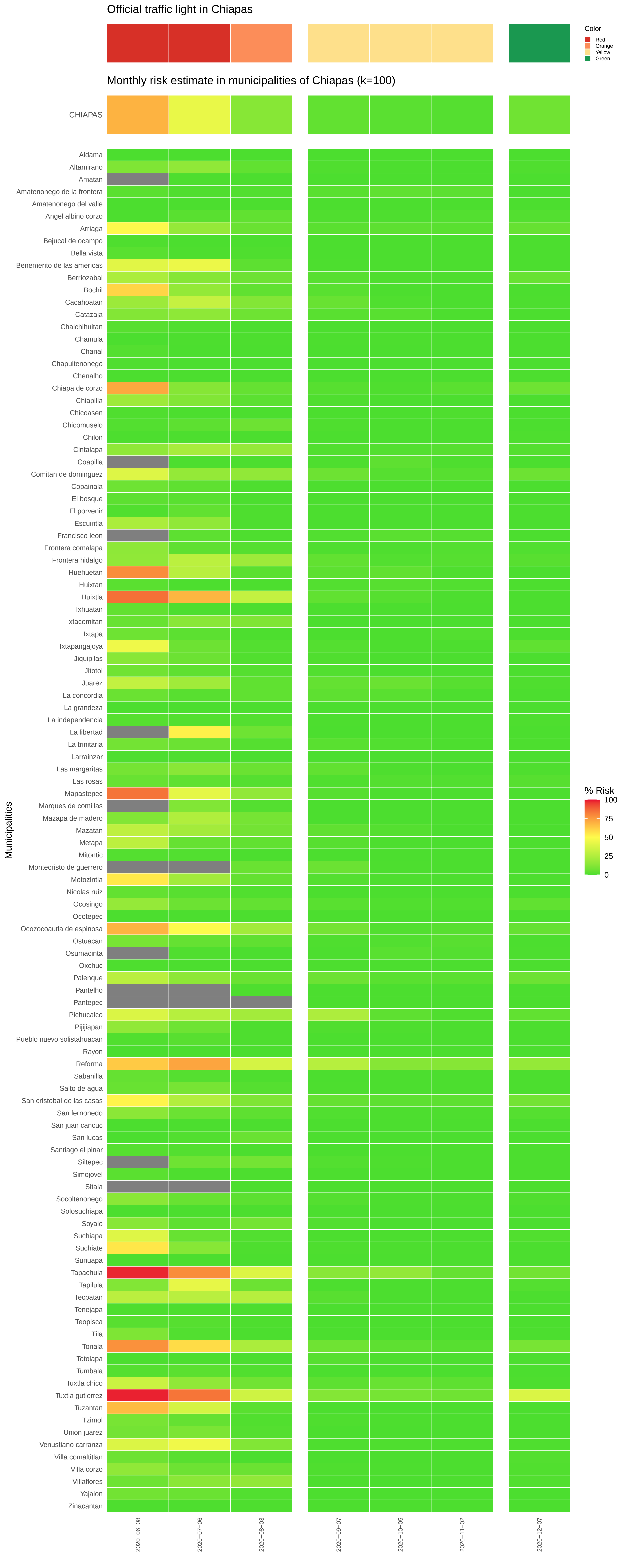
Official traffic light in Chiapas
Color
Red
Orange
Yellow
Green
Monthly risk estimate in municipalities of Chiapas (k=100)
CHIAPAS
% Risk
100
75
50
25
0
Municipalities
Aldama
Altamirano
Amatan
Amatenonego de la frontera
Amatenonego del valle
Angel albino corzo
Arriaga
Bejucal de ocampo
Bella vista
Benemerito de las americas
Berriozabal
Bochil
Cacahoatan
Catazaja
Chalchihuitan
Chamula
Chanal
Chapultenonego
Chenalho
Chiapa de corzo
Chiapilla
Chicoasen
Chicomuselo
Chilon
Cintalapa
Coapilla
Comitan de dominguez
Copainala
El bosque
El porvenir
Escuintla
Francisco leon
Frontera comalapa
Frontera hidalgo
Huehuetan
Huixtan
Huixtla
Ixhuatan
Ixtacomitan
Ixtapa
Ixtapangajoya
Jiquipilas
Jitotol
Juarez
La concordia
La grandeza
La independencia
La libertad
La trinitaria
Larrainzar
Las margaritas
Las rosas
Mapastepec
Marques de comillas
Mazapa de madero
Mazatan
Metapa
Mitontic
Montecristo de guerrero
Motozintla
Nicolas ruiz
Ocosingo
Ocotepec
Ocozocoautla de espinosa
Ostuacan
Osumacinta
Oxchuc
Palenque
Pantelho
Pantepec
Pichucalco
Pijijiapan
Pueblo nuevo solistahuacan
Rayon
Reforma
Sabanilla
Salto de agua
San cristobal de las casas
San fernonedo
San juan cancuc
San lucas
Santiago el pinar
Siltepec
Simojovel
Sitala
Socoltenonego
Solosuchiapa
Soyalo
Suchiapa
Suchiate
Sunuapa
Tapachula
Tapilula
Tecpatan
Tenejapa
Teopisca
Tila
Tonala
Totolapa
Tumbala
Tuxtla chico
Tuxtla gutierrez
Tuzantan
Tzimol
Union juarez
Venustiano carranza
Villa comaltitlan
Villa corzo
Villaflores
Yajalon
Zinacantan
2020−06−08
2020−07−06
2020−08−03
2020−09−07
2020−10−05
2020−11−02
2020−12−07

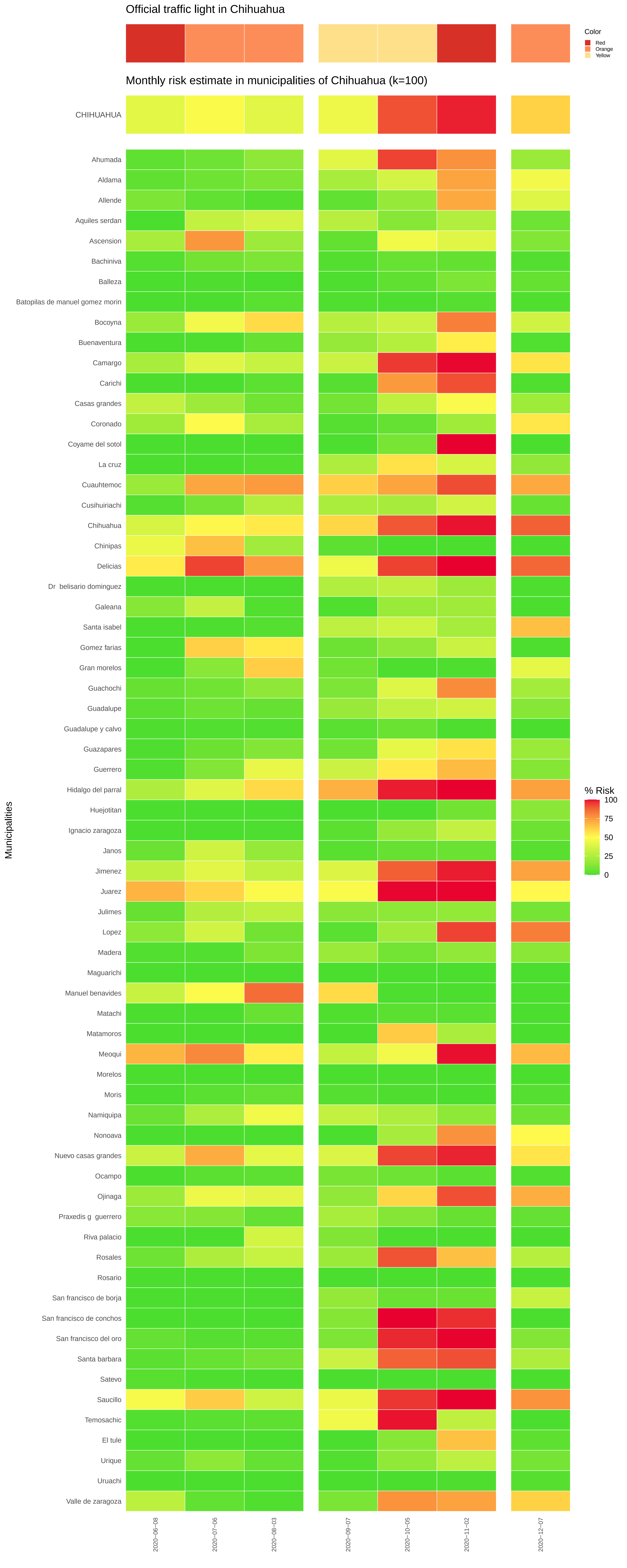
Official traffic light in Chihuahua
Color
Red
Orange
Yellow
Monthly risk estimate in municipalities of Chihuahua (k=100)
CHIHUAHUA
Ahumada
Aldama
Allende
Aquiles serdan
Ascension
Bachiniva
Balleza
Batopilas de manuel gomez morin
Bocoyna
Buenaventura
Camargo
Carichi
Casas grandes
Coronado
Coyame del sotol
La cruz
Cuauhtemoc
Cusihuiriachi
Chihuahua
Chinipas
Delicias
Dr belisario dominguez
Galeana
Santa isabel
Gomez farias
Gran morelos
Guachochi
Guadalupe
Guadalupe y calvo
Guazapares
Guerrero
Hidalgo del parral
Huejotitan
Ignacio zaragoza
Janos
Jimenez
Juarez
Julimes
Lopez
Madera
Maguarichi
Manuel benavides
Matachi
Matamoros
Meoqui
Morelos
Moris
Namiquipa
Nonoava
Nuevo casas grandes
Ocampo
Ojinaga
Praxedis g guerrero
Riva palacio
Rosales
Rosario
San francisco de borja
San francisco de conchos
San francisco del oro
Santa barbara
Satevo
Saucillo
Temosachic
El tule
Urique
Uruachi
Valle de zaragoza
Municipalities
% Risk
100
75
50
25
0
2020−06−08
2020−07−06
2020−08−03
2020−09−07
2020−10−05
2020−11−02
2020−12−07

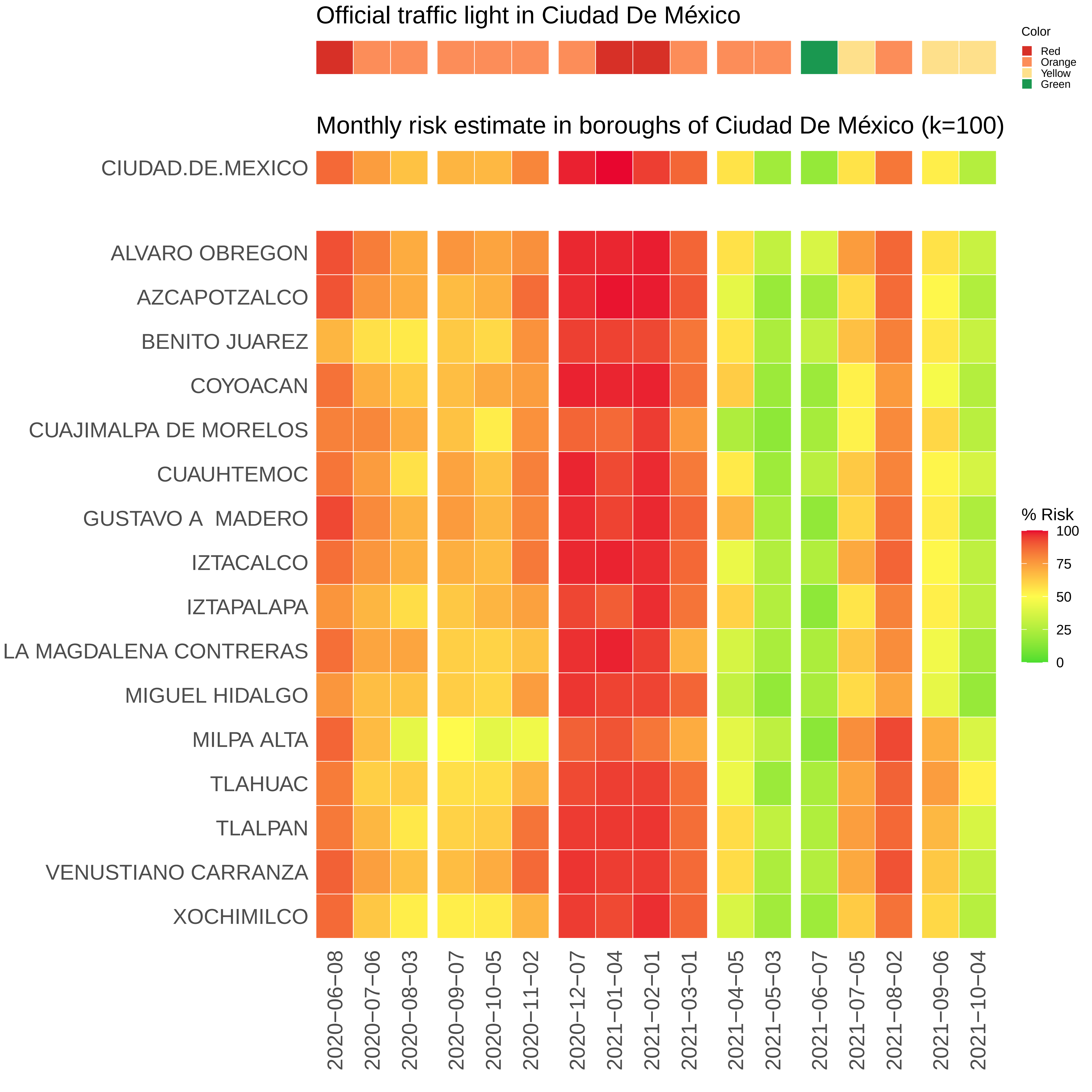

Official traffic light in Ciudad De México
Color
Red
Orange
Yellow
Green
Monthly risk estimate in boroughs of Ciudad De México (k=100)
CIUDAD.DE.MEXICO
ALVARO OBREGON
AZCAPOTZALCO
BENITO JUAREZ
COYOACAN
CUAJIMALPA DE MORELOS
CUAUHTEMOC
GUSTAVO A MADERO
IZTACALCO
IZTAPALAPA
LA MAGDALENA CONTRERAS
MIGUEL HIDALGO
MILPA ALTA
TLAHUAC
TLALPAN
VENUSTIANO CARRANZA
XOCHIMILCO
% Risk
100
75
50
25
0
2020−06−08
2020−07−06
2020−08−03
2020−09−07
2020−10−05
2020−11−02
2020−12−07
2021−01−04
2021−02−01
2021−03−01
2021−04−05
2021−05−03
2021−06−07
2021−07−05
2021−08−02
2021−09−06
2021−10−04

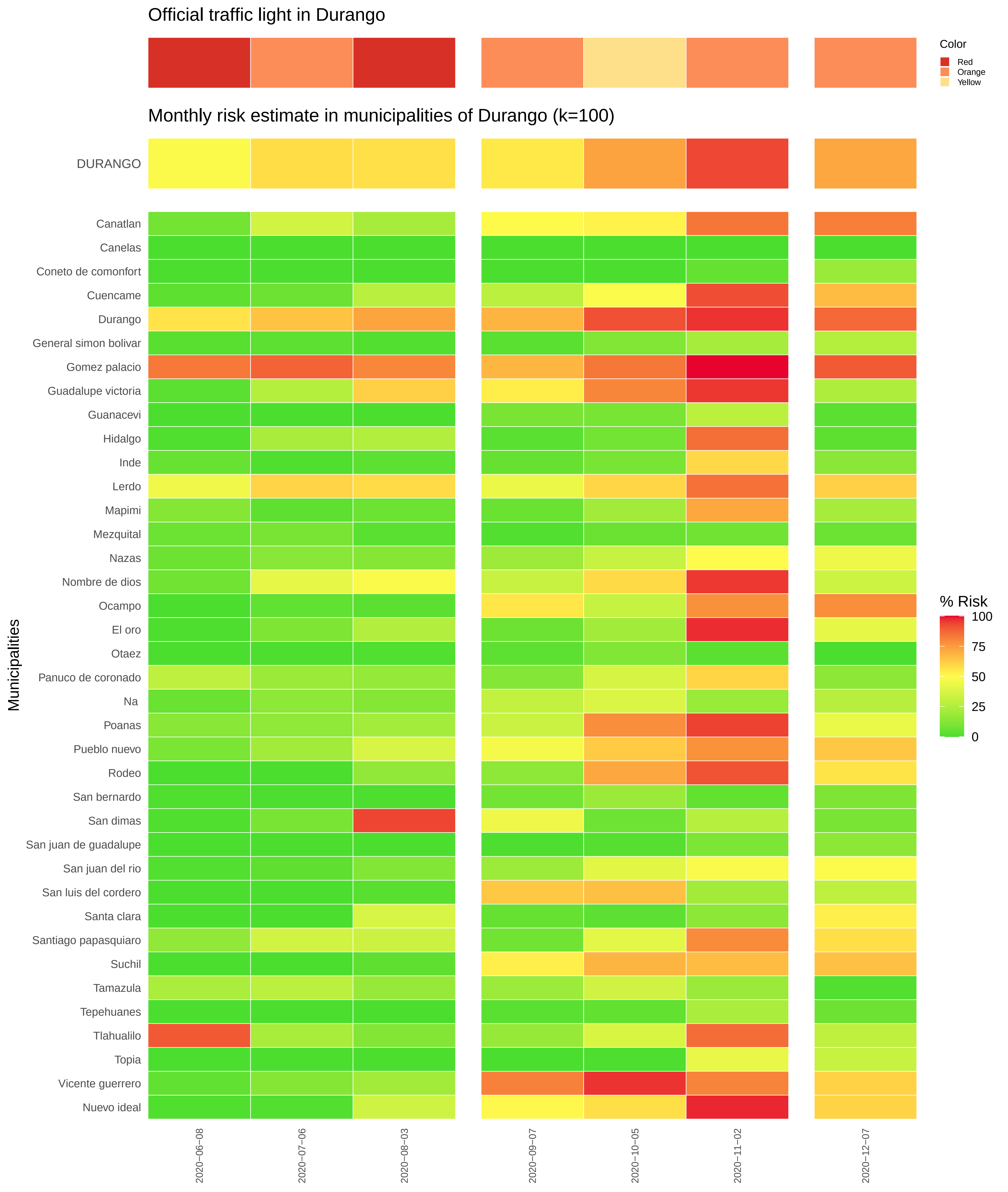

Official traffic light in Durango
Color
Red
Orange
Yellow
Monthly risk estimate in municipalities of Durango (k=100)
DURANGO
Canatlan
Canelas
Coneto de comonfort
Cuencame
Durango
General simon bolivar
Gomez palacio
Guadalupe victoria
Guanacevi
Hidalgo
Inde
Lerdo
Mapimi
Mezquital
Nazas
Nombre de dios
Ocampo
El oro
Otaez
Panuco de coronado
Na
Poanas
Pueblo nuevo
Rodeo
San bernardo
San dimas
San juan de guadalupe
San juan del rio
San luis del cordero
Santa clara
Santiago papasquiaro
Suchil
Tamazula
Tepehuanes
Tlahualilo
Topia
Vicente guerrero
Nuevo ideal
Municipalities
% Risk
100
75
50
25
0
2020−06−08
2020−07−06
2020−08−03
2020−09−07
2020−10−05
2020−11−02
2020−12−07

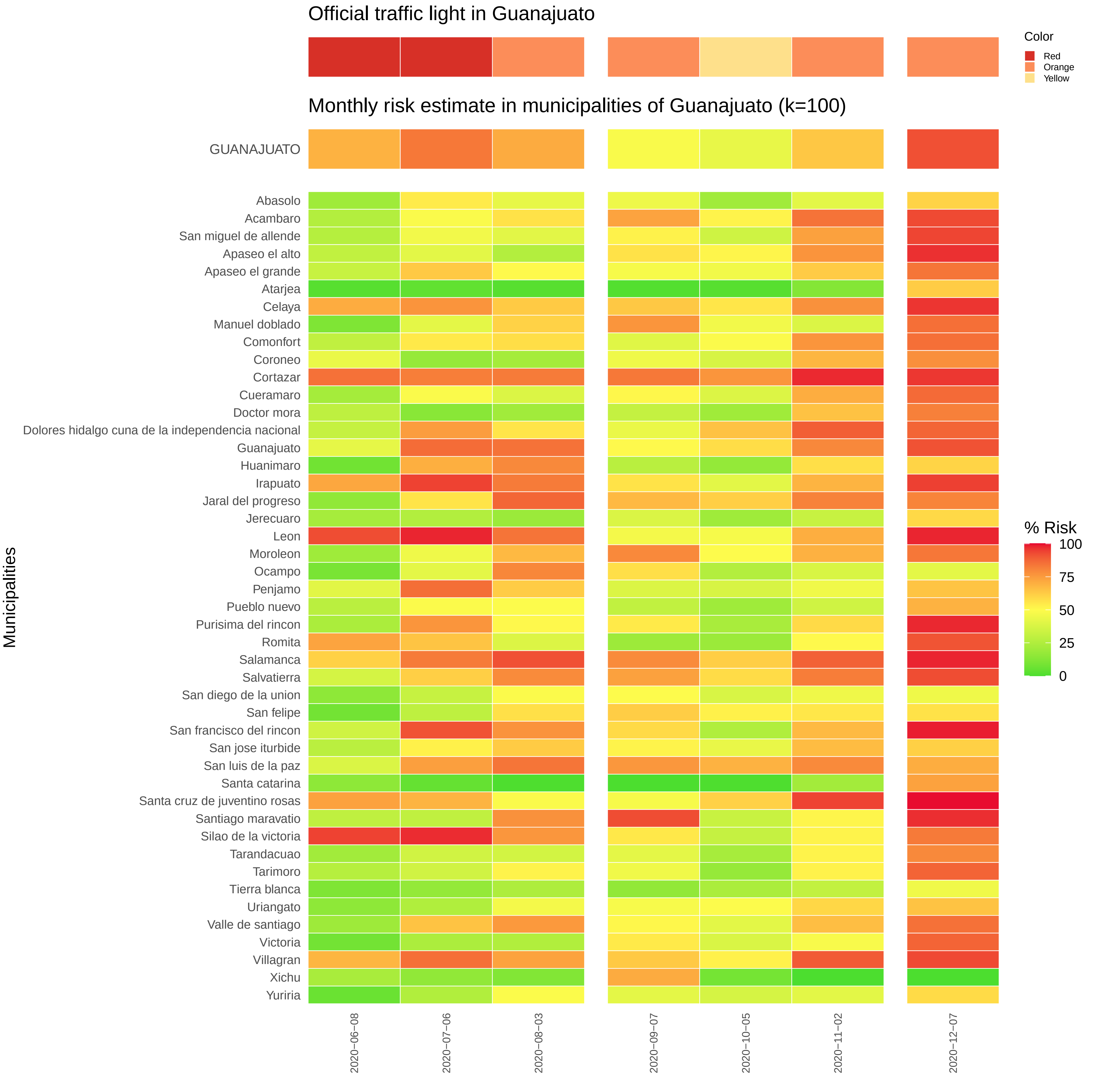

Official traffic light in Guanajuato
Color
Red
Orange
Yellow
Monthly risk estimate in municipalities of Guanajuato (k=100)
GUANAJUATO
Municipalities
Abasolo
Acambaro
San miguel de allende
Apaseo el alto
Apaseo el grande
Atarjea
Celaya
Manuel doblado
Comonfort
Coroneo
Cortazar
Cueramaro
Doctor mora
Dolores hidalgo cuna de la independencia nacional
Guanajuato
Huanimaro
Irapuato
Jaral del progreso
Jerecuaro
Leon
Moroleon
Ocampo
Penjamo
Pueblo nuevo
Purisima del rincon
Romita
Salamanca
Salvatierra
San diego de la union
San felipe
San francisco del rincon
San jose iturbide
San luis de la paz
Santa catarina
Santa cruz de juventino rosas
Santiago maravatio
Silao de la victoria
Tarandacuao
Tarimoro
Tierra blanca
Uriangato
Valle de santiago
Victoria
Villagran
Xichu
Yuriria
2020−06−08
2020−07−06
2020−08−03
2020−09−07
2020−10−05
2020−11−02
2020−12−07
% Risk
100
75
50
25
0

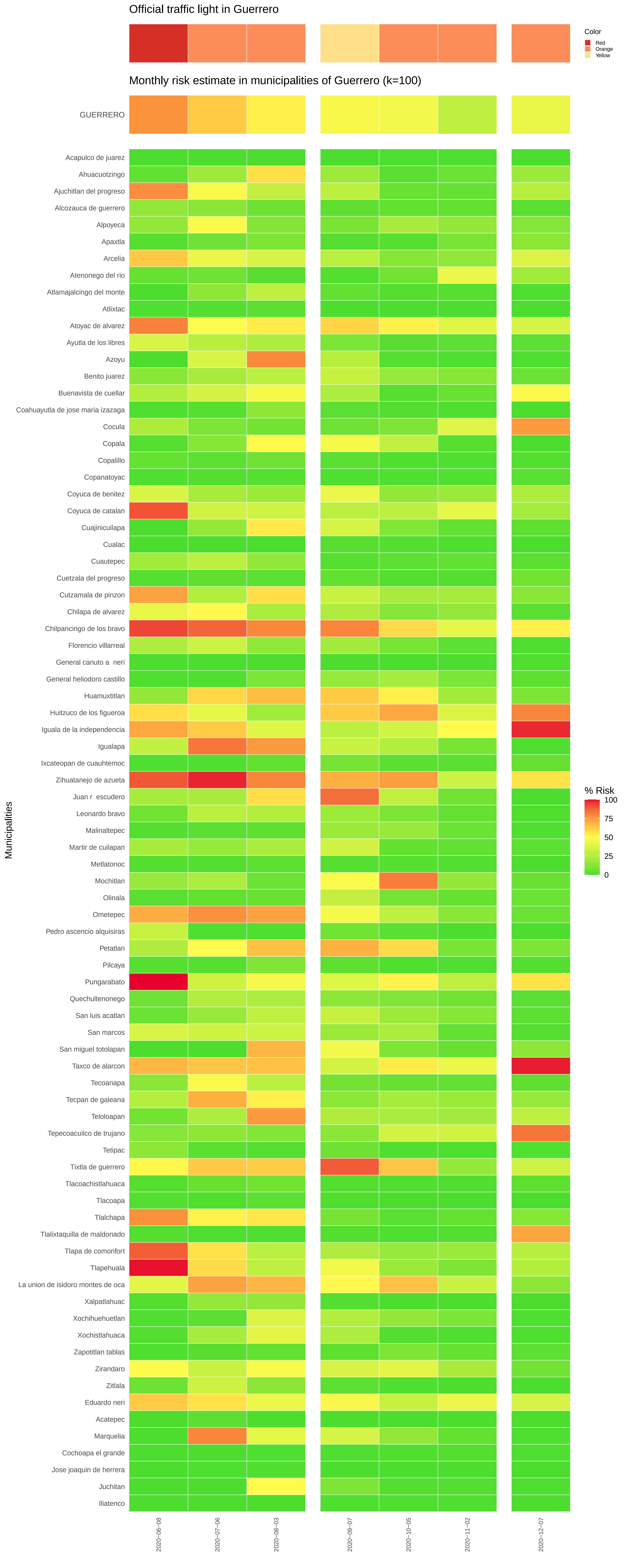
Official traffic light in Guerrero
Color
Red
Orange
Yellow
Monthly risk estimate in municipalities of Guerrero (k=100)
GUERRERO
Acapulco de juarez
Ahuacuotzingo
Ajuchitlan del progreso
Alcozauca de guerrero
Alpoyeca
Apaxtla
Arcelia
Atenonego del rio
Atlamajalcingo del monte
Atlixtac
Atoyac de alvarez
Ayutla de los libres
Azoyu
Benito juarez
Buenavista de cuellar
Coahuayutla de jose maria izazaga
Cocula
Copala
Copalillo
Copanatoyac
Coyuca de benitez
Coyuca de catalan
Cuajinicuilapa
Cualac
Cuautepec
Cuetzala del progreso
Cutzamala de pinzon
Chilapa de alvarez
Chilpancingo de los bravo
Florencio villarreal
General canuto a neri
General heliodoro castillo
Huamuxtitlan
Huitzuco de los figueroa
Iguala de la independencia
Igualapa
Ixcateopan de cuauhtemoc
Zihuatanejo de azueta
Juan r escudero
Leonardo bravo
Malinaltepec
Martir de cuilapan
Metlatonoc
Mochitlan
Olinala
Ometepec
Pedro ascencio alquisiras
Petatlan
Pilcaya
Pungarabato
Quechultenonego
San luis acatlan
San marcos
San miguel totolapan
Taxco de alarcon
Tecoanapa
Tecpan de galeana
Teloloapan
Tepecoacuilco de trujano
Tetipac
Tixtla de guerrero
Tlacoachistlahuaca
Tlacoapa
Tlalchapa
Tlalixtaquilla de maldonado
Tlapa de comonfort
Tlapehuala
La union de isidoro montes de oca
Xalpatlahuac
Xochihuehuetlan
Xochistlahuaca
Zapotitlan tablas
Zirandaro
Zitlala
Eduardo neri
Acatepec
Marquelia
Cochoapa el grande
Jose joaquin de herrera
Juchitan
Iliatenco
Municipalities
% Risk
100
75
50
25
0
2020-06-08
2020-07-06
2020-08-03
2020-09-07
2020-10-05
2020-11-02
2020-12-07

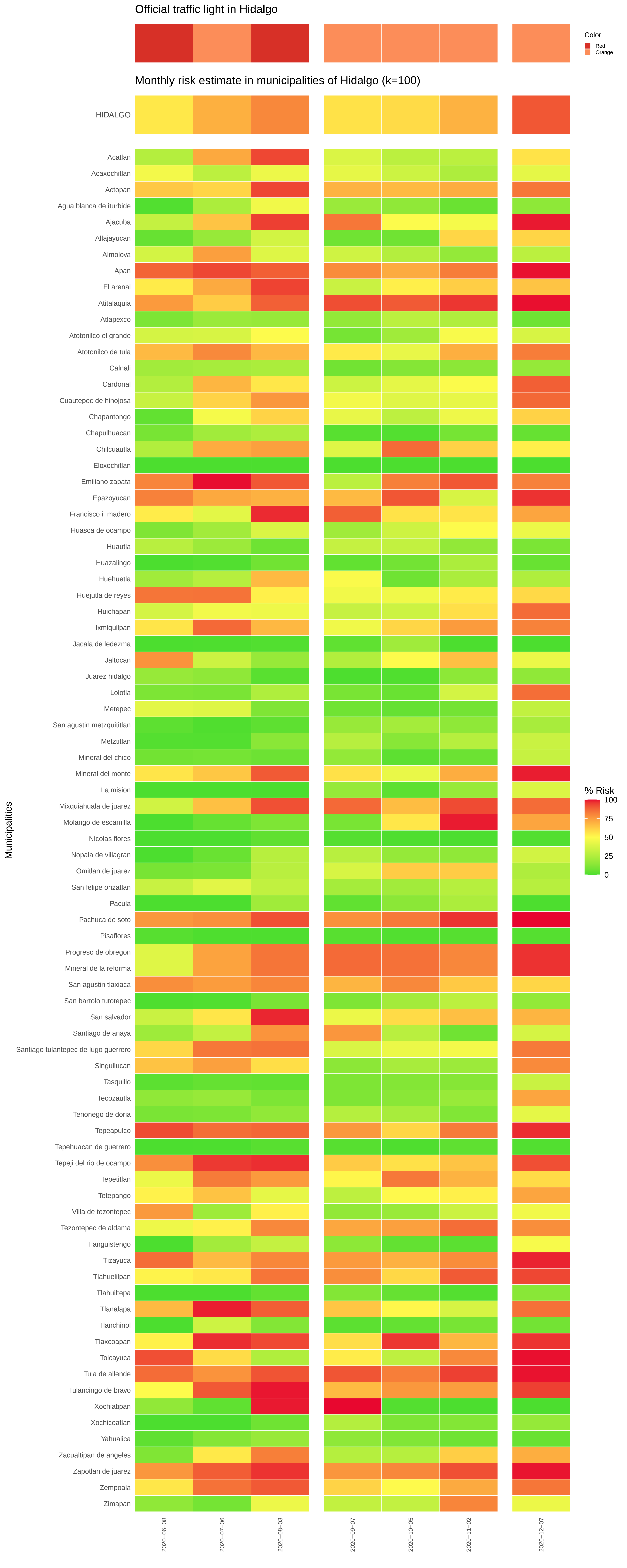
Official traffic light in Hidalgo
Color
Red
Orange
Monthly risk estimate in municipalities of Hidalgo (k=100)
HIDALGO
Acatlan
Acaxochitlan
Actopan
Agua blanca de iturbide
Ajacuba
Alfajayucan
Almoloya
Apan
El arenal
Atitalaquia
Atlapexco
Atotonilco el grande
Atotonilco de tula
Calnali
Cardonal
Cuautepec de hinojosa
Chapantongo
Chapulhuacan
Chilcuautla
Eloxochitlan
Emiliano zapata
Epazoyucan
Francisco i  madero
Huasca de ocampo
Huautla
Huazalingo
Huehuetla
Huejutla de reyes
Huichapan
Ixmiquilpan
Jacala de ledezma
Jaltocan
Juarez hidalgo
Lolotla
Metepec
San agustin metzquititlan
Metztitlan
Mineral del chico
Mineral del monte
La mision
Mixquiahuala de juarez
Molango de escamilla
Nicolas flores
Nopala de villagran
Omitlan de juarez
San felipe orizatlan
Pacula
Pachuca de soto
Pisaflores
Progreso de obregon
Mineral de la reforma
San agustin tlaxiaca
San bartolo tutotepec
San salvador
Santiago de anaya
Santiago tulantepec de lugo guerrero
Singuilucan
Tasquillo
Tecozautla
Tenonego de doria
Tepeapulco
Tepehuacan de guerrero
Tepeji del rio de ocampo
Tepetitlan
Tetepango
Villa de tezontepec
Tezontepec de aldama
Tianguistengo
Tizayuca
Tlahuelilpan
Tlahuiltepa
Tlanalapa
Tlanchinol
Tlaxcoapan
Tolcayuca
Tula de allende
Tulancingo de bravo
Xochiatipan
Xochicoatlan
Yahualica
Zacualtipan de angeles
Zapotlan de juarez
Zempoala
Zimapan
Municipalities
% Risk
100
75
50
25
0
2020-06-08
2020-07-06
2020-08-03
2020-09-07
2020-10-05
2020-11-02
2020-12-07

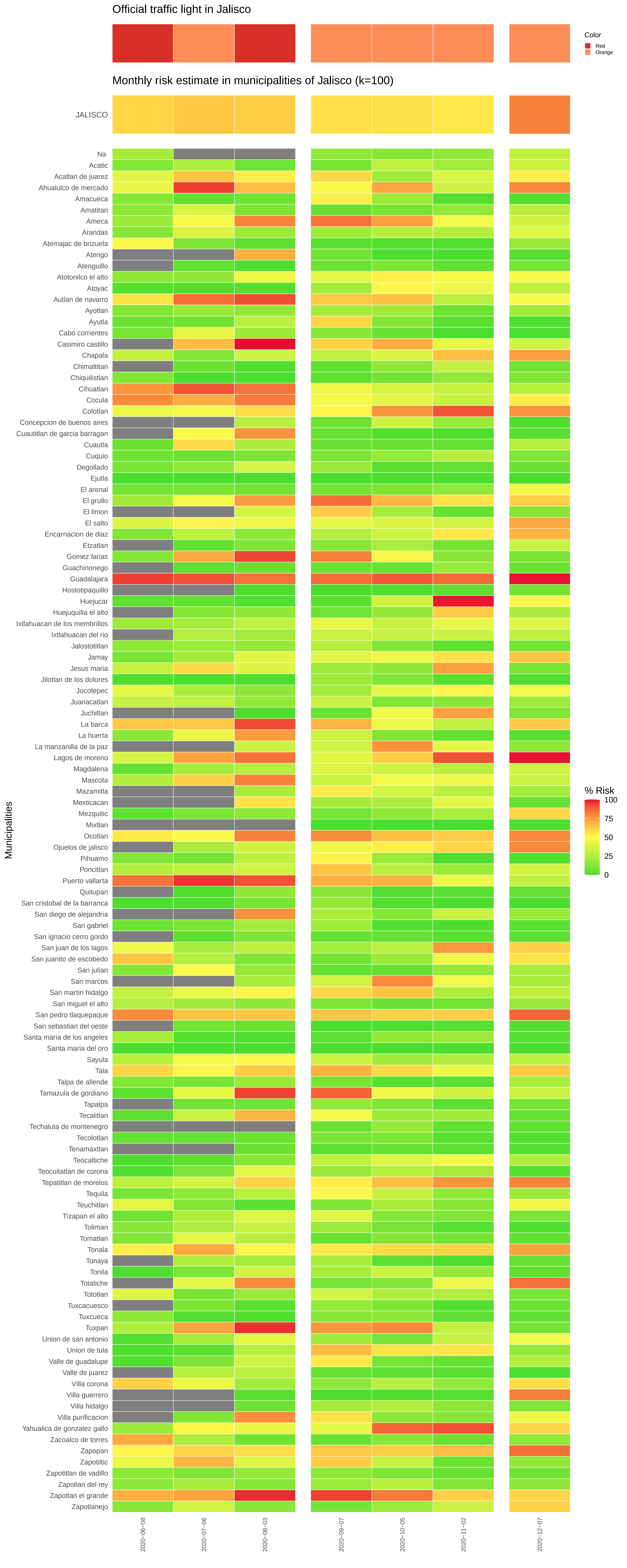
Official traffic light in Jalisco
Color
Red
Orange
Monthly risk estimate in municipalities of Jalisco (k=100)
JALISCO
Na
Acatic
Acatlan de juarez
Ahualulco de mercado
Amacueca
Amatitan
Ameca
Arandas
Atemajac de brizuela
Atengo
Atenguillo
Atotonilco el alto
Atoyac
Autlan de navarro
Ayotlan
Ayutla
Cabo corrientes
Casimiro castillo
Chapala
Chimaltitan
Chiquilistlan
Cihuatlan
Cocula
Colotlan
Concepcion de buenos aires
Cuautitlan de garcia barragan
Cuautla
Cuquio
Degollado
Ejutla
El arenal
El grullo
El limon
El salto
Encarnacion de diaz
Etzatlan
Gomez farias
Guachinonego
Guadalajara
Hostotipaquillo
Huejucar
Huejuquilla el alto
Ixtlahuacan de los membrillos
Ixtlahuacan del rio
Jalostotitlan
Jamay
Jesus maria
Jilotlan de los dolores
Jocotepec
Juanacatlan
Juchitlan
La barca
La huerta
La manzanilla de la paz
Lagos de moreno
Magdalena
Mascota
Mazamitla
Mexticacan
Mezquitic
Mixtlan
Ocotlan
Ojuelos de jalisco
Pihuamo
Poncitlan
Puerto vallarta
Quitupan
San cristobal de la barranca
San diego de alejandria
San gabriel
San ignacio cerro gordo
San juan de los lagos
San juanito de escobedo
San julian
San marcos
San martin hidalgo
San miguel el alto
San pedro tlaquepaque
San sebastian del oeste
Santa maria de los angeles
Santa maria del oro
Sayula
Tala
Talpa de allende
Tamazula de gordiano
Tapalpa
Tecalitlan
Techaluta de montenegro
Tecolotlan
Tenamaxtlan
Teocaltiche
Teocuitatlan de corona
Tepatitlan de morelos
Tequila
Teuchitlan
Tizapan el alto
Toliman
Tomatlan
Tonala
Tonaya
Tonila
Totatiche
Tototlan
Tuxcacuesco
Tuxcueca
Tuxpan
Union de san antonio
Union de tula
Valle de guadalupe
Valle de juarez
Villa corona
Villa guerrero
Villa hidalgo
Villa purificacion
Yahualica de gonzalez gallo
Zacoalco de torres
Zapopan
Zapotiltic
Zapotitlan de vadillo
Zapotlan del rey
Zapotlan el grande
Zapotlanejo
Municipalities
% Risk
100
75
50
25
0
2020−06−08
2020−07−06
2020−08−03
2020−09−07
2020−10−05
2020−11−02
2020−12−07

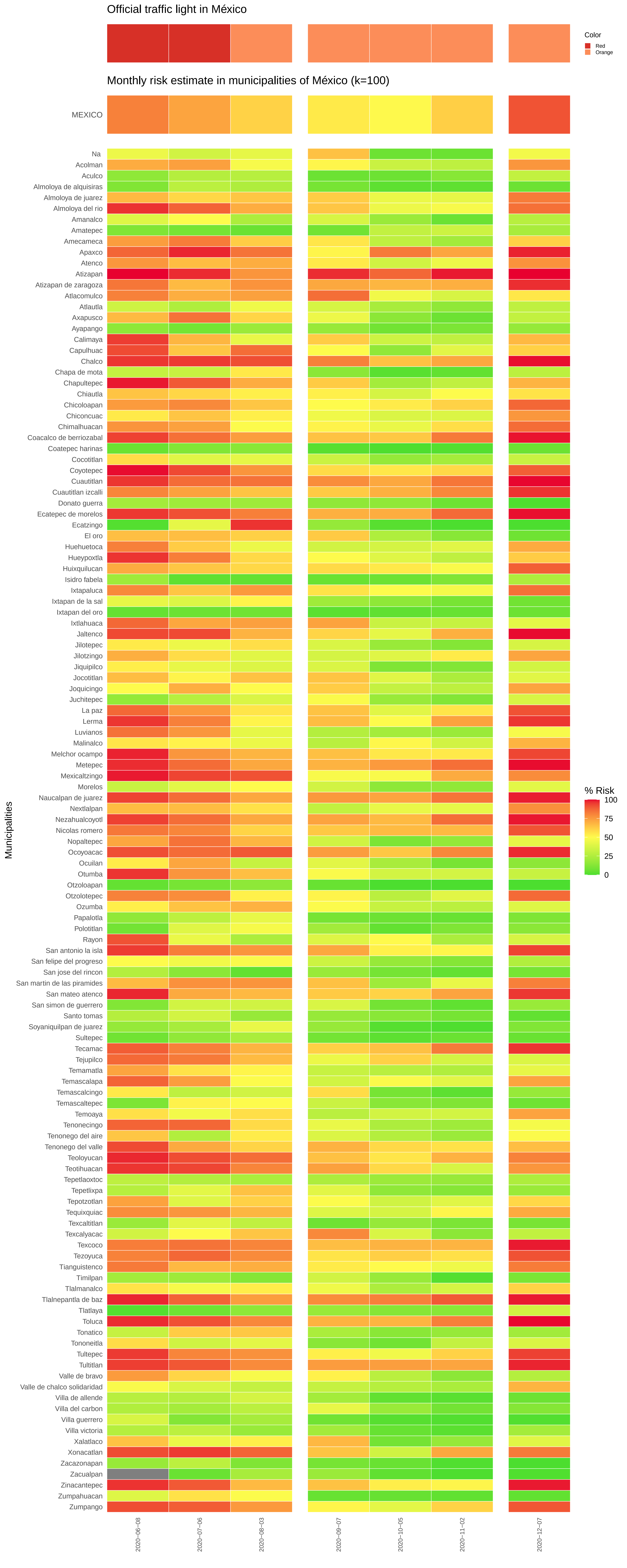

Official traffic light in México
Color
Red
Orange
Monthly risk estimate in municipalities of México (k=100)
MEXICO
Municipalities
% Risk
100
75
50
25
0
Na
Acolman
Aculco
Almoloya de alquisiras
Almoloya de juarez
Almoloya del rio
Amanalco
Amatepec
Amecameca
Apaxco
Atenco
Atizapan
Atizapan de zaragoza
Atlacomulco
Atlautla
Axapusco
Ayapango
Calimaya
Capulhuac
Chalco
Chapa de mota
Chapultepec
Chiautla
Chicoloapan
Chiconcuac
Chimalhuacan
Coacalco de berriozabal
Coatepec harinas
Cocotitlan
Coyotepec
Cuautitlan
Cuautitlan izcalli
Donato guerra
Ecatepec de morelos
Ecatzingo
El oro
Huehuetoca
Hueypoxtla
Huixquilucan
Isidro fabela
Ixtapaluca
Ixtapan de la sal
Ixtapan del oro
Ixtlahuaca
Jaltenco
Jilotepec
Jilotzingo
Jiquipilco
Jocotitlan
Joquicingo
Juchitepec
La paz
Lerma
Luvianos
Malinalco
Melchor ocampo
Metepec
Mexicaltzingo
Morelos
Naucalpan de juarez
Nextlalpan
Nezahualcoyotl
Nicolas romero
Nopaltepec
Ocoyoacac
Ocuilan
Otumba
Otzoloapan
Otzolotepec
Ozumba
Papalotla
Polotitlan
Rayon
San antonio la isla
San felipe del progreso
San jose del rincon
San martin de las piramides
San mateo atenco
San simon de guerrero
Santo tomas
Soyaniquilpan de juarez
Sultepec
Tecamac
Tejupilco
Temamatla
Temascalapa
Temascalcingo
Temascaltepec
Temoaya
Tenonecingo
Tenonego del aire
Tenonego del valle
Teoloyucan
Teotihuacan
Tepetlaoxtoc
Tepetlixpa
Tepotzotlan
Tequixquiac
Texcaltitlan
Texcalyacac
Texcoco
Tezoyuca
Tianguistenco
Timilpan
Tlalmanalco
Tlalnepantla de baz
Tlatlaya
Toluca
Tonatico
Tononeitla
Tultepec
Tultitlan
Valle de bravo
Valle de chalco solidaridad
Villa de allende
Villa del carbon
Villa guerrero
Villa victoria
Xalatlaco
Xonacatlan
Zacazonapan
Zacualpan
Zinacantepec
Zumpahuacan
Zumpango
2020-06-08
2020-07-06
2020-08-03
2020-09-07
2020-10-05
2020-11-02
2020-12-07

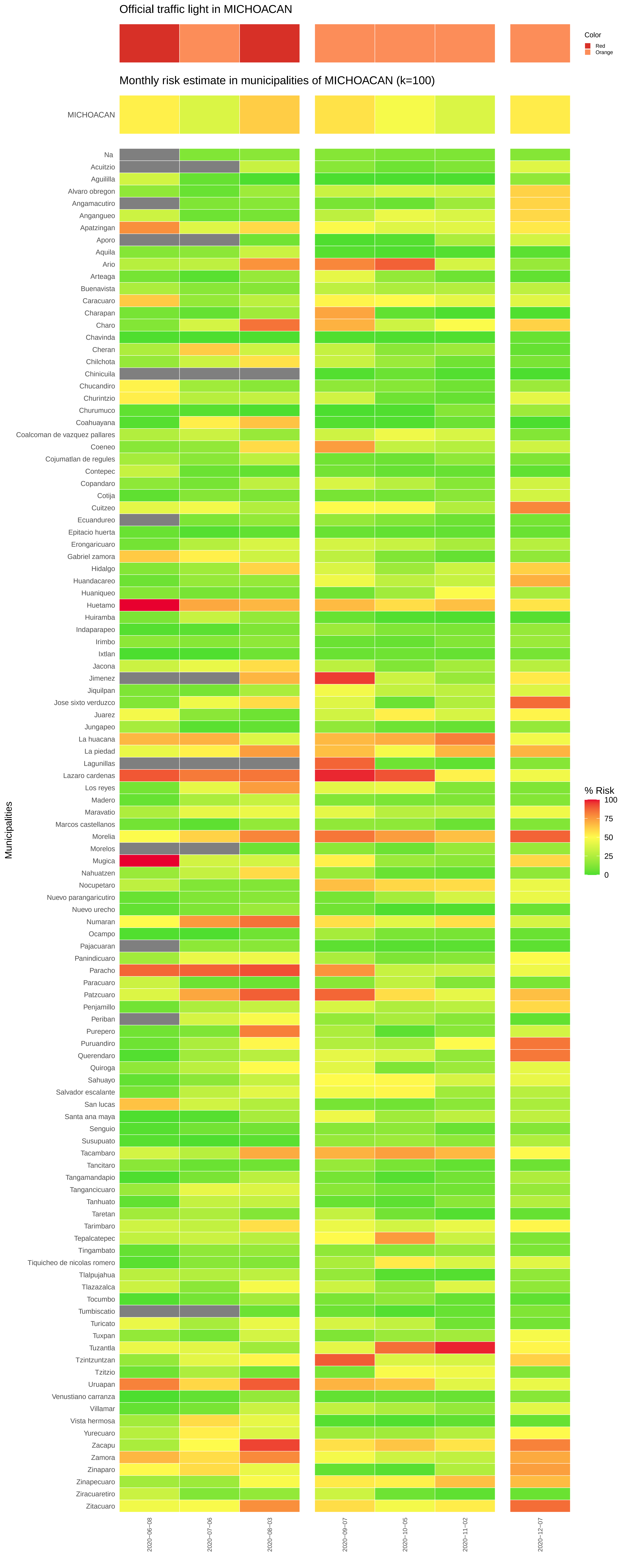

Official traffic light in MICHOACAN
Color
Red
Orange
Monthly risk estimate in municipalities of MICHOACAN (k=100)
MICHOACAN
Municipalities
Na
Acuitzio
Aguililla
Alvaro obregon
Angamacutiro
Angangueo
Apatzingan
Aporo
Aquila
Ario
Arteaga
Buenavista
Caracuaro
Charapan
Charo
Chavinda
Cheran
Chilchota
Chinicuila
Chucandiro
Churintzio
Churumuco
Coahuayana
Coalcoman de vazquez pallares
Coeneo
Cojumatlan de regules
Contepec
Copandaro
Cotija
Cuitzeo
Ecuandureo
Epitacio huerta
Erongaricuaro
Gabriel zamora
Hidalgo
Huandacareo
Huaniqueo
Huetamo
Huiramba
Indaparapeo
Irimbo
Ixtlan
Jacona
Jimenez
Jiquilpan
Jose sixto verduzco
Juarez
Jungapeo
La huacana
La piedad
Lagunillas
Lazaro cardenas
Los reyes
Madero
Maravatio
Marcos castellanos
Morelia
Morelos
Mugica
Nahuatzen
Nocupetaro
Nuevo parangaricutiro
Nuevo urecho
Numaran
Ocampo
Pajacuaran
Panindicuaro
Paracho
Paracuaro
Patzcuaro
Penjamillo
Periban
Purepero
Puruandiro
Querendaro
Quiroga
Sahuayo
Salvador escalante
San lucas
Santa ana maya
Senguio
Susupuato
Tacambaro
Tancitaro
Tangamandapio
Tangancicuaro
Tanhuato
Taretan
Tarimbaro
Tepalcatepec
Tingambato
Tiquicheo de nicolas romero
Tlalpujahua
Tlazazalca
Tocumbo
Tumbiscatio
Turicato
Tuxpan
Tuzantla
Tzintzuntzan
Tzitzio
Uruapan
Venustiano carranza
Villamar
Vista hermosa
Yurecuaro
Zacapu
Zamora
Zinaparo
Zinapecuaro
Ziracuaretiro
Zitacuaro
% Risk
100
75
50
25
0
2020–06–08
2020–07–06
2020–08–03
2020–09–07
2020–10–05
2020–11–02
2020–12–07

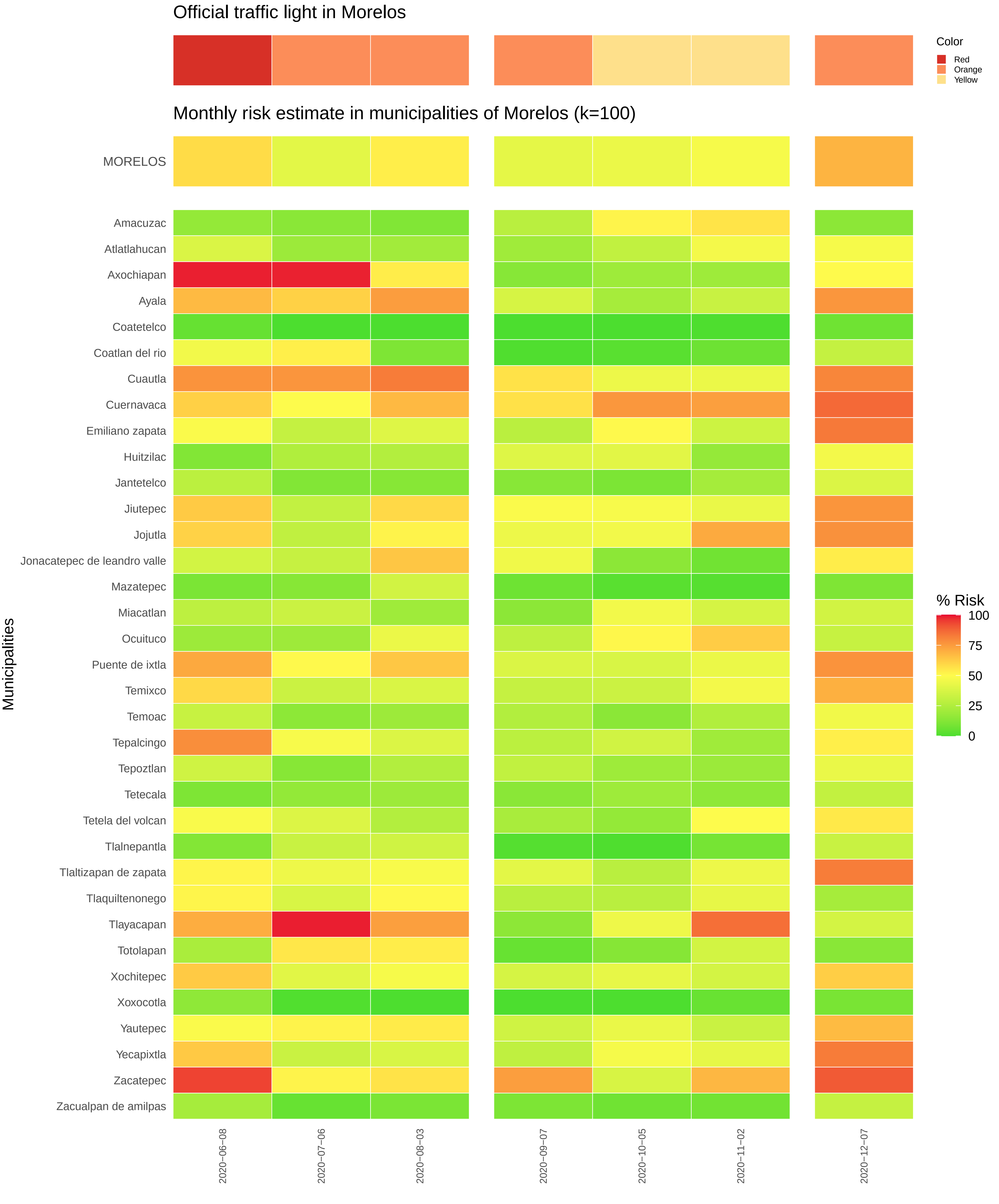

Official traffic light in Morelos
Color
Red
Orange
Yellow
Monthly risk estimate in municipalities of Morelos (k=100)
MORELOS
Amacuzac
Atlatlahucan
Axochiapan
Ayala
Coatetelco
Coatlan del rio
Cuautla
Cuernavaca
Emiliano zapata
Huitzilac
Jantetelco
Jiutepec
Jojutla
Jonacatepec de leandro valle
Mazatepec
Miacatlan
Ocuituco
Puente de ixtla
Temixco
Temoac
Tepalcingo
Tepoztlan
Tetecala
Tetela del volcan
Tlalnepantla
Tlaltizapan de zapata
Tlaquiltenonego
Tlayacapan
Totolapan
Xochitepec
Xoxocotla
Yautepec
Yecapixtla
Zacatepec
Zacualpan de amilpas
Municipalities
% Risk
100
75
50
25
0
2020−06−08
2020−07−06
2020−08−03
2020−09−07
2020−10−05
2020−11−02
2020−12−07

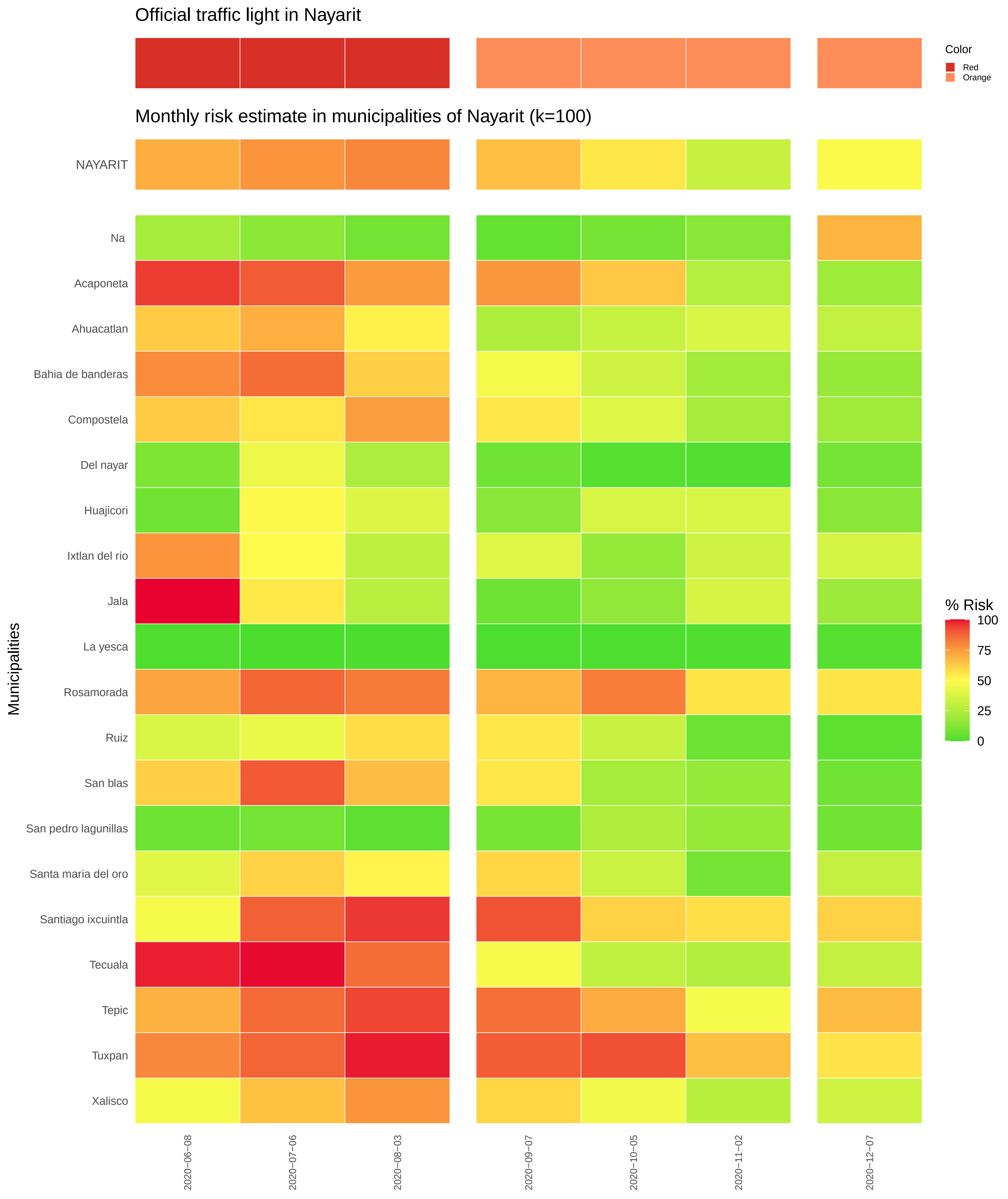

Official traffic light in Nayarit
Color
Red
Orange
Monthly risk estimate in municipalities of Nayarit (k=100)
NAYARIT
Na
Acaponeta
Ahuacatlan
Bahia de banderas
Compostela
Del nayar
Huajicori
Ixtlan del rio
Jala
La yesca
Rosamorada
Ruiz
San blas
San pedro lagunillas
Santa maria del oro
Santiago ixcuintla
Tecuala
Tepic
Tuxpan
Xalisco
Municipalities
% Risk
100
75
50
25
0
2020−06−08
2020−07−06
2020−08−03
2020−09−07
2020−10−05
2020−11−02
2020−12−07

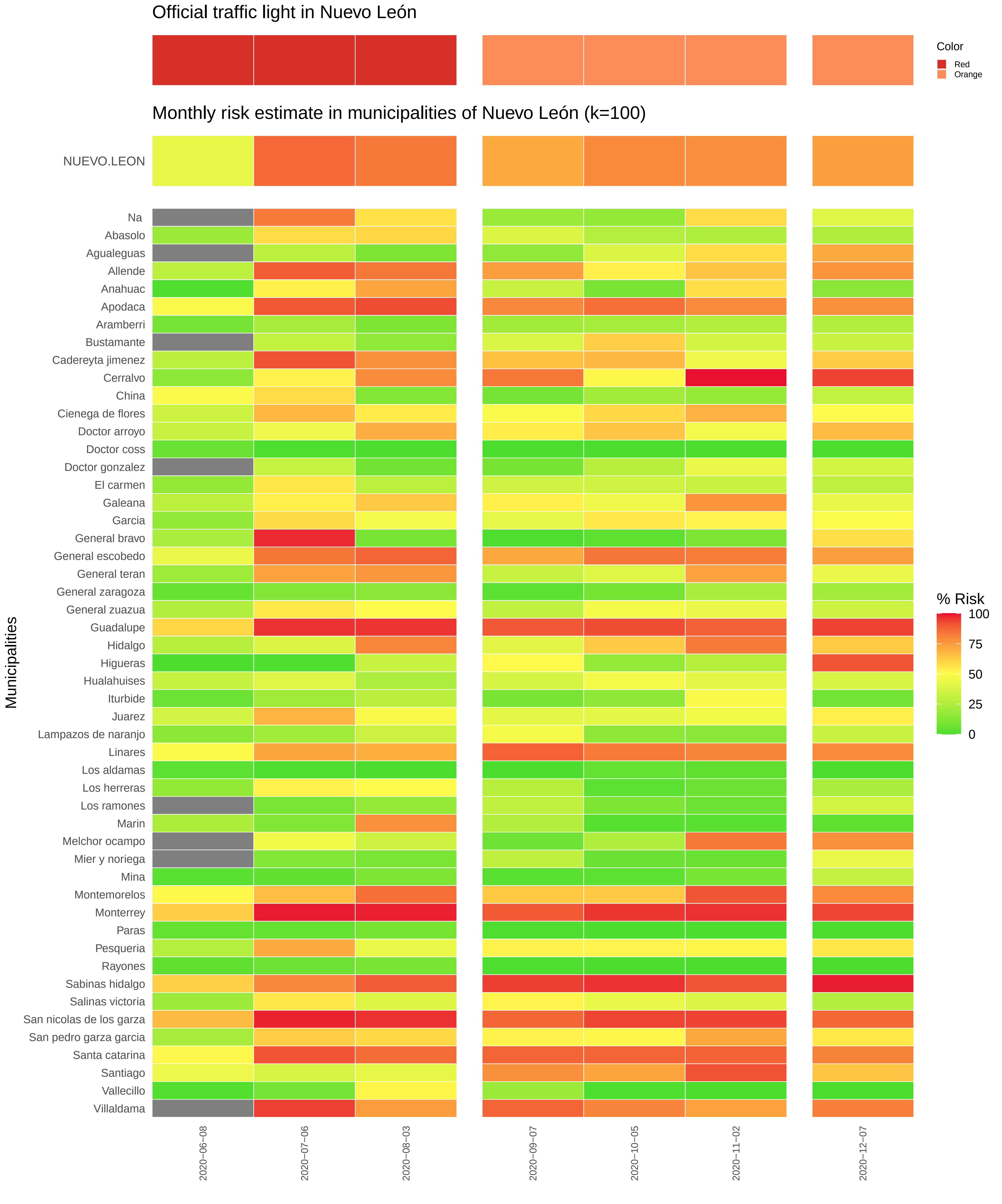

Official traffic light in Nuevo León
Color
Red
Orange
Monthly risk estimate in municipalities of Nuevo León (k=100)
NUEVO.LEON
Municipalities
Na
Abasolo
Agualeguas
Allende
Anahuac
Apodaca
Aramberri
Bustamante
Cadereyta jimenez
Cerralvo
China
Cienega de flores
Doctor arroyo
Doctor coss
Doctor gonzalez
El carmen
Galeana
Garcia
General bravo
General escobedo
General teran
General zaragoza
General zuazua
Guadalupe
Hidalgo
Higueras
Hualahuises
Iturbide
Juarez
Lampazos de naranjo
Linares
Los aldamas
Los herreras
Los ramones
Marin
Melchor ocampo
Mier y noriega
Mina
Montemorelos
Monterrey
Paras
Pesqueria
Rayones
Sabinas hidalgo
Salinas victoria
San nicolas de los garza
San pedro garza garcia
Santa catarina
Santiago
Vallecillo
Villaldama
% Risk
100
75
50
25
0
2020−06−08
2020−07−06
2020−08−03
2020−09−07
2020−10−05
2020−11−02
2020−12−07

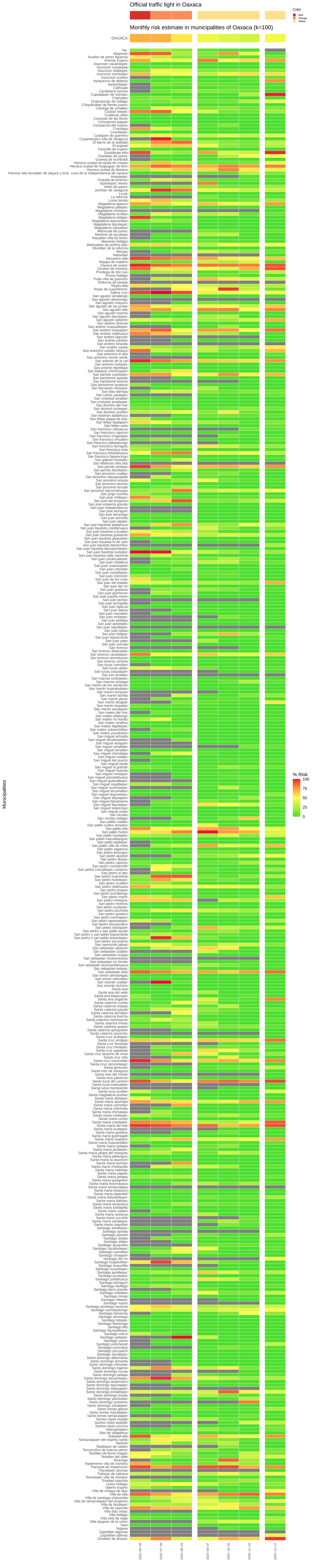
Official traffic light in Oaxaca
Monthly risk estimate in municipalities of Oaxaca (k=100)
OAXACA
Color
Red
Orange
Yellow
% Risk
100
75
50
25
0
Municipalities

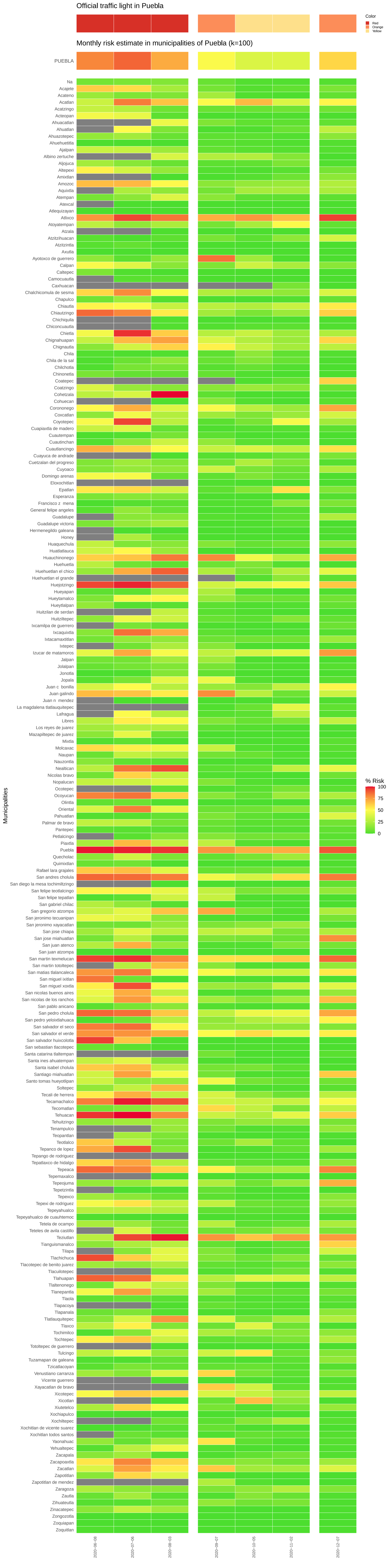
Official traffic light in Puebla
Color
Red
Orange
Yellow
Monthly risk estimate in municipalities of Puebla (k=100)
PUEBLA
Municipalities
% Risk
100
75
50
25
0
Na
Acajete
Acateno
Acatlan
Acatzingo
Acteopan
Ahuacatlan
Ahuatlan
Ahuazotepec
Ahuehuetitla
Ajalpan
Albino zertuche
Aljojuca
Altepexi
Amixtlan
Amozoc
Aquixtla
Atempan
Atexcal
Atlequizayan
Atlixco
Atoyatempan
Atzala
Atzitzihuacan
Atzitzintla
Axutla
Ayotoxco de guerrero
Calpan
Caltepec
Camocuautla
Caxhuacan
Chalchicomula de sesma
Chapulco
Chiautla
Chiautzingo
Chichiquila
Chiconcuautla
Chietla
Chignahuapan
Chignautla
Chila
Chila de la sal
Chilchotla
Chinontla
Coatepec
Coatzingo
Cohetzala
Cohuecan
Coronango
Coxcatlan
Coyotepec
Cuapiaxtla de madero
Cuautempan
Cuautinchan
Cuautlancingo
Cuayuca de andrade
Cuetzalan del progreso
Cuyoaco
Domingo arenas
Eloxochitlan
Epatlan
Esperanza
Francisco z mena
General felipe angeles
Guadalupe
Guadalupe victoria
Hermenegildo galeana
Honey
Huaquechula
Huatlatlauca
Huauchinango
Huehuetla
Huehuetlan el chico
Huehuetlan el grande
Huejotzingo
Hueyapan
Hueytamalco
Hueytlalpan
Huitzilan de serdan
Huitziltepec
Ixcamilpa de guerrero
Ixcaquixtla
Ixtacamaxtitlan
Ixtepec
Izucar de matamoros
Jalpan
Jolalpan
Jonotla
Jopala
Juan c bonilla
Juan galindo
Juan n mendez
La magdalena tlatlauquitepec
Lafragua
Libres
Los reyes de juarez
Mazapiltepec de juarez
Mixtla
Molcaxac
Naupan
Nauzontla
Nealtican
Nicolas bravo
Nopalucan
Ocotepec
Ocoyucan
Olintla
Oriental
Pahuatlan
Palmar de bravo
Pantepec
Petlalcingo
Piaxtla
Puebla
Quecholac
Quimixtlan
Rafael lara grajales
San andres cholula
San diego la mesa tochimiltzingo
San felipe teotlalcingo
San felipe tepatlan
San gabriel chilac
San gregorio atzompa
San jeronimo tecuanipan
San jeronimo xayacatlan
San jose chiapa
San jose miahuatlan
San juan atenco
San juan atzompa
San martin texmelucan
San martin totoltepec
San matias tlalancaleca
San miguel ixitlan
San miguel xoxtla
San nicolas buenos aires
San nicolas de los ranchos
San pablo anicano
San pedro cholula
San pedro yeloixtlahuaca
San salvador el seco
San salvador el verde
San salvador huixcolotla
San sebastian tlacotepec
Santa catarina tlaltempan
Santa ines ahuatempan
Santa isabel cholula
Santiago miahuatlan
Santo tomas hueyotlipan
Soltepec
Tecali de herrera
Tecamachalco
Tecomatlan
Tehuacan
Tehuitzingo
Tenampulco
Teopantlan
Teotlalco
Tepanco de lopez
Tepango de rodriguez
Tepatlaxco de hidalgo
Tepeaca
Tepemaxalco
Tepeojuma
Tepetzintla
Tepexco
Tepexi de rodriguez
Tepeyahualco
Tepeyahualco de cuauhtemoc
Tetela de ocampo
Teteles de avila castillo
Teziutlan
Tianguismanalco
Tilapa
Tlachichuca
Tlacotepec de benito juarez
Tlacuilotepec
Tlahuapan
Tlaltenango
Tlanepantla
Tlaola
Tlapacoya
Tlapanala
Tlatlauquitepec
Tlaxco
Tochimilco
Tochtepec
Totoltepec de guerrero
Tulcingo
Tuzamapan de galeana
Tzicatlacoyan
Venustiano carranza
Vicente guerrero
Xayacatlan de bravo
Xicotepec
Xicotlan
Xiutetelco
Xochiapulco
Xochiltepec
Xochitlan de vicente suarez
Xochitlan todos santos
Yaonahuac
Yehualtepec
Zacapala
Zacapoaxtla
Zacatlan
Zapotitlan
Zapotitlan de mendez
Zaragoza
Zautla
Zihuateutla
Zinacatepec
Zongozotla
Zoquiapan
Zoquitlan
2020−06−08
2020−07−06
2020−08−03
2020−09−07
2020−10−05
2020−11−02
2020−12−07

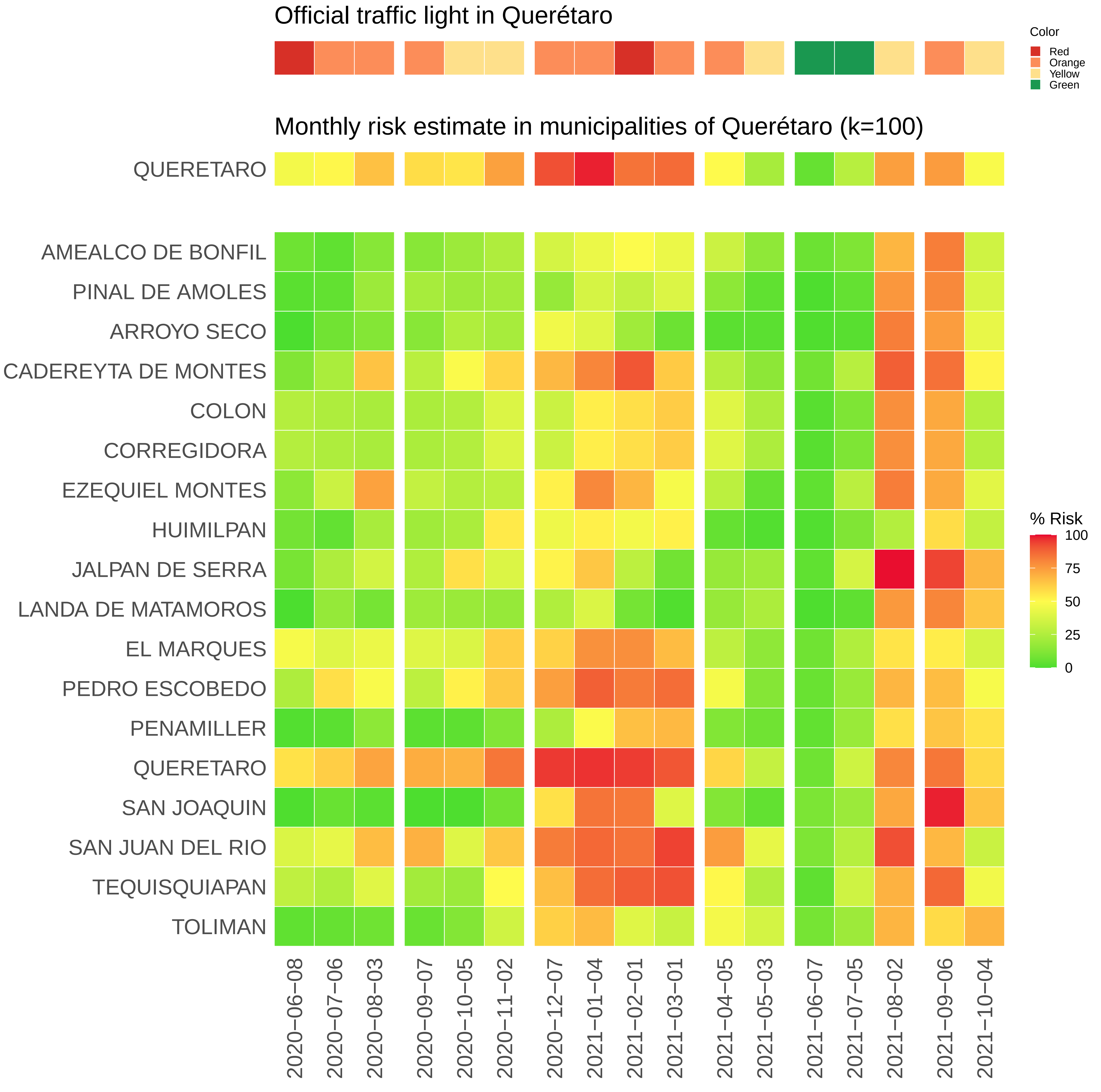

Official traffic light in Querétaro
Color
Red
Orange
Yellow
Green
Monthly risk estimate in municipalities of Querétaro (k=100)
QUERETARO
AMEALCO DE BONFIL
PINAL DE AMOLES
ARROYO SECO
CADEREYTA DE MONTES
COLON
CORREGIDORA
EZEQUIEL MONTES
HUIMILPAN
JALPAN DE SERRA
LANDA DE MATAMOROS
EL MARQUES
PEDRO ESCOBEDO
PENAMILLER
QUERETARO
SAN JOAQUIN
SAN JUAN DEL RIO
TEQUISQUIAPAN
TOLIMAN
% Risk
100
75
50
25
0
2020−06−08
2020−07−06
2020−08−03
2020−09−07
2020−10−05
2020−11−02
2020−12−07
2021−01−04
2021−02−01
2021−03−01
2021−04−05
2021−05−03
2021−06−07
2021−07−05
2021−08−02
2021−09−06
2021−10−04

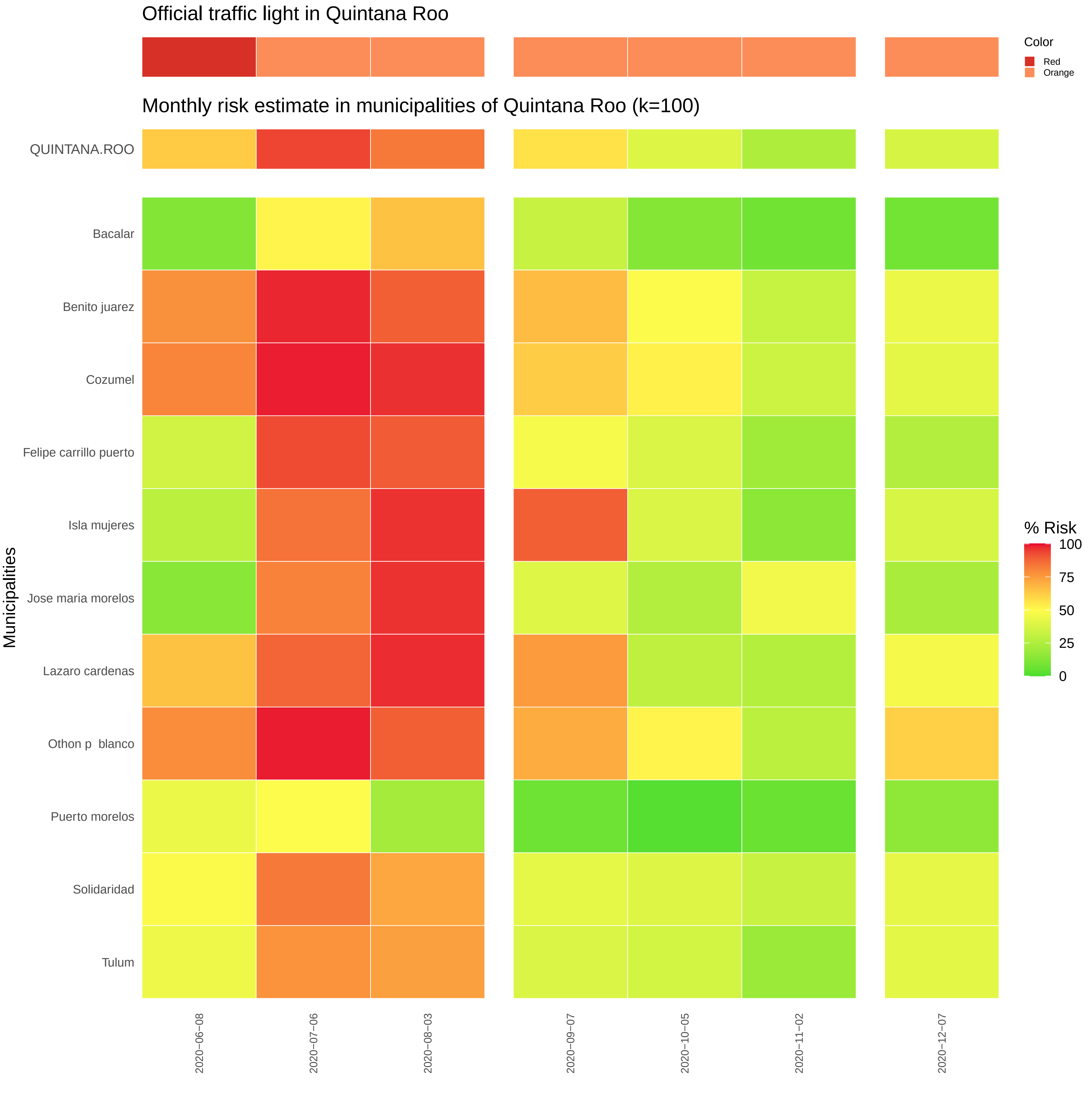

Official traffic light in Quintana Roo
Color
Red
Orange
Monthly risk estimate in municipalities of Quintana Roo (k=100)
QUINTANA.ROO
Bacalar
Benito juarez
Cozumel
Felipe carrillo puerto
Isla mujeres
Jose maria morelos
Lazaro cardenas
Othon p blanco
Puerto morelos
Solidaridad
Tulum
Municipalities
% Risk
100
75
50
25
0
2020−06−08
2020−07−06
2020−08−03
2020−09−07
2020−10−05
2020−11−02
2020−12−07

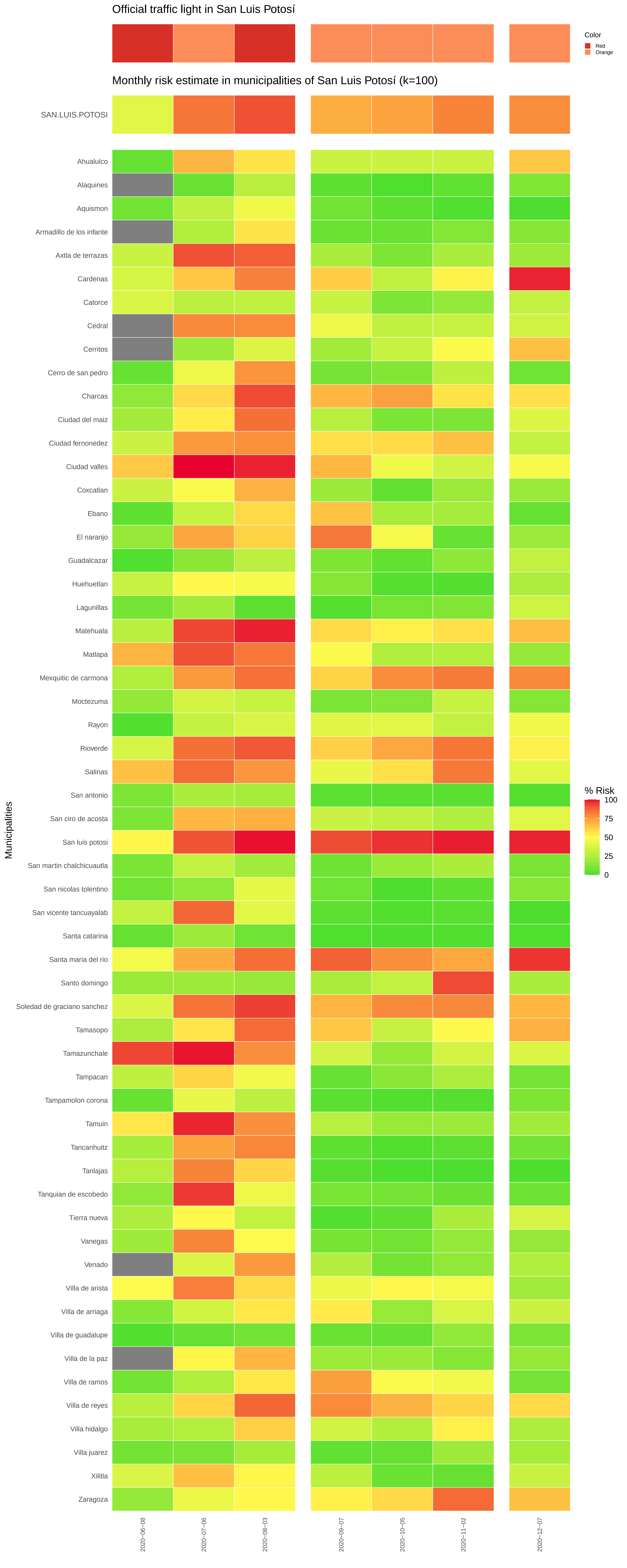

Official traffic light in San Luis Potosí
Color
Red
Orange
Monthly risk estimate in municipalities of San Luis Potosí (k=100)
SAN.LUIS.POTOSI
Municipalities
Ahualulco
Alaquines
Aquismon
Armadillo de los infante
Axtla de terrazas
Cardenas
Catorce
Cedral
Cerritos
Cerro de san pedro
Charcas
Ciudad del maiz
Ciudad fernonedez
Ciudad valles
Coxcatlan
Ebano
El naranjo
Guadalcazar
Huehuetlan
Lagunillas
Matehuala
Matlapa
Mexquitic de carmona
Moctezuma
Rayon
Rioverde
Salinas
San antonio
San ciro de acosta
San luis potosi
San martin chalchicuautla
San nicolas tolentino
San vicente tancuayalab
Santa catarina
Santa maria del rio
Santo domingo
Soledad de graciano sanchez
Tamasopo
Tamazunchale
Tampacan
Tampamolon corona
Tamuin
Tancanhuitz
Tanlajas
Tanquian de escobedo
Tierra nueva
Vanegas
Venado
Villa de arista
Villa de arriaga
Villa de guadalupe
Villa de la paz
Villa de ramos
Villa de reyes
Villa hidalgo
Villa juarez
Xilitla
Zaragoza
% Risk
100
75
50
25
0
2020−06−08
2020−07−06
2020−08−03
2020−09−07
2020−10−05
2020−11−02
2020−12−07

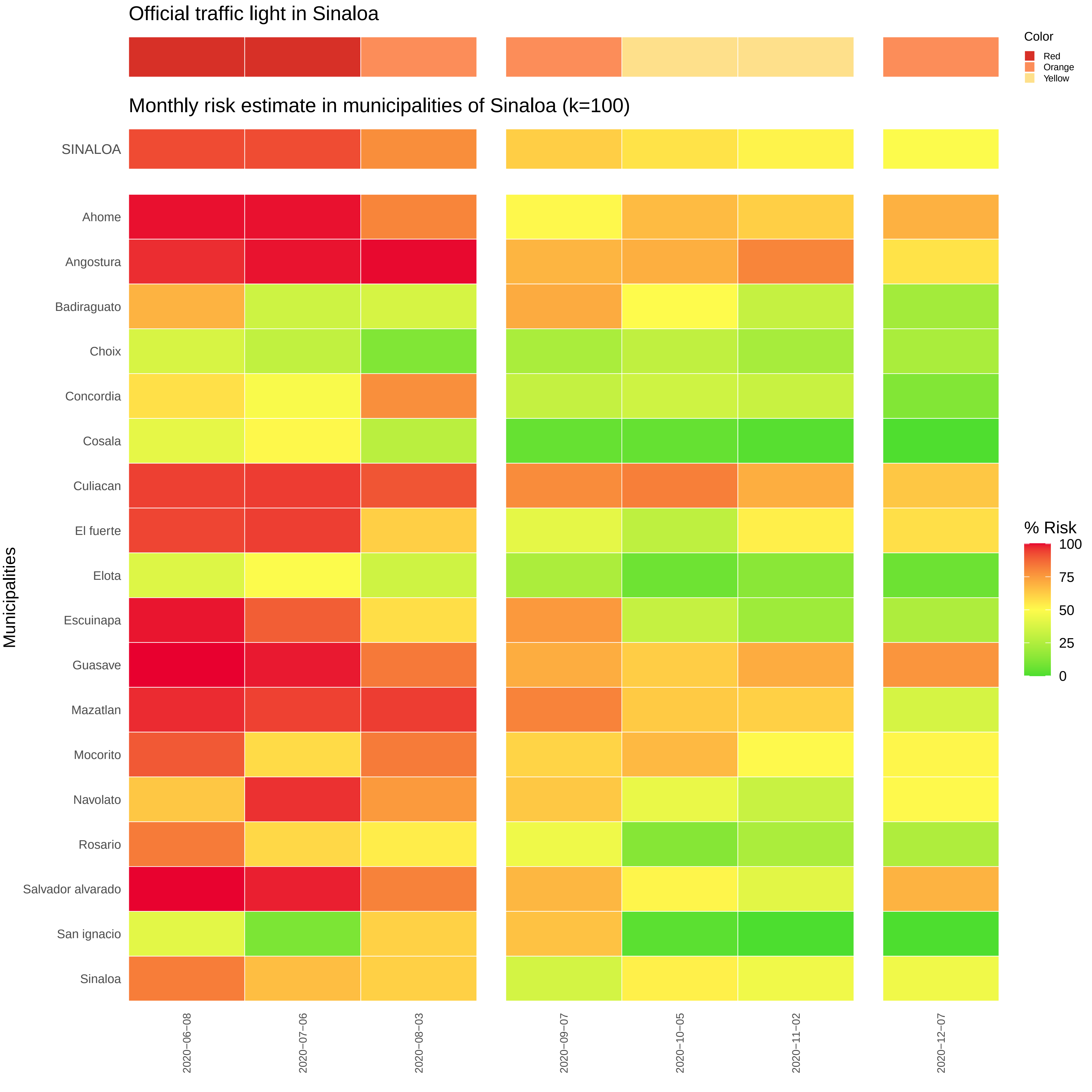

Official traffic light in Sinaloa
Color
Red
Orange
Yellow
Monthly risk estimate in municipalities of Sinaloa (k=100)
SINALOA
Ahome
Angostura
Badiraguato
Choix
Concordia
Cosala
Culiacan
El fuerte
Elota
Escuinapa
Guasave
Mazatlan
Mocorito
Navolato
Rosario
Salvador alvarado
San ignacio
Sinaloa
Municipalities
% Risk
100
75
50
25
0
2020−06−08
2020−07−06
2020−08−03
2020−09−07
2020−10−05
2020−11−02
2020−12−07

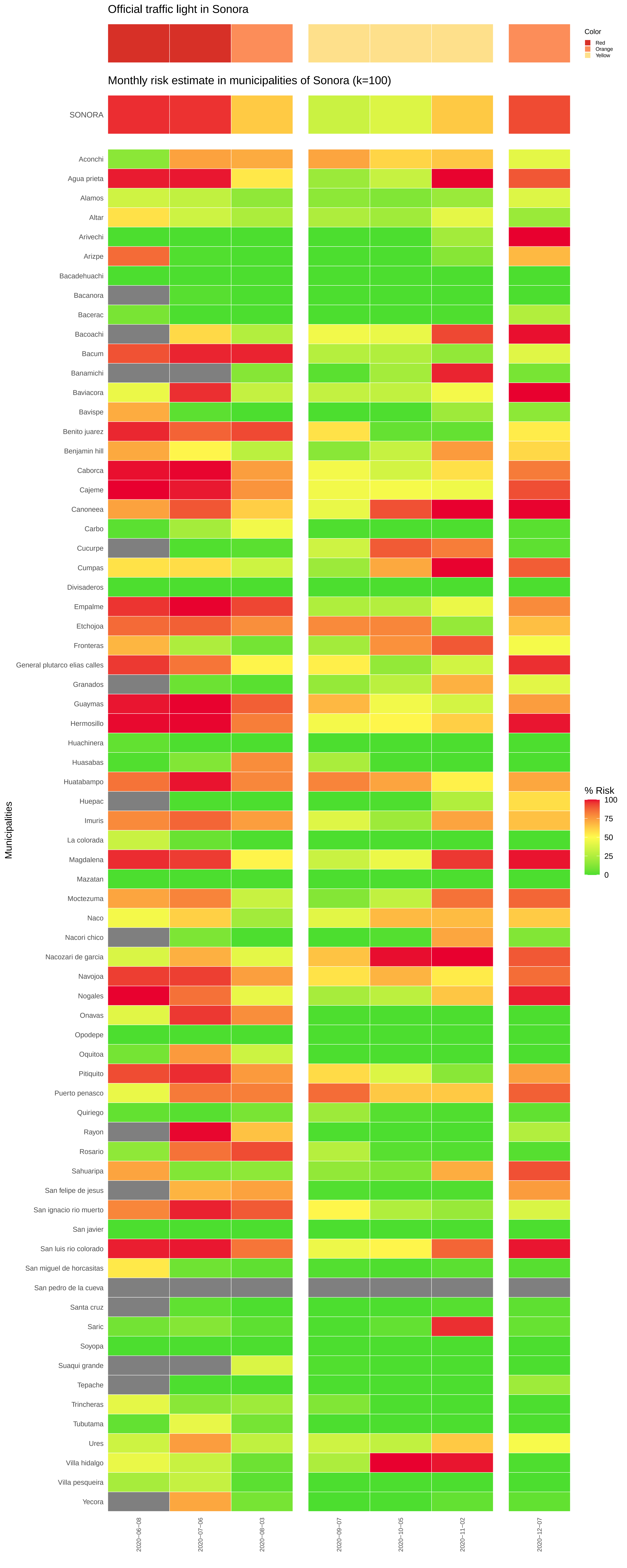

Official traffic light in Sonora
Color
Red
Orange
Yellow
Monthly risk estimate in municipalities of Sonora (k=100)
SONORA
Aconchi
Agua prieta
Alamos
Altar
Arivechi
Arizpe
Bacadehuachi
Bacanora
Bacerac
Bacoachi
Bacum
Banamichi
Baviacora
Bavispe
Benito juarez
Benjamin hill
Caborca
Cajeme
Canoneea
Carbo
Cucurpe
Cumpas
Divisaderos
Empalme
Etchojoa
Fronteras
General plutarco elias calles
Granados
Guaymas
Hermosillo
Huachinera
Huasabas
Huatabampo
Huepac
Imuris
La colorada
Magdalena
Mazatan
Moctezuma
Naco
Nacori chico
Nacozari de garcia
Navojoa
Nogales
Onavas
Opodepe
Oquitoa
Pitiquito
Puerto penasco
Quiriego
Rayon
Rosario
Sahuaripa
San felipe de jesus
San ignacio rio muerto
San javier
San luis rio colorado
San miguel de horcasitas
San pedro de la cueva
Santa cruz
Saric
Soyopa
Suaqui grande
Tepache
Trincheras
Tubutama
Ures
Villa hidalgo
Villa pesqueira
Yecora
Municipalities
% Risk
100
75
50
25
0
2020−06−08
2020−07−06
2020−08−03
2020−09−07
2020−10−05
2020−11−02
2020−12−07

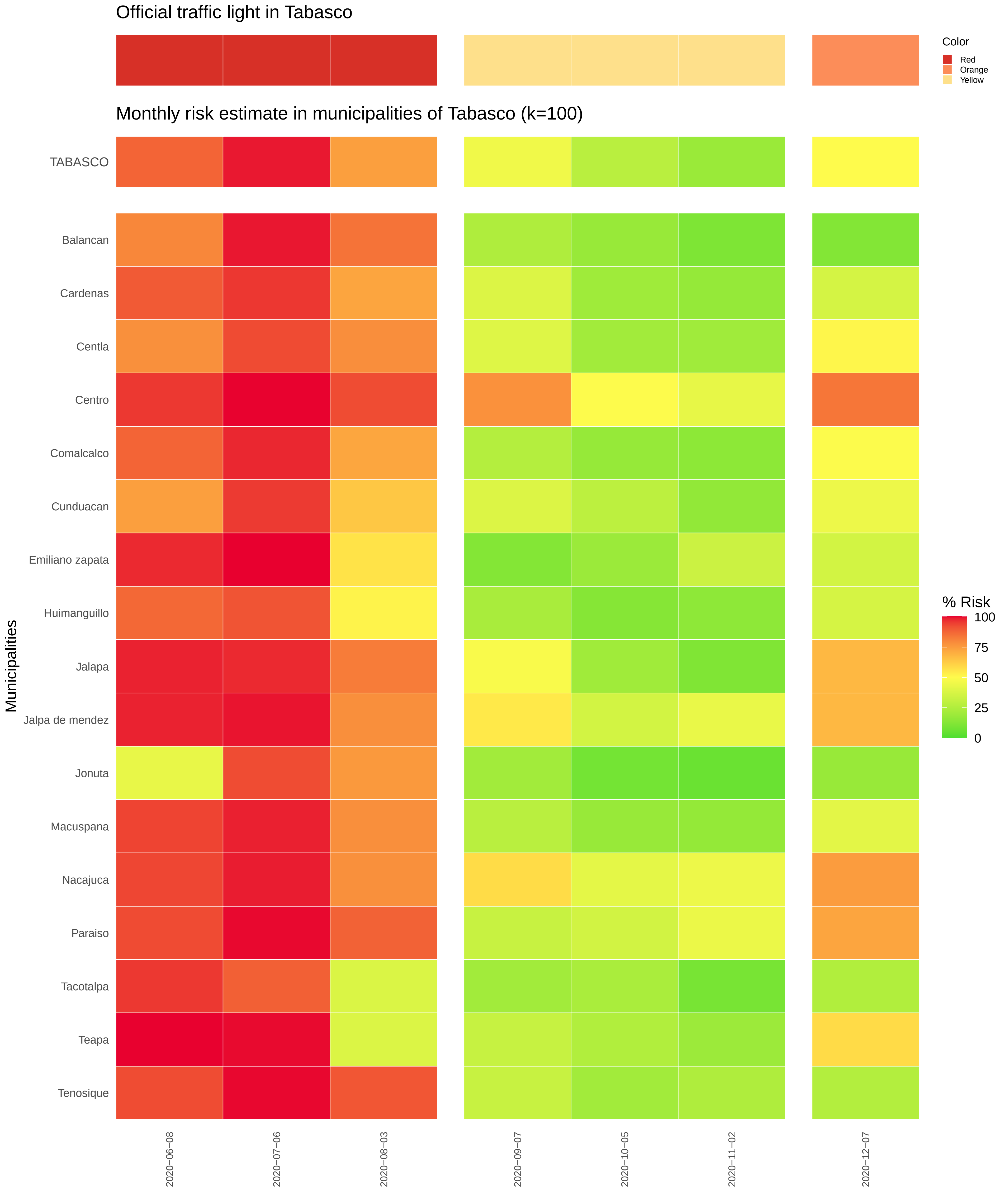
Official traffic light in Tabasco
Color
Red
Orange
Yellow
Monthly risk estimate in municipalities of Tabasco (k=100)
TABASCO
Balancan
Cardenas
Centla
Centro
Comalcalco
Cunduacan
Emiliano zapata
Huimanguillo
Jalapa
Jalpa de mendez
Jonuta
Macuspana
Nacajuca
Paraiso
Tacotalpa
Teapa
Tenosique
Municipalities
% Risk
100
75
50
25
0
2020−06−08
2020−07−06
2020−08−03
2020−09−07
2020−10−05
2020−11−02
2020−12−07

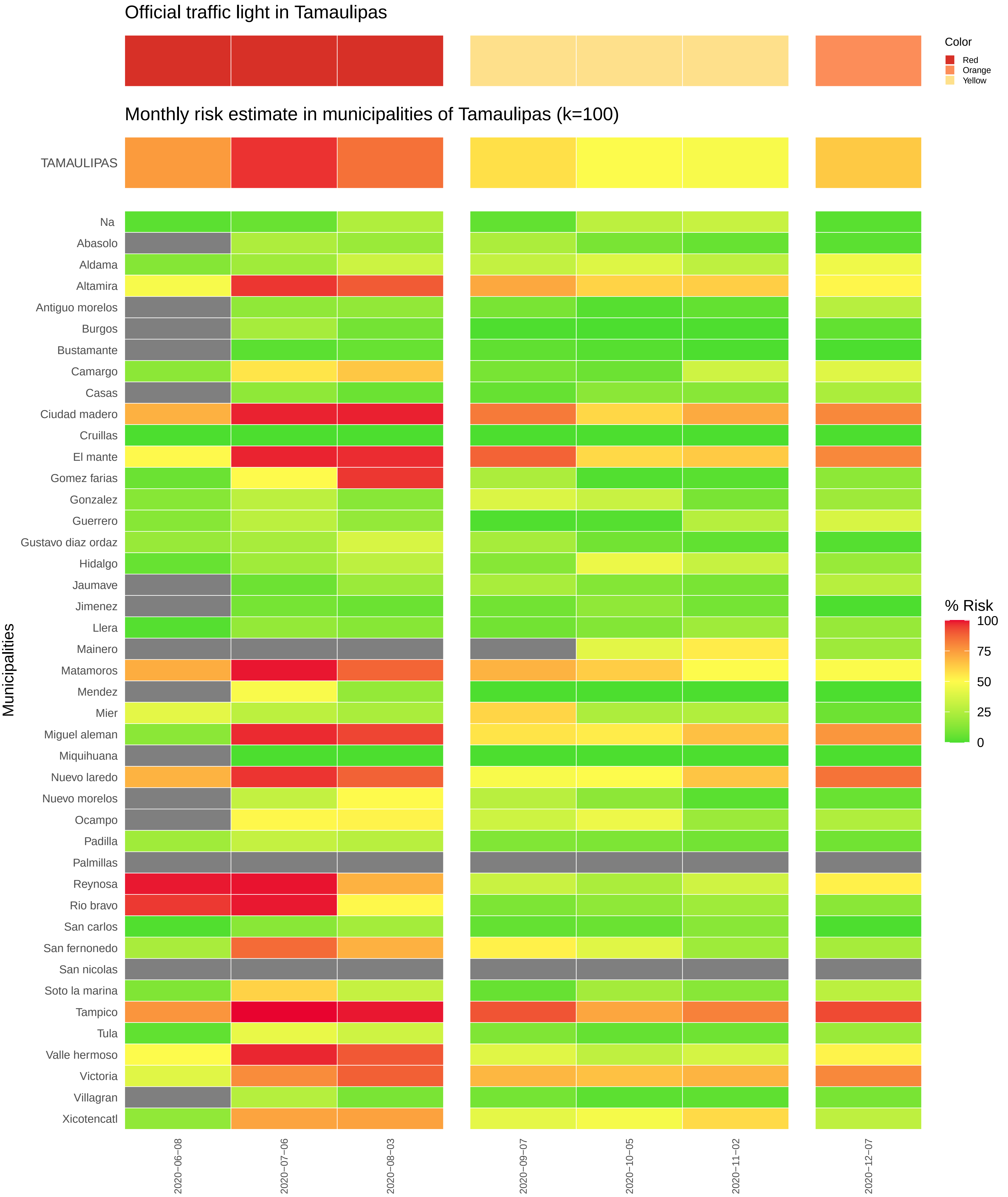
Official traffic light in Tamaulipas
Color
Red
Orange
Yellow
Monthly risk estimate in municipalities of Tamaulipas (k=100)
TAMAULIPAS
Municipalities
Na
Abasolo
Aldama
Altamira
Antiguo morelos
Burgos
Bustamante
Camargo
Casas
Ciudad madero
Cruillas
El mante
Gomez farias
Gonzalez
Guerrero
Gustavo diaz ordaz
Hidalgo
Jaumave
Jimenez
Llera
Mainero
Matamoros
Mendez
Mier
Miguel aleman
Miquihuana
Nuevo laredo
Nuevo morelos
Ocampo
Padilla
Palmillas
Reynosa
Rio bravo
San carlos
San fernonedo
San nicolas
Soto la marina
Tampico
Tula
Valle hermoso
Victoria
Villagran
Xicotencatl
% Risk
100
75
50
25
0
2020−06−08
2020−07−06
2020−08−03
2020−09−07
2020−10−05
2020−11−02
2020−12−07

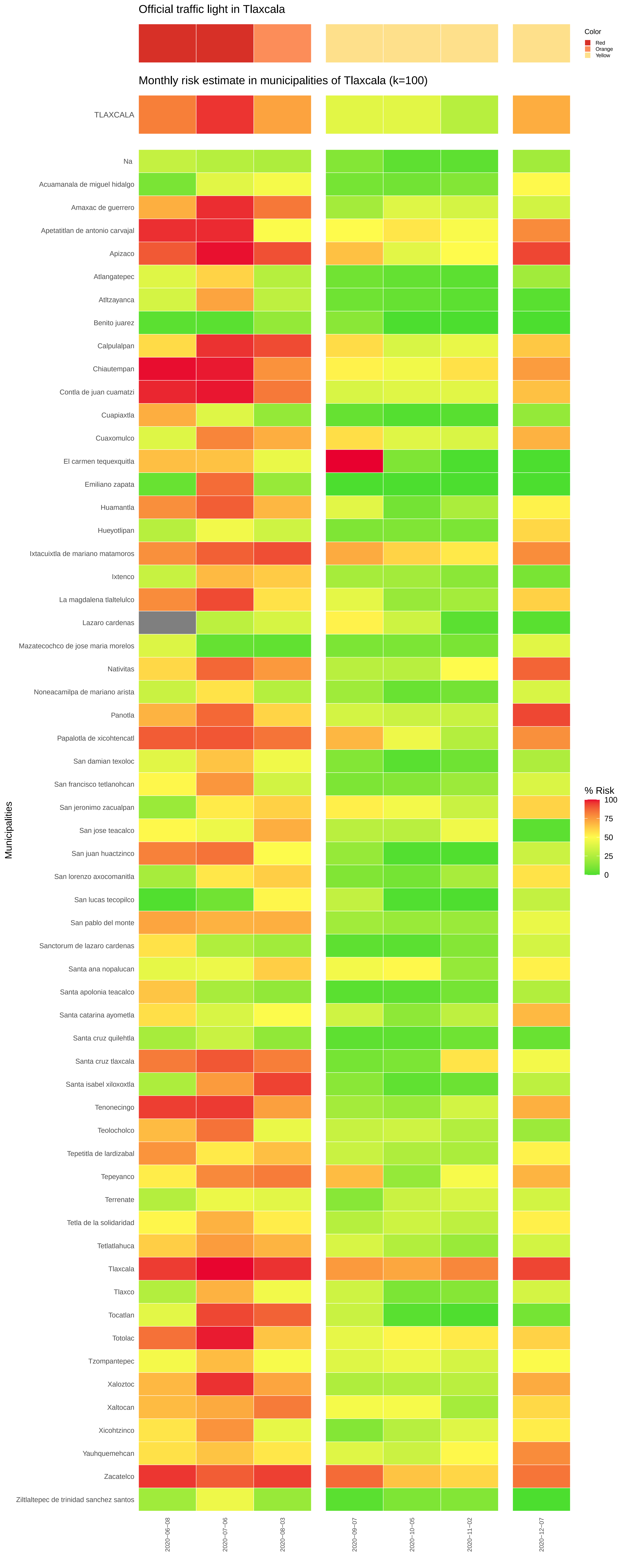

Official traffic light in Tlaxcala
Color
Red
Orange
Yellow
Monthly risk estimate in municipalities of Tlaxcala (k=100)
TLAXCALA
Na
Acuamanala de miguel hidalgo
Amaxac de guerrero
Apetatitlan de antonio carvajal
Apizaco
Atlangatepec
Atltzayanca
Benito juarez
Calpulalpan
Chiautempan
Contla de juan cuamatzi
Cuapiaxtla
Cuaxomulco
El carmen tequexquitla
Emiliano zapata
Huamantla
Hueyotlipan
Ixtacuixtla de mariano matamoros
Ixtenco
La magdalena tlaltelulco
Lazaro cardenas
Mazatecochco de jose maria morelos
Nativitas
Noneacamilpa de mariano arista
Panotla
Papalotla de xicohtencatl
San damian texoloc
San francisco tetlanohcan
San jeronimo zacualpan
San jose teacalco
San juan huactzinco
San lorenzo axocomanitla
San lucas tecopilco
San pablo del monte
Sanctorum de lazaro cardenas
Santa ana nopalucan
Santa apolonia teacalco
Santa catarina ayometla
Santa cruz quilehtla
Santa cruz tlaxcala
Santa isabel xiloxoxtla
Tenonecingo
Teolocholco
Tepetitla de lardizabal
Tepeyanco
Terrenate
Tetla de la solidaridad
Tetlatlahuca
Tlaxcala
Tlaxco
Tocatlan
Totolac
Tzompantepec
Xaloztoc
Xaltocan
Xicohtzinco
Yauhquemehcan
Zacatelco
Ziltlaltepec de trinidad sanchez santos
Municipalities
% Risk
100
75
50
25
0
2020-06-08
2020-07-06
2020-08-03
2020-09-07
2020-10-05
2020-11-02
2020-12-07

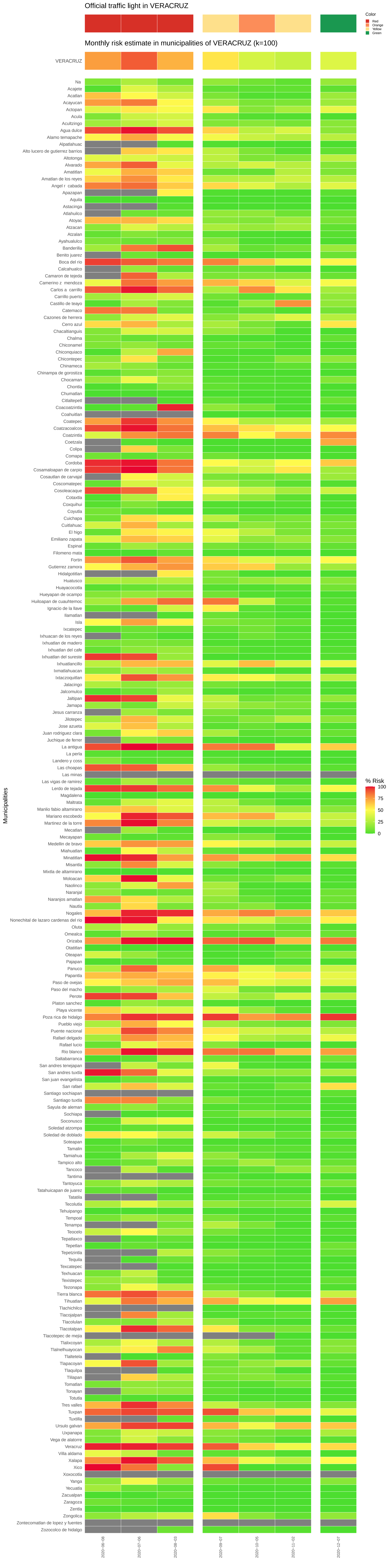
Official traffic light in VERACRUZ
Monthly risk estimate in municipalities of VERACRUZ (k=100)
Color
Red
Orange
Yellow
Green
VERACRUZ
Municipalities
% Risk
100
75
50
25
0
Na
Acajete
Acatlan
Acayucan
Actopan
Acula
Acultzingo
Agua dulce
Alamo temapache
Alpatlahuac
Alto lucero de gutierrez barrios
Altotonga
Alvarado
Amatitlan
Amatlan de los reyes
Angel r cabada
Apazapan
Aquila
Astacinga
Atlahuilco
Atoyac
Atzacan
Atzalan
Ayahualulco
Banderilla
Benito juarez
Boca del rio
Calcahualco
Camaron de tejeda
Camerino z mendoza
Carlos a carrillo
Carrillo puerto
Castillo de teayo
Catemaco
Cazones de herrera
Cerro azul
Chacaltianguis
Chalma
Chiconamel
Chiconquiaco
Chicontepec
Chinameca
Chinampa de gorostiza
Chocaman
Chontla
Chumatlan
Citlaltepetl
Coacoatzintla
Coahuitlan
Coatepec
Coatzacoalcos
Coatzintla
Coetzala
Colipa
Comapa
Cordoba
Cosamaloapan de carpio
Cosautlan de carvajal
Coscomatepec
Cosoleacaque
Cotaxtla
Coxquihui
Coyutla
Cuichapa
Cuitlahuac
El higo
Emiliano zapata
Espinal
Filomeno mata
Fortin
Gutierrez zamora
Hidalgotitlan
Huatusco
Huayacocotla
Hueyapan de ocampo
Huiloapan de cuauhtemoc
Ignacio de la llave
Ilamatlan
Isla
Ixcatepec
Ixhuacan de los reyes
Ixhuatlan de madero
Ixhuatlan del cafe
Ixhuatlan del sureste
Ixhuatlancillo
Ixmatlahuacan
Ixtaczoquitlan
Jalacingo
Jalcomulco
Jaltipan
Jamapa
Jesus carranza
Jilotepec
Jose azueta
Juan rodriguez clara
Juchique de ferrer
La antigua
La perla
Landero y coss
Las choapas
Las minas
Las vigas de ramirez
Lerdo de tejada
Magdalena
Maltrata
Manlio fabio altamirano
Mariano escobedo
Martinez de la torre
Mecatlan
Mecayapan
Medellin de bravo
Miahuatlan
Minatitlan
Misantla
Mixtla de altamirano
Moloacan
Naolinco
Naranjal
Naranjos amatlan
Nautla
Nogales
Nonechital de lazaro cardenas del rio
Oluta
Omealca
Orizaba
Otatitlan
Oteapan
Pajapan
Panuco
Papantla
Paso de ovejas
Paso del macho
Perote
Platon sanchez
Playa vicente
Poza rica de hidalgo
Pueblo viejo
Puente nacional
Rafael delgado
Rafael lucio
Rio blanco
Saltabarranca
San andres tenejapan
San andres tuxtla
San juan evangelista
San rafael
Santiago sochiapan
Santiago tuxtla
Sayula de aleman
Sochiapa
Soconusco
Soledad atzompa
Soledad de doblado
Soteapan
Tamalin
Tamiahua
Tampico alto
Tancoco
Tantima
Tantoyuca
Tatahuicapan de juarez
Tatatila
Tecolutla
Tehuipango
Tempoal
Tenampa
Tenochtitlan
Teocelo
Tepatlaxco
Tepetlan
Tepetzintla
Tequila
Texcatepec
Texhuacan
Texistepec
Tezonapa
Tierra blanca
Tihuatlan
Tlachichilco
Tlacojalpan
Tlacolulan
Tlacotalpan
Tlacotepec de mejia
Tlalixcoyan
Tlalnelhuayocan
Tlaltetela
Tlapacoyan
Tlaquilpa
Tlilapan
Tomatlan
Tonayan
Totutla
Tres valles
Tuxpan
Tuxtilla
Ursulo galvan
Uxpanapa
Vega de alatorre
Veracruz
Villa aldama
Xalapa
Xico
Xoxocotla
Yanga
Yecuatla
Zacualpan
Zaragoza
Zentla
Zongolica
Zontecomatlan de lopez y fuentes
Zozocolco de hidalgo
2020-06-08
2020-07-06
2020-08-03
2020-09-07
2020-10-05
2020-11-02
2020-12-07

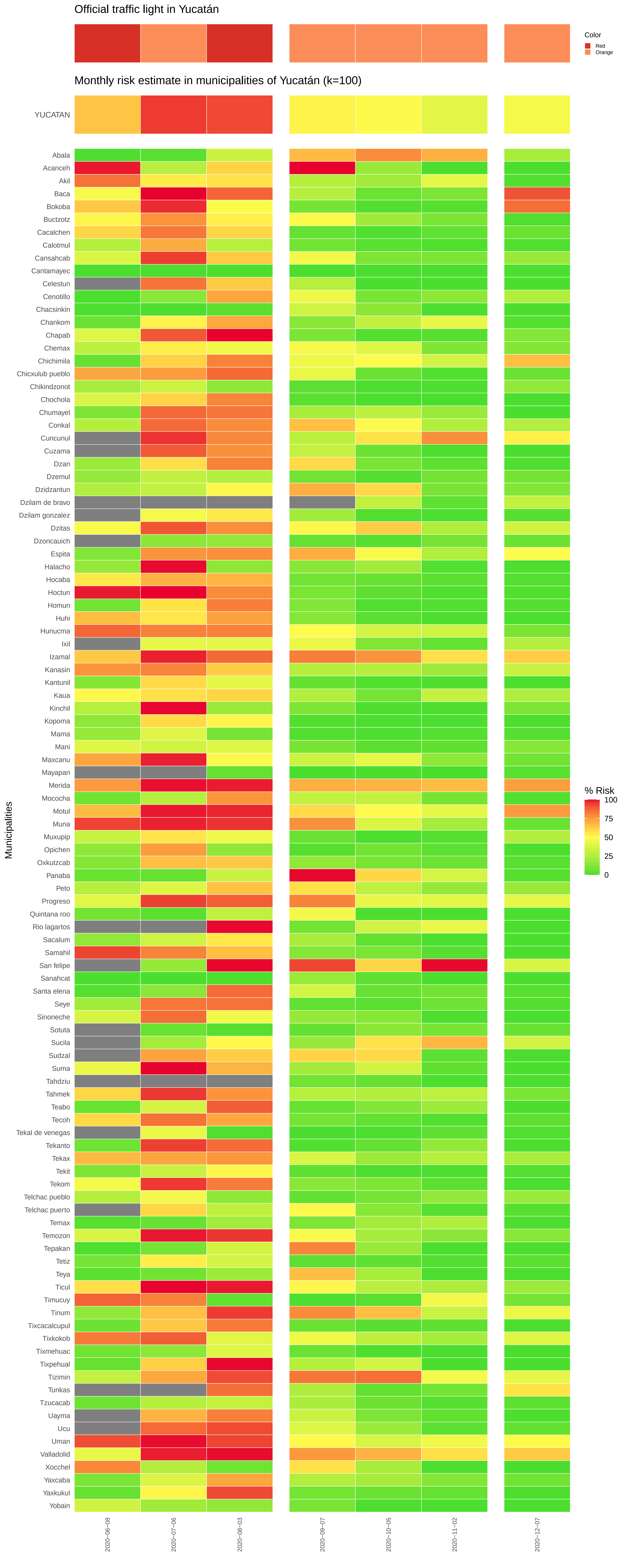
Official traffic light in Yucatán
Color
Red
Orange
Monthly risk estimate in municipalities of Yucatán (k=100)
YUCATAN
% Risk
100
75
50
25
0
Municipalities
Abala
Acanceh
Akil
Baca
Bokoba
Buctzotz
Cacalchen
Calotmul
Cansahcab
Cantamayec
Celestun
Cenotillo
Chacsinkin
Chankom
Chapab
Chemax
Chichimila
Chicxulub pueblo
Chikindzonot
Chochola
Chumayel
Conkal
Cuncunul
Cuzama
Dzan
Dzemul
Dzidzantun
Dzilam de bravo
Dzilam gonzalez
Dzitas
Dzoncauich
Espita
Halacho
Hocaba
Hoctun
Homun
Huhi
Hunucma
Ixil
Izamal
Kanasin
Kantunil
Kaua
Kinchil
Kopoma
Mama
Mani
Maxcanu
Mayapan
Merida
Mococha
Motul
Muna
Muxupip
Opichen
Oxkutzcab
Panaba
Peto
Progreso
Quintana roo
Rio lagartos
Sacalum
Samahil
San felipe
Sanahcat
Santa elena
Seye
Sinoneche
Sotuta
Sucila
Sudzal
Suma
Tahdziu
Tahmek
Teabo
Tecoh
Tekal de venegas
Tekanto
Tekax
Tekit
Tekom
Telchac pueblo
Telchac puerto
Temax
Temozon
Tepakan
Tetiz
Teya
Ticul
Timucuy
Tinum
Tixcacalcupul
Tixkokob
Tixmehuac
Tixpehual
Tizimin
Tunkas
Tzucacab
Uayma
Ucu
Uman
Valladolid
Xocchel
Yaxcaba
Yaxkukul
Yobain
2020−06−08
2020−07−06
2020−08−03
2020−09−07
2020−10−05
2020−11−02
2020−12−07

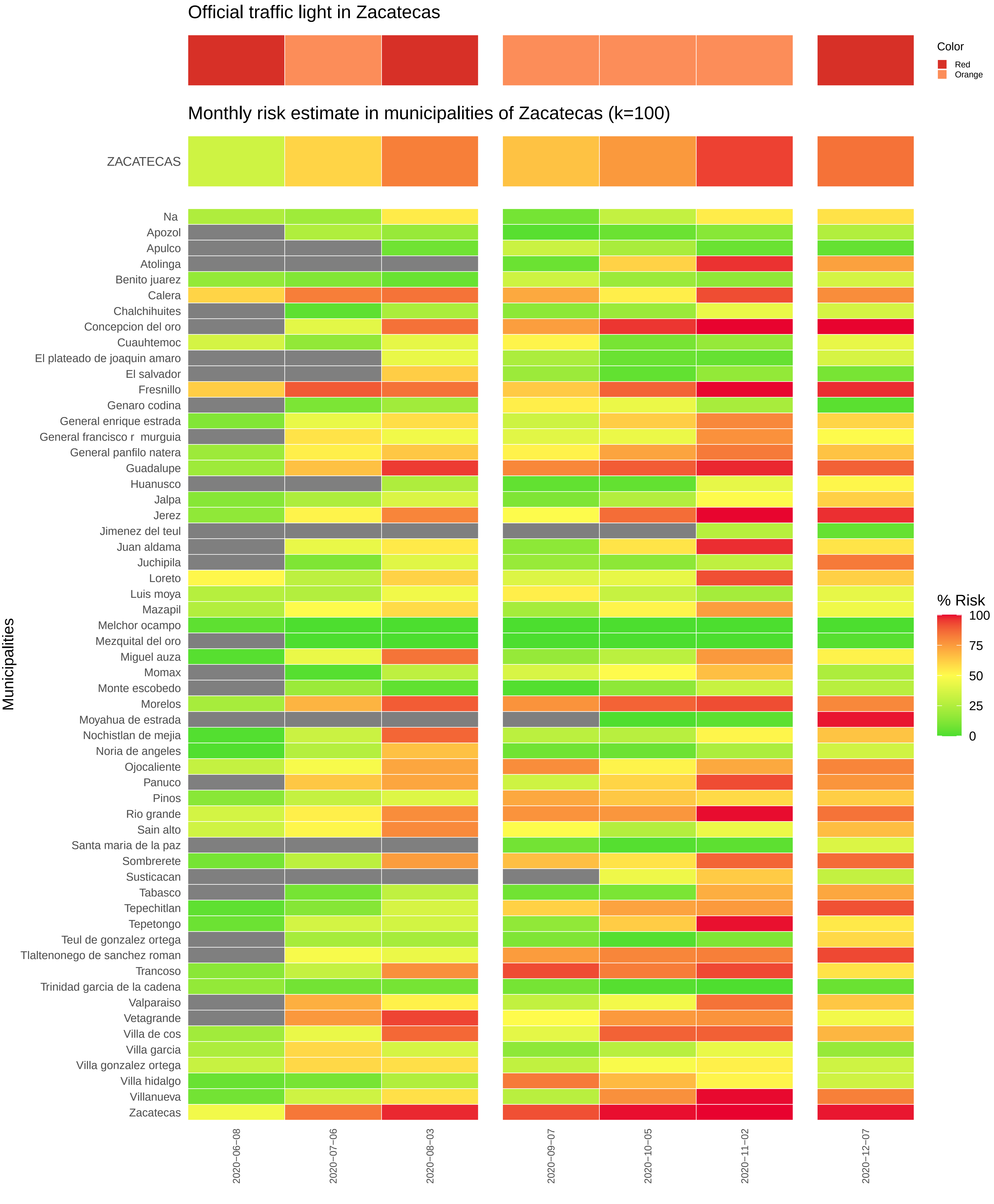
Official traffic light in Zacatecas
Color
Red
Orange
Monthly risk estimate in municipalities of Zacatecas (k=100)
ZACATECAS
Municipalities
Na
Apozol
Apulco
Atolinga
Benito juarez
Calera
Chalchihuites
Concepcion del oro
Cuauhtemoc
El plateado de joaquin amaro
El salvador
Fresnillo
Genaro codina
General enrique estrada
General francisco r murguia
General panfilo natera
Guadalupe
Huanusco
Jalpa
Jerez
Jimenez del teul
Juan aldama
Juchipila
Loreto
Luis moya
Mazapil
Melchor ocampo
Mezquital del oro
Miguel auza
Momax
Monte escobedo
Morelos
Moyahua de estrada
Nochistlan de mejia
Noria de angeles
Ojocaliente
Panuco
Pinos
Rio grande
Sain alto
Santa maria de la paz
Sombrerete
Susticacan
Tabasco
Tepechitlan
Tepetongo
Teul de gonzalez ortega
Tlaltenonego de sanchez roman
Trancoso
Trinidad garcia de la cadena
Valparaiso
Vetagrande
Villa de cos
Villa garcia
Villa gonzalez ortega
Villa hidalgo
Villanueva
Zacatecas
% Risk
100
75
50
25
0
2020−06−08
2020−07−06
2020−08−03
2020−09−07
2020−10−05
2020−11−02
2020−12−07
